# Partitioning the Genomic Components of Behavioral Disinhibition and Substance Use (Disorder) Using Genomic Structural Equation Modeling

**DOI:** 10.1101/2024.02.20.24303036

**Authors:** Tanya B. Horwitz, Katerina Zorina-Lichtenwalter, Daniel E. Gustavson, Andrew D. Grotzinger, Michael C. Stallings

**Affiliations:** Institute for Behavioral Genetics, University of Colorado Boulder, 1480 30th St, Boulder, CO, United States of America 80303; Psychology and Neuroscience, University of Colorado Boulder, Meunzinger D244, 345 UCB, Boulder, CO, United States of America 80303

**Keywords:** Substance use, externalizing, behavioral genetics, GenomicSEM

## Abstract

Externalizing behaviors encompass manifestations of risk-taking, self-regulation, aggression, sensation-/reward-seeking, and impulsivity. Externalizing research often includes substance use (SU), substance use disorder (SUD), and other (non-SU/SUD) “behavioral disinhibition” (BD) traits. Genome-wide and twin research have pointed to overlapping genetic architecture within and across SUB, SUD, and BD. We created single-factor measurement models—each describing SUB, SUD, or BD traits--based on mutually exclusive sets of European ancestry genome-wide association study (GWAS) statistics exploring externalizing variables. We then applied trivariate Cholesky decomposition to these factors in order to identify BD-specific genomic variation and assess the partitioning of BD’s genetic covariance with each of the other facets. Even when the residuals for indicators relating to the same substance were correlated across the SUB and SUD factors, the two factors yielded a large zero-order correlation (r_g_=.803). BD correlated strongly with the SUD (r_g_=.774) and SUB factors (r_g_=.778). In our initial decompositions, 33% of total BD variance remained after removing variance associated with SUD and SUB. The majority of covariance between BD and SU and between BD and SUD was shared across all factors. When only nicotine/tobacco, cannabis, and alcohol were included for the SUB/SUD factors, their zero-order correlation increased to r_g_=.861; in corresponding decompositions, BD-specific variance decreased to 27%. In summary, BD, SU, and SUD were highly genetically correlated at the latent factor level, and a significant minority of genomic BD variation was not shared with SU and/or SUD. Further research can better elucidate the properties of BD-specific variation by exploring its genetic/molecular correlates.

## Introduction

### 1.1 Background

Behavioral researchers have traditionally placed phenotypes that relate to risk-taking, lack of self-regulation, aggression, and/or impulsivity on the “externalizing” psychopathology spectrum. Related behaviors that exemplify sensation- or reward-seeking without meeting diagnostic psychopathology criteria have likewise been grouped under the externalizing framework (Karlsson Linnér et al., 2021). Studies that probe externalizing often consider substance use disorders (SUDs), which are characterized by obsessive/compulsive substance use-seeking and -intake patterns, negative emotional responses to cessation of the substance (Koob and Volkow, 2016), and other related symptoms (Kendler and Myers, 2014; Krueger et al., 2005). However, substance consumption behaviors not associated with these life-complicating factors (i.e., substance initiation and intake quantity/frequency; hereafter, SUB), have also been considered in externalizing research (Karlsson Linnér et al., 2021). Externalizing behavior additionally encompasses a combination of several pathological and non-pathological traits—i.e., risk tolerance, number of sexual partners, age at first sex (Karlsson Linnér et al., 2021), antisocial personality disorder, and conduct disorder (Kendler and Myers, 2014; Krueger et al., 2005)--that do not directly measure substance use or substance use disorders, which we will refer to as “behavioral disinhibition” (BD) (Poore et al., 2023).

Genome-wide association studies (GWAS) and twin studies have shown traits and latent constructs related to SUB, SUD, and BD to be genetically correlated, suggesting that these constructs have overlapping genetic architectures (Karlsson Linnér et al., 2021; Kendler and Myers, 2014; Poore et al., 2023). There is evidence for “general” genetic liability underlying SUD and for additional genetic liability specific to certain individual SUDs (Hatoum et al., 2022). Though SUDs examined in large GWAS are unsurprisingly genetically correlated with use measures for the corresponding substance, the degree of genetic overlap varies across substances. For example, while the point estimate of the genetic correlation between the Fagerström Test for Nicotine Dependence (FTND) and number of cigarettes smoked per day is around unity (r_g_ = .97, s.e.= .12) (Hatoum et al., 2022), the correlation between cannabis use and cannabis use disorder is lower, at r_g_ = .50 (s.e. = .05) (Johnson et al., 2020). In addition to twin and family studies that have explored the genetics of externalizing based on phenotypic resemblance among relatives, a recent multivariate analysis estimated a general genomic externalizing factor using summary association data from seven high-powered GWAS (Karlsson Linnér et al., 2021), a design that evaluates the relationship between measured genotypes and a phenotype of interest. Of these seven GWAS, one examined alcohol use disorder, two examined non-pathological substance use traits (cannabis initiation and smoking initiation), and four examined traits that could be classified as forms of BD (age at first sex, number of sexual partners, attention-deficit/hyperactivity disorder, and risk tolerance). This study identified 579 independent genomic loci associated with general externalizing. A more recent publication reported a broader externalizing factor that included additional SUD constructs (Poore et al., 2023). Factor-level analysis of general genomic SUD liability (Hatoum et al., 2023) has yielded a substantially narrower set of associated genome-wide significant loci than did the seven-indicator externalizing factor (though the externalizing factor was likely better-powered). Further research is required to determine the extent to which genomic variation associated with facets of externalizing overlaps and the extent to which it operates through independent pathways.

### 1.2 Broad Analysis Plan

In the present study, we used genome-wide association data and Genomic Structural Equation Modeling software (Grotzinger et al., 2019) to estimate genetic correlations between externalizing traits, extract latent factors underlying three externalizing facets (SUD, SUB, and BD), and decompose their shared and unshared genetic variance via hierarchical trivariate Cholesky models. The findings from this analysis will shed light on whether phenotypic subcategories of externalizing behavior constitute separable genomic entities.

## Material and Methods

### 2.1 Description of Genomic Structural Equation Modeling

In accordance with our preregistered plan (https://osf.io/fjy5h), we used the software Genomic Structural Equation Modeling (Genomic SEM) (Grotzinger et al., 2019) to model the genomic factor-level structure of general and domain-specific externalizing. Genomic SEM leverages LD Score regression (LDSC) (Bulik-Sullivan et al., 2015b), lavaan (Rosseel et al., 2023), and GWAS summary statistics to fit confirmatory models based on genetic (rather than phenotypic) correlations.

In the first stage of Genomic SEM, LDSC creates an empirical genetic covariance matrix (*S*_LDSC_) and an associated sampling covariance matrix (*V*_S_LDSC_). The unstandardized *S* matrix contains single nucleotide polymorphism (SNP) heritabilities—the proportion of phenotypic variance that can be statistically explained by SNPs--of each phenotype on the diagonals and genetic covariances between each phenotype on the off-diagonals. Meanwhile, the *V* matrix is a sampling covariance matrix used to account for study sample overlap among the source GWAS summary statistics. In the second stage, Genomic SEM fits a user-specified structural equation model, estimating factor loading and correlation parameters that minimize discrepancy between the model-implied genetic covariance matrix and the empirical covariance matrix.

### 2.2 Measures

We began by defining three lower-order general factors (the measurement models): Behavioral Disinhibition (BD), (non-pathological) substance use (SUB), and Substance Use Disorder (SUD)—each representing a dimension of externalizing and each comprising mutually exclusive groups of reflective indicators—with the intention of investigating how SUB and SUD may relate to BD differently. Per our pre-registration, we only included GWAS for traits with sample sizes of at least 10,000 and SNP heritability (h^2^SNP) Z-statistics > 4 (Bulik-Sullivan et al., 2015a). For traits that were meta-analyzed but whose meta-analyzed association statistics were not available, we ran inverse-variance weighted meta-analysis ourselves in METAL (Willer et al., 2010). All samples were comprised of individuals of European ancestry, as non-European ancestry groups are under-powered for some of the indicators of interest and may have different patterns of linkage disequilibrium and allele frequencies (Peterson et al., 2019). Table 1 displays a summary of the four GWAS used for the BD factor, the four GWAS used for the SUD factor, and the seven pre-registered GWAS initially explored for the SUB factor, as well as their associated publications.

**Table 1.**
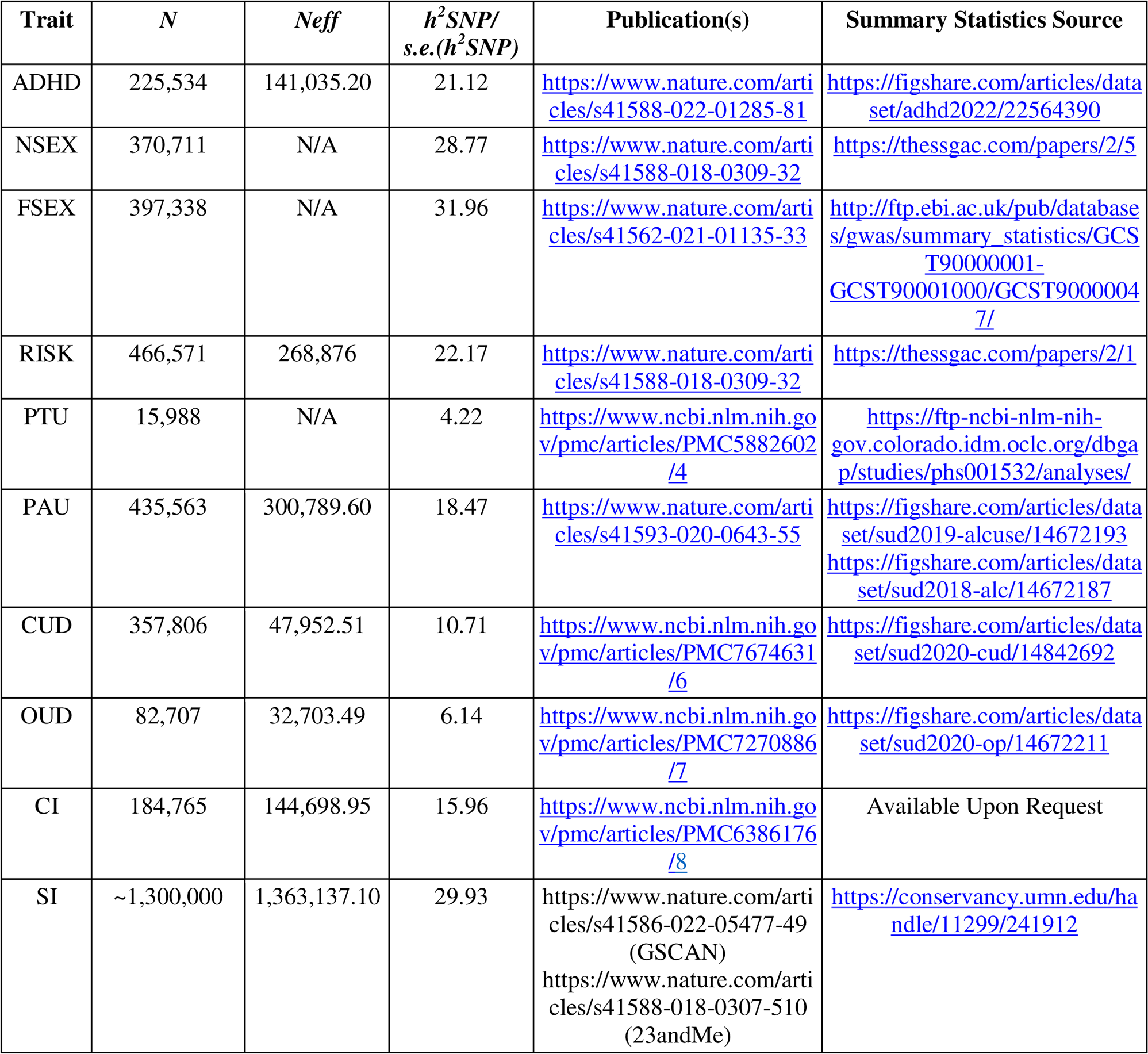
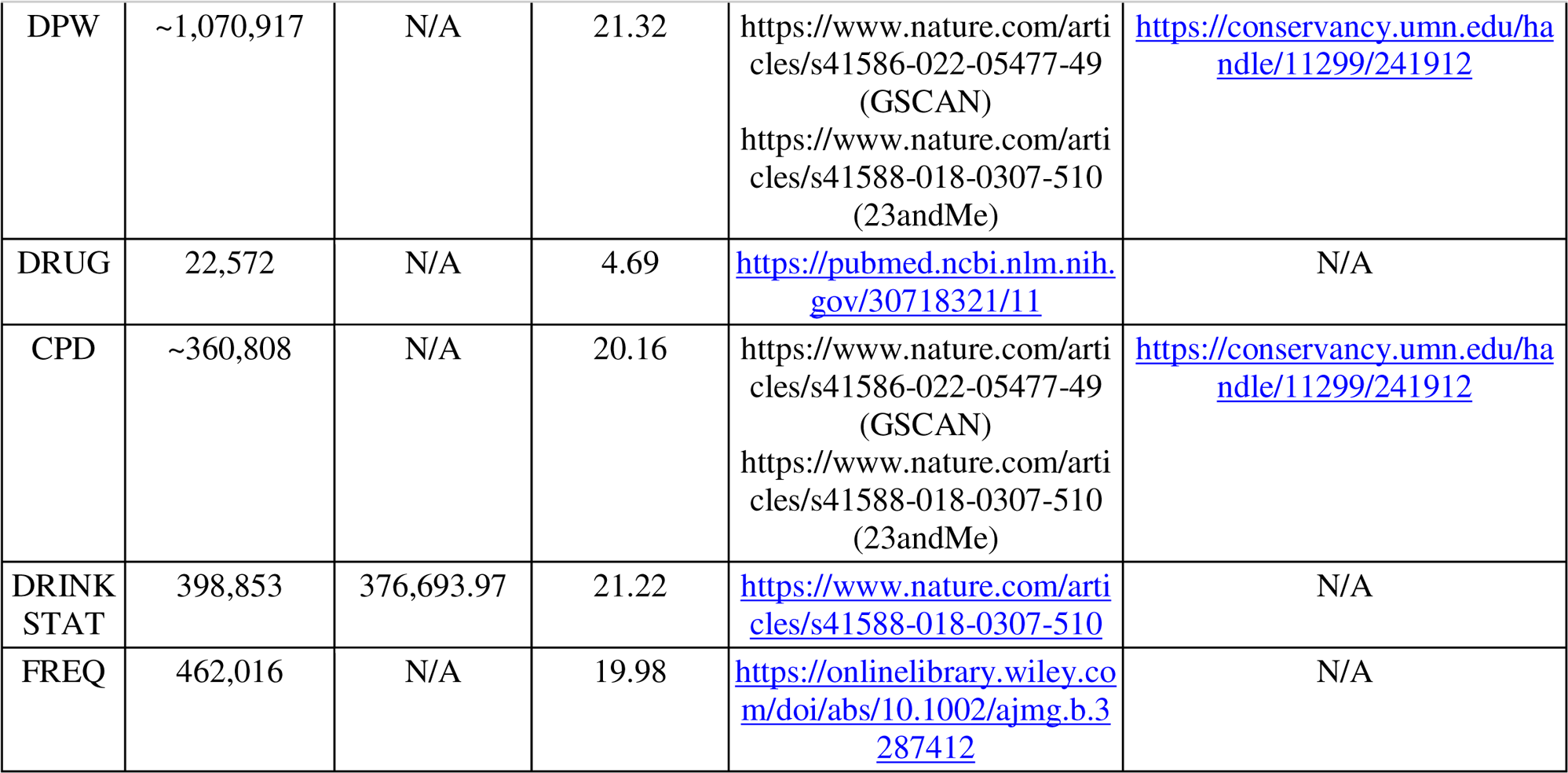
Summary data, associated publications, and data sources for the four Behavioral Disinhibition traits, four Substance Use Disorder traits, and seven pre-registered Substance Use traits initially considered for the Substance Use Factor. *N =* raw sample size used. Note that, in some cases, sample sizes in publicly available data are lower than total sample size reported in the corresponding publication(s) or in the table because of data access restrictions on some cohorts. *N_eff_* = effective sample size (for dichotomous traits), as provided in the summary statistics or calculated with respect to prevalence and sample size across cohorts. *h*^2^ = the proportion of trait variation in the sample(s) accounted for by variation in single nucleotide polymorphisms (SNPs). See sections 2.2.1-2.2.3 for further descriptions of phenotypes. ADHD = Attention-deficit Hyperactivity Disorder, NSEX = Number of Lifetime Sexual Partners, FSEX = Age at First Sexual Intercourse (reverse-coded), RISK = General Risk Tolerance, PTU = Problem Tobacco Use, PAU = Problem Alcohol Use, CUD = Cannabis Use Disorder, OUD = Opioid Use Disorder, CI = Cannabis Initiation, SI = Smoking Initiation, DPW = Drinks per Week, DRUG = Drug Experimentation (the number of different classes of drugs an individual has used out of eleven), CPD = Cigarettes per Day, DRINK STAT = Drinking Status (drinker vs. non-drinker), FREQ = Drinking Frequency.

#### 2.2.1 BD Factor

The lower-order BD factor utilized the four (non-substance-associated) BD indicators established in Poore *et al*. (2023):

- **Attention-deficit/hyperactivity disorder (ADHD)** 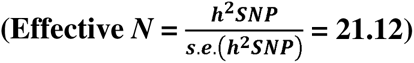, as defined by the International Classification of Diseases (ICD)-10 (with some samples taken from an inpatient or outpatient psychiatric setting), or prescription for an ADHD medication, depending on the cohort; ADHD for the Psychiatric Genomics Consortium was diagnosed using multiple different measures (case/control) (Demontis et al., 2023).
- **Number of lifetime sexual partners (NSEX)** (Karlsson Linnér et al., 2019) 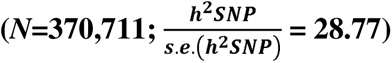
- **Age at first sexual intercourse (FSEX)** (reverse-coded) (Mills et al., 2021) 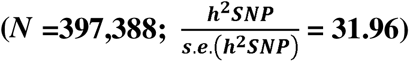
- **General risk tolerance (RISK)** 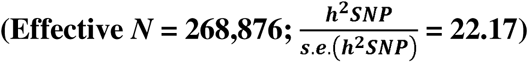, which was based on several items that varied depending on the cohort and that were similar to the question: “Would you describe yourself as someone who takes risks?” (Karlsson Linnér et al., 2019)

#### 2.2.2 SUD Factor

The lower-order SUD factor described the indicators in Hatoum *et al*. (2022) and Hatoum *et al*. (2023):

- **Problem alcohol use (PAU)** 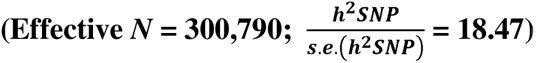, defined by alcohol dependence in the fourth edition of the Diagnostic and Statistical Manual of Mental Disorders (DSM-IV) or alcohol use disorder, as determined by the Alcohol Use Disorders Identification Test problem items (AUDIT-P), a continuous metric, or by the ICD-9 or -10, depending on the cohort (Zhou et al., 2020b).
- **Opioid use disorder (OUD)** 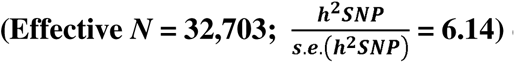 defined by at least one inpatient or two outpatient ICD-9 or ICD-10 codes, or by opioid dependence, as defined by the DSM-IV, depending on the cohort (case vs. exposed control) (Zhou et al., 2020a).
- **Cannabis use disorder (CUD)** 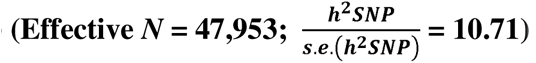, defined by cannabis abuse or dependence according to the DSM-III or DSM-IV, cannabis abuse or dependence according to the ICD-10, or cannabis use disorder according to the DSM-5, depending on the cohort (case vs. exposed *or* unexposed control) (Johnson et al., 2020).
- **Problem tobacco use (PTU)** 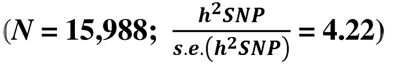, based on the Fagerström Test for Nicotine Dependence (FTND) in ever-smokers (Hancock et al., 2018). Unlike in the Hatoum et al. study, we only used the FTND—which is used to determine degree of nicotine dependence (mild, moderate, or severe)--as a measure of PTU and did not integrate cigarettes per day into the indicator. We also had a smaller sample size for the FTND GWAS because of data restrictions.

#### 2.2.3 SUB Factor

We initially explored a total of seven pre-registered non-pathological substance use measures, three of which were not ultimately included. Thus, the lower-order SUB factor was based around four indicators:

- **Lifetime cannabis initiation (CI)** 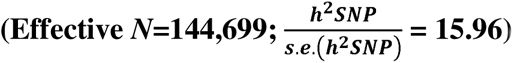, which was based on several items that varied depending on the cohort. For example: “Have you ever in your life used the following: Marijuana?”; “Have you taken CANNABIS (marijuana, grass, hash, ganja, blow, draw, skunk, weed, spliff, dope), even if it was a long time ago?” (Pasman et al., 2018)
- **Smoking initiation (SI)** 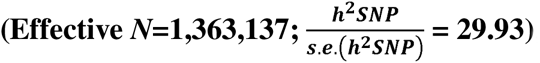, as defined by having ever or never been a regular smoker, or having ever smoked 100 or more cigarettes in one’s lifetime, depending on the cohort (Liu et al., 2019; Saunders et al., 2022).
- **Drinks per Week (DPW)** 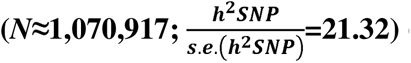 (Liu et al., 2019; Saunders et al., 2022).
- **Drug Experimentation (DRUG)** 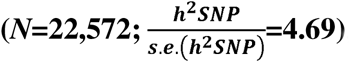, the number of different classes of drugs an individual has used out of eleven (Sanchez-Roige et al., 2019a).

### 2.3 Preparation of summary statistics for Genomic SEM

We formatted downloaded summary data from relevant GWAS using Genomic SEM’s munge() function. We filtered summary statistics for SNPs available in the reference dataset, HapMap3, and further restricted to SNPs with a minimum minor allele frequency (MAF) of .01 and a minimum imputation quality (INFO) score of .90 (except for opioid use disorder, for which we used a minimum INFO threshold of .70) (Hatoum et al., 2023, 2022; Poore et al., 2023). Additionally, for each dichotomous trait, we used cohort-specific prevalence rates to calculate the sum of effective sample sizes (*N*); effective *N* corresponds to the *N* for a GWAS with equal power to that of the cohort’s raw sample size in a 1:1 case:control cohort design and is calculated as 4*v_k_*(1-*v_k_*)*n_k_*, where *v* is equivalent to cohort-specific prevalence and *n* is equivalent to the cohort’s raw sample size (Grotzinger et al., 2023). Thus, effective *N* corrects for ascertainment in studies that disproportionately sample for cases. Using the ldsc() function, we then calculated pairwise genetic covariances for all traits using the 1000 Genomes Phase 3 European ancestry linkage disequilibrium (LD) scores as a reference (Bulik-Sullivan et al., 2015a; Bulik-Sullivan et al., 2015b; The 1000 Genomes Project Consortium, 2015). Figure 1 shows the unstandardized genetic covariances (in the lower triangle) and genetic correlations (in the upper triangle) among the twelve traits we included in our main models, along with SNP-based heritabilities on the diagonal. An expanded genetic covariance/correlation matrix that additionally includes the pre-registered SUB indicators that were excluded from the final models is shown in Supplementary Figure 1.

### 2.4 Factor Analysis

Next, we used Genomic SEM to specify models with the three externalizing factors (SUD, SUB, and BD) as lower-order factors in the ensuing structural models. In order to extract influences of within-substance intercorrelations, we correlated the residuals of PTU and SI, CUD and CI, and PAU and DPW across the SUD and SUB factors. We constructed a correlated factors model including all three factors. Then, we integrated the lower-order factors into a hierarchical trivariate Cholesky decomposition framework. The orthogonal higher-order latent factors in the Cholesky decomposition completely broke down the genetic variance and covariance among the three lower-order latent factors into shared and independent sources of variance (Demange et al., 2021; Neale, 1992). The leftmost higher-order factor captured all genetic variance of the leftmost lower-order factor as well as its shared variance with the other lower-order factors. The middle higher-order factor captured all of the leftover genetic variance for the middle lower-order factor and its shared variance with the rightmost lower-order factor. Finally, the rightmost higher-order factor captured the genetic variance unique (in the model) to the rightmost lower-order factor. Model 1 had the following order of lower-order factors, from left-to-right: SUD, SUB, BD; and Model 2 had the order: SUB, SUD, BD. Thus, both models had BD in the final lower-order factor position. We utilized the usermodel() function in Genomic SEM to run both models using diagonally weighted least squares (DWLS) estimation. We scaled the higher-order factors using unit variance identification.

### 2.5 Model Notation

In the model descriptions, the letters in the factors’ subscripts denote the model number, where *a* corresponds to Model 1 and *b* corresponds to Model 2 in the two models that followed the pre-registered plan (additionally, *c* corresponds to Modified Model 1 and *d* to Modified Model 2 in the post-hoc models; see below). Meanwhile, the numerals in the factors’ subscripts denote the level of the factor, where 1 denotes higher-order and 0 denotes lower-order.

### 2.6 Description of Fit Metrics

Though not the primary metric of interest for the decompositions, we assessed model fit for the measurement models, the main correlated factor models, and the main Cholesky decompositions using three indices: the comparative fit index (CFI), the Akaike Information Criterion (AIC), and the Standardized Root Mean Square Residual (SRMR). In Genomic SEM, the CFI measures the improvement in fit of the specified model compared to a model that estimates heritability of phenotypes but assumes no genetic covariances between them. The AIC is a relative fit index that balances fit with parsimony, incorporating *x*^2^ and the number of free parameters in the model. The SRMR, an index of approximate model fit, is the standardized root mean square difference between the model-implied and observed correlation matrices. CFI values, which can range from 0-1, are considered acceptable at .90 or greater and good if at least .95. Models with an SRMR under .10 suggest acceptable fit, while an SRMR under .05 indicates good fit. Similarly, lower AIC’s indicate better fit (Grotzinger et al., 2019).

## Results

### 3.1 Lower-Order BD, SUD, and SUB Factors

As expected, the measurement models for the BD and SUD latent constructs had good model fit (CFI = .949, SRMR = .065, AIC = 181.793; CFI = 1.000, SRMR = .013, AIC = 16.277, respectively; Supplementary Tables S1-S2). We tested a series of models for our SUB factor (described further in the Supplementary Note; the associated parameters for these models are displayed in Supplementary Tables S3-S7). Our final four-indicator SUB measurement model (see Supplementary Table S7 and Section 2.2.3) satisfied our pre-registered criteria and achieved good model fit (CFI = .994, SRMR = .063, AIC = 26.145). Supplementary Figures 2A-2C show the measurement models for the BD, SUD, and SUB factors separately with their standardized loadings. The SUD and SUB factors each yielded one standardized loading (for OUD and DRUG, respectively) that was slightly greater than 1.00. However, these loadings dropped below 1.00 when the three latent factors were allowed to correlate with one another (Figure 2) and when they were integrated into the Cholesky models (Figures 3A-3B).

### 3.2 Correlated Factors

Before performing Cholesky decomposition, we fit a correlated factors model, which showed substantial zero-order genetic correlations between the SUD factor, the BD factor, and the SUB factor (Figure 2). The correlated factors model showed decent fit (CFI =.921, SRMR = .106, AIC =1638.065). All indicators were associated with significant moderate or high loadings, ranging from a standardized loading of .367 (s.e.=.022) for DPW to a standardized loading of .961 (s.e.=.039) for CUD. The SUB factor correlated with SUD at r_g_ = .803 (s.e. = .027) and with BD at r_g_ = .778 (s.e.= .016), and the latter two correlated at r_g_ = .774 (s.e.= .027). Of the residual correlations, only the correlation between DPW’s and PAU’s error terms was significant (at r_g_ = .666, *p* = 1.066e-34). Thus, cross-factor associations for the same substance were not a primary driver of the inter-factor correlations. Rather, the variance captured in the latent SUD and SUB factors likely reflected common genomic variation across substance classes.

### 3.3 Cholesky Decomposition

Models 1 and 2, the main Cholesky decompositions, are depicted in Figures 3A and 3B, respectively, along with loadings standardized with respect to the full model, including endogenous latent factors. Model 1 sought to capture the proportion of BD variance explained by a higher-order construction of SUD, as well as genetic covariance specific to lower-order SUB and lower-order BD independent of lower-order SUD. In Model 1, the first-position higher-order factor (SUD_G1a_) captured all of the variance in the (left-most) lower-order SUD factor (SUD_G0a_) and also captured covariance between SUD_G0a_ and the other lower-order factors in the model (SUB_G0a_ and BD_G0a_) as well as covariance between the latter two that was shared with SUD_G0a_ variance. SUBRes_G1a_ accounted for the remaining SUB_G0a_ factor variation and, by virtue of its partialing out covariance shared with SUD, also captured the independent influence of (non-pathological) substance-related indicators on BD_G0a_ in the model. Finally, BDRes_G1a_ captured the BD-specific variation remaining after both pathological and non-pathological substance use were accounted for. In Model 2, the higher-order substance use factor (SUB_G1b_) was the left-most factor such that SUB_G1b_ captured all of the variation in the corresponding lower-order factor (SUB_G0b_), while residual higher-order substance use disorder (SUDRes_G1b_) was in the middle position.

#### 3.3.1 Fit Statistics and Model Comparison

Models 1 and 2 (CFI =.921, SRMR=.106, AIC =1638.063 for Model 1, 1638.067 for Model 2) were associated with a significant improvement in fit (Δ*X*^2^(3) = 782.469 for Model 1, 782.465 for Model 2, *p* < .001) when compared to a single twelve-indicator common factor solution (CFI =.881, SRMR = .121, AIC =2414.532), the latter of which produced standardized loadings ranging from .333 (s.e.=.021) for DPW to .868 (s.e.=.063) for DRUG (see Supplementary Table S8). However, it is important to note that the chi-squared statistic is highly sensitive to small differences in Genomic SEM due to the well-powered nature of the GWAS used as input (Grotzinger et al., 2019). The full parameter output for the correlated factors model is in Supplementary Table S9, while the full parameter output for Models 1 and 2 are in Supplementary Tables S10 and S11, respectively.

#### 3.3.2 Partitioning of Variance

In Model 1, 64% of SUB_G0a_ and 60% of BD_G0a_ was shared with the higher-order SUD_G1a_ factor. Genetic variance associated with substance use but not substance use disorder (SUBRes_G1a_) explained only 7% of the total genetic variance in BD_G0a_. Approximately 33% of BD_G0a_ was not explained by either SUD_G1a_ or SUBRes_G1a_ and was therefore BD-specific. Additionally, 20% of the genetic covariance between SUB_G0a_ and BD_G0a_ was independent of SUD_G1a_. In Model 2, SUB_G1b_ explained 64% of SUD_G0b_ and 61% of BD_G0b_, while SUDRes_G1b_ accounted for 6% of BD_G0b_ variance. As in Model 1, 33% of BD_G0b_ was BD-specific. Finally, 19% of the genetic covariance between SUD_G0b_ and BD_G0b_ in Model 2 was independent of SUB_G1b_.

### 3.4 Post-hoc Models

Because the polysubstance use indicator (DRUG) in our SUB factor did not have a corresponding polysubstance indicator in the SUD factor and because the OUD indicator for SUD did not have a corresponding recreational opioid use indicator for SUB, we also tested a post-hoc iteration of our correlated factors and two initial Cholesky models that dropped the DRUG and OUD indicators. In a correlated factors model, there was a somewhat higher correlation between the modified SUD and SUB factors (r_g_ = .861, s.e. = .033). The point estimate of the correlation between BD and the modified SUD factor, r_g_ = .849 (s.e. = .035), was higher than that of the correlation between BD and the modified SUB factor (r_g_ = .773, s.e. = .017) (see Supplementary Figure 3 and Supplementary Table S12). As a result, Model 1 and Model 2 with the Modified SUB and SUD factors, when compared to the original models, yielded somewhat less BD-specific variance (27% of total BD). Additionally, Modified Model 1 produced a non-significant cross-loading (*p* = .383) of BD_G0c_ on MOD_SUBRes_G1c_. Thus, in this model, effectively all of the covariance between MOD_SUB_G0c_ and BD_G0c_ was shared with MOD_SUD_G0c_ (see Supplementary Figures 4A-4B and see Supplementary Tables S13-S14 for depictions and full parameter outputs of Modified Models 1 and 2).

When we dropped the residual correlations, the zero-order correlation between SUB and SUD increased somewhat (from .803 up to .852) (Supplementary Figure 5, Supplementary Table S15) when compared to the original correlated factors model. Correlations with BD remained essentially unchanged (from .778 to .780 for SUB; from .774 to .770 for SUD). Meanwhile, dropping the residual correlations in the modified model increased the point estimate of the correlation between MOD_SUB and MOD_SUD (from .861 up to .932), while BD’s zero-order correlations with MOD_SUB and MOD_SUD did not change notably (from .773 to .780 and from .849 to .844, respectively) (Supplementary Figure 6, Supplementary Table S16). Supplementary Figures 7A-7B and Supplementary Tables S17-S18 correspond to results for Cholesky Decompositions utilizing the original factors from Models 1 and 2 with the residual correlations dropped. Supplementary Figures 8A-8B and Supplementary Tables S19-S20 correspond to results for Cholesky Decompositions utilizing the modified factors with the residual correlations dropped.

## Discussion

### 4.1 Summary

We used Genomic SEM to partition the genome-wide components of externalizing behaviors into a four-indicator behavioral disinhibition (BD) factor, a four-indicator substance use disorder (SUD) factor, and a four-indicator (non-pathological) substance use (SUB) factor. The three factors showed high zero-order intercorrelations, even when the residuals for indicators relating to the same substance were correlated across the SUB and SUD factors. Using hierarchical trivariate Cholesky decomposition, we then analyzed the residual genomic variance components that remained for externalizing above and beyond 1) SUD, 2) SUB, and 3) both SUD and SUB. The majority of the genetic variation in BD intersected with the joint variation shared across the other two domains, which reinforces findings from past research examining genomic links between externalizing constructs (Karlsson Linnér et al., 2021; Poore et al., 2023). The covariance between BD and each substance factor independent of the other was considerably smaller than covariance shared among all three factors. The modified SUB and SUD factors, which included only the three most commonly-used recreational substance categories—nicotine/tobacco, alcohol, and cannabis—had a larger genetic correlation point estimate than did the original substance factors that included polysubstance use and opioid use disorder. Finally, across all models, between approximately a quarter and approximately a third of BD-associated genetic variance was independent of both SUB and SUD.

### 4.2 Relevance to Past Research and Future Directions

Depending on the factor definition for SUB and SUD, the point estimates of their zero-order genetic correlation ranged from .803-.932. In light of the large genomic overlap between the two, combining genomic information associated with pathological and non-pathological substance use phenotypes has the potential to boost power for substance-related research questions. For example, it is possible that combining such pathological and non-pathologic genetic information is a more powerful approach to predicting substance use disorder liability. Additionally, interrogating the *non*-overlapping genetic characteristics of these two factors—as well as that of their constituent traits--could provide illuminating insights into their shared and independent associations with psychopathological, psychological, disinhibitory, personality, and medical outcomes. For example, genomic SUB variance not implicated in SUD may be less likely to include genomic loci that are associated with negative side-effects upon cessation of substances. Somewhat less intuitively: even though substance use is a prerequisite for substance use disorder, the approach used in the present study and in similar designs allows for the dissociation of genetic prediction of pathological substance use outcomes from that of non-pathological substance use outcomes. Evidence has suggested that substance use disorders tend to be more positively genetically associated with certain psychopathological traits when compared to externalizing (Poore et al., 2023) and substance consumption (Gelernter and Polimanti, 2021; Mallard et al., 2022; Sanchez-Roige et al., 2019b). Though it is currently unclear to what extent the sampling procedures for SUD GWAS—which draw many cases from psychiatric populations--may be contributing to this finding (Poore et al., 2023), loci implicated in SUD but not SUB could point to a critical subset of pleiotropic effects implicated in psychopathologies. Further exploration of this independent signal may provide valuable context for attempts to identify treatments and evaluate sources of psychiatric or medical comorbidity. Additionally, it will be important to examine non-biological phenotypes such as socioeconomic status and clinician bias in further elucidating which substance users are most likely to be diagnosed with a substance use disorder.

Research has shown that SUD and externalizing are highly genetically correlated at the common latent factor level and that putatively causal SNPs of their constituent summary statistics are largely overlapping (Poore et al., 2023). The models we tested additionally revealed that there may be important specific covariance between BD and SU/SUD not shared between all three factors. Though this independent covariance only accounted for one fifth of the total genetic covariance between the residual higher-order factors at most, it was nonetheless non-negligible in both of the main models. Though this covariance only explained a small proportion of total BD variance in our models, the relationship between this residual genetic overlap and prediction of outcomes outside of an externalizing framework is worthy of further study.

Finally, evidence in the literature (Brick et al., 2023; Poore et al., 2023) suggests there is a genome-wide/polygenic signal associated with externalizing and BD independent of substance use *and* substance use disorder, further indicating that there could be significant genomic correlates specific to the current study’s construction of BD. While we evaluated BD using a combination of psychiatric and non-psychiatric traits--age at first sex (FSEX, reverse-coded), general risk tolerance (RISK), number of lifetime sexual partners (NSEX), and attention-deficit/hyperactivity disorder (ADHD)--all four traits relate to reward-seeking (i.e., sexual behaviors) and/or impulsivity (i.e., certain hyperactive characteristics in ADHD). While these mechanisms are relevant to substance use and substance use disorder as well, additional domains such as attentional processes (i.e., in ADHD) and subjective opinions of one’s own disinhibitory tendencies (i.e., for self-reported risk tolerance) are more specifically characteristic of BD. Given the substantial proportion of BD-specific residual variance we calculated, additional investigation into this construct’s genomic architecture could uncover an alternative molecular context for understanding risk-taking, reward-seeking, and sensation-seeking over and above its direct common pathways with substance use and/or substance use disorder.

### 4.3 Limitations

It is important to note that there are limitations to consider in the interpretation of our findings. Firstly, the samples on which we based our analyses were not representative of the general human population. All of our samples came from individuals of European ancestry, and many of the participants included were taken from the UK Biobank, which is an older-skewing British sample, while Opioid Use Disorder and Problem Alcohol Use utilized the Million Veterans Program sample, which is comprised entirely of U.S. veterans, is only 9% female, and skews towards older participants (U.S. Department of Veterans Affairs 2024). Some cohorts also utilized a combination of childhood and adult data (i.e., in the ADHD GWAS) (Demontis et al., 2023).

While our study provided important context for understanding the genetic architecture of externalizing at the genome-wide common SNP level, a more complete picture of substance use, substance use disorders, and reward-seeking behaviors, as well as the relationships between these constructs, will require evaluation of environment effects and non-genomic biological phenomena--which GWAS do not directly measure--as well as potential effects of gene-environment interactions. Factors such as racial/ethnic disparities in access to analgesic drugs (Samuel et al., 2019) and consequences of trauma on externalizing and substance use behaviors would add vital context to a broader understanding of the phenomena considered in the current study. In addition, we did not include specific single nucleotide polymorphism (SNP) effects in our model, and so we did not directly identify specific genes or biological pathways associated with the constructs we measured.

Because our models’ latent factors were based on a small number of indicators, the GWAS we included in our models were unlikely to exhaustively capture the genetic components associated with the focal constructs. Because very large sample sizes are required for powerful genome-wide analysis of complex traits, smaller sample sizes—in addition to less precise heritability estimates--for traits such as opioid use disorder (OUD), polysubstance use (DRUG), and problem tobacco use (PTU) may have biased results due to being underpowered. Larger sample sizes and GWAS of more variable substance use outcomes (i.e., recreational opioid use, polysubstance addiction) will be instructive to portraying a clearer picture of the genetic architecture(s) of substance phenotypes.

### 4.4 Conclusion

Substance use disorder (SUD), substance use (SUB), and behavioral disinhibition (BD) represent three related constructs that have been subjects of common interest in psychological and genetic research. Using hierarchical Cholesky decomposition in Genomic SEM, this study attempted to isolate the extent to which genetic variability underlying BD exists beyond shared variance with the prior two constructs and examined the nature of the genetic covariation between all three constructs.

BD was highly correlated with SUD and SUB, the latter two of which showed some evidence of being more correlated with one another still. More than half of the variation in BD could be explained by a broad higher-order factor absorbing variation across all three constructs, while, depending on the model, between none and a small fraction of the variation in BD operated via an independent pathway with SUD or SUB outside of this broad effect. Moreover, a significant minority of residual BD variability remained after partialing out covariance with *both* SUD and SUB, indicating that there is sizable genomic BD variation that does not overlap with substance use or substance use disorder. Future research could demonstrate the utility of boosting power through combining data across pathological and non-pathological substance use indicators though could also shed interesting light on the differences in genetic and molecular correlates of SUD vs. SUB, including outside of direct implications for behavioral disinhibition/externalizing. Extending these lines of inquiry is likely to yield important insights into genetic mechanisms linked to reward response, drug metabolization, risk-taking, and psychopathology.

## Acknowledgements

We thank Alexander S. Hatoum for helpful feedback during the analytic stages of this project. The Substance Use Disorders Working Group of the Psychiatric Genomics Consortium (PGC-SUD) is supported by funds from NIDA and NIMH to MH109532. We gratefully acknowledge their contributing studies and the participants in those studies without whom this effort would not be possible. The MVP summary statistics were obtained via an approved dbGaP application (phs001672.v9.p1). See https://www.research.va.gov/mvp/ and Gaziano, J.M. et al. Million Veteran Program: A mega-biobank to study genetic influences on health and disease. J Clin Epidemiol 70, 214-23 (2016) for more details. This research is based on data from the Million Veteran Program, Office of Research and Development, Veterans Health Administration, and was supported by the Veterans Administration (VA) Million Veteran Program (MVP) award #000. The authors thank Million Veteran Program (MVP) staff, researchers, and volunteers who have contributed to MVP, and especially participants who generously agreed to enroll in the study. This study included summary statistics of a genetic study on cannabis use (Pasman et al., 2018, Nature Neuroscience). We would like to acknowledge all participating groups of the International Cannabis Consortium, and in particular the members of the working group including Joelle Pasman, Karin Verweij, Nathan Gillespie, Eske Derks, and Jacqueline Vink. Pasman et al. (2018) included data from the UK Biobank resource under application numbers 9905, 16406 and 25331. We also thank the research participants and employees of 23andMe, Inc. for making this work possible.

## Author Contributions

**TBH:** Conceptualization, Methodology, Software, Validation, Formal analysis, Resources, Data Curation, Writing – Original Draft, Visualization, Project administration **KZ:** Methodology, Writing – Review & Editing, Visualization **DEG:** Software, Writing – Review & Editing, Supervision **ADG:** Methodology, Software, Writing – Review & Editing, Supervision **MCS:** Conceptualization, Methodology, Resources, Writing – Review & Editing, Supervision, Project administration, Funding acquisition

## Data Availability Statement

Links to available (unrestricted) summary statistics are provided in Table 1.

## Declarations

### Funding

This research was supported by the National Institute on Drug Abuse T32 Training Grant DA017637 (TBH; KZ) and R01 Grant DA053693 (MCS; DEG), as well as by the National Institute of Mental Health R01 Grant MH120219 and the National Institute of Aging RF1 Grant MH120219 (ADG). The funding sources had no involvement in the study design, in the collection, analysis, and interpretation of data, in the writing of the report, or in the decision to submit the article for publication.

### Conflict of interest

The authors declare that they have no conflict of interest.

---

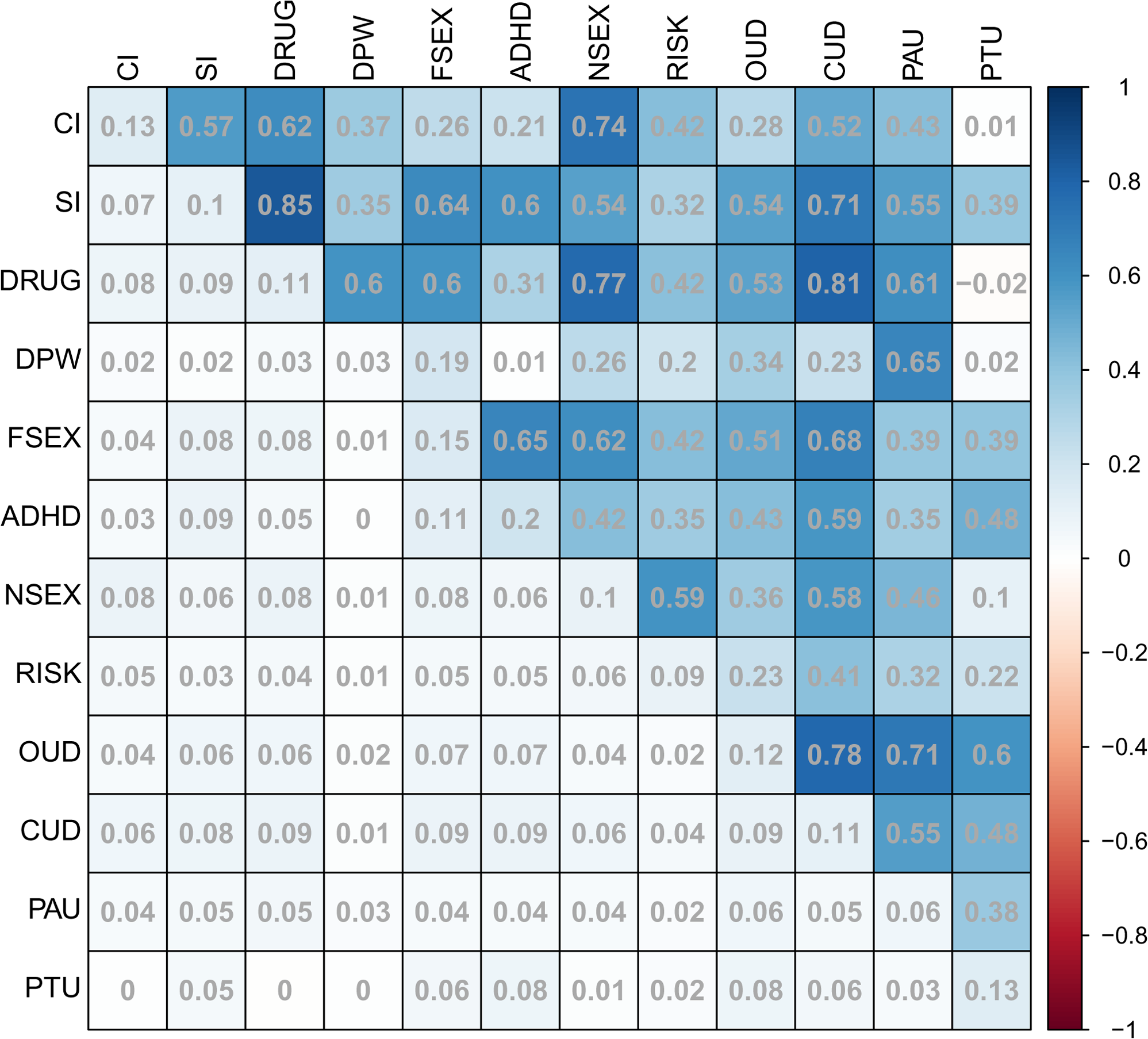

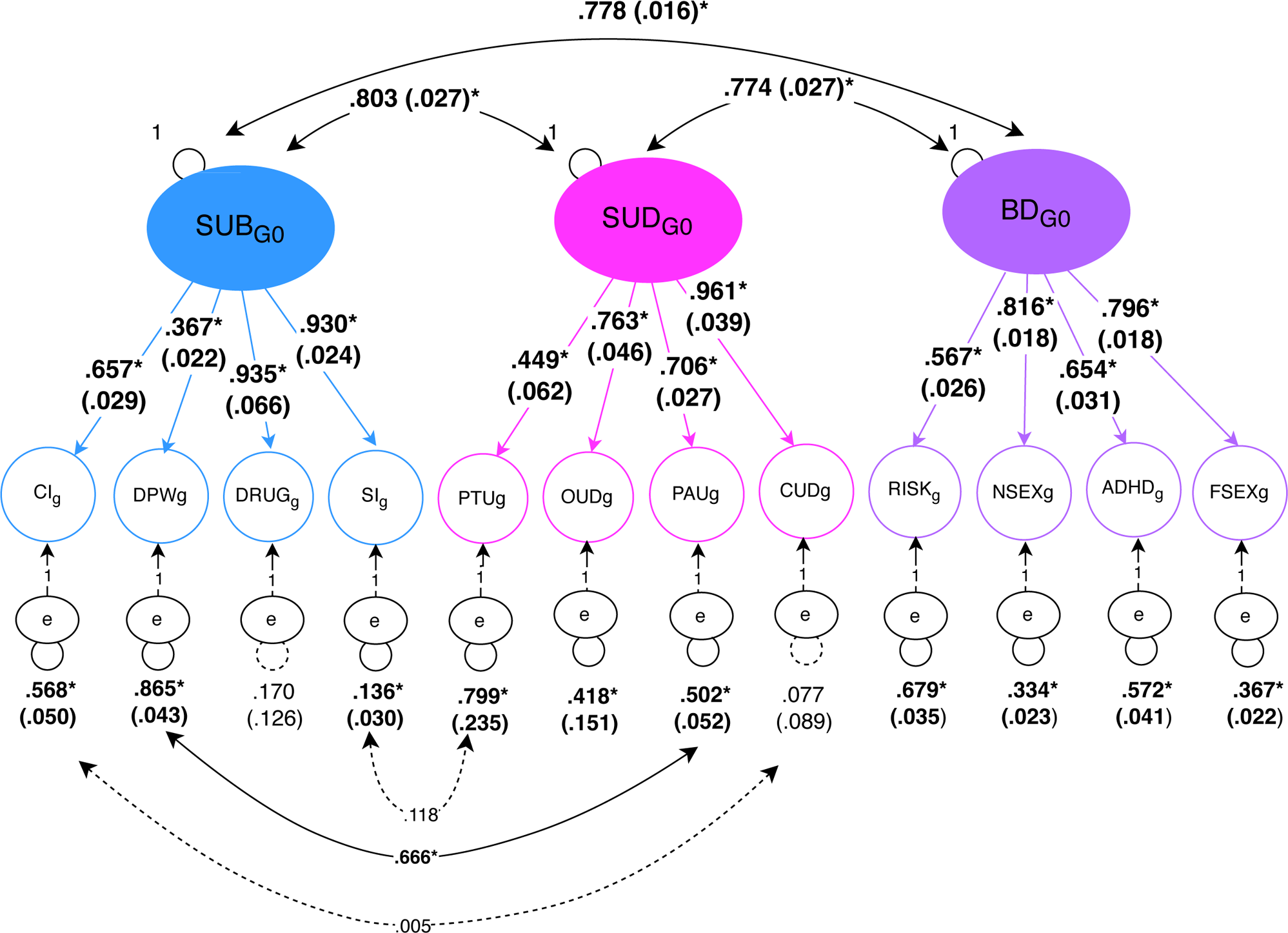

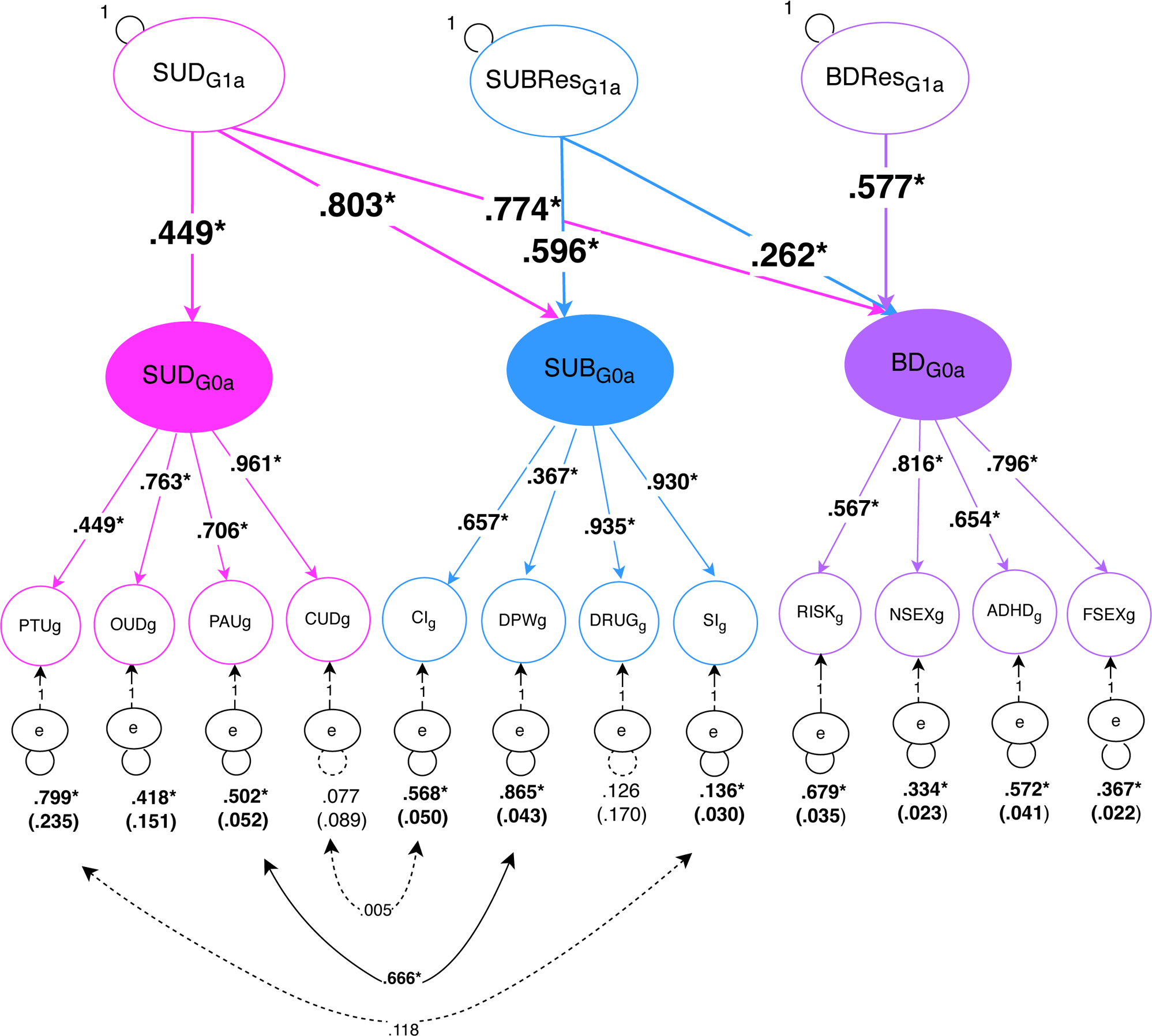

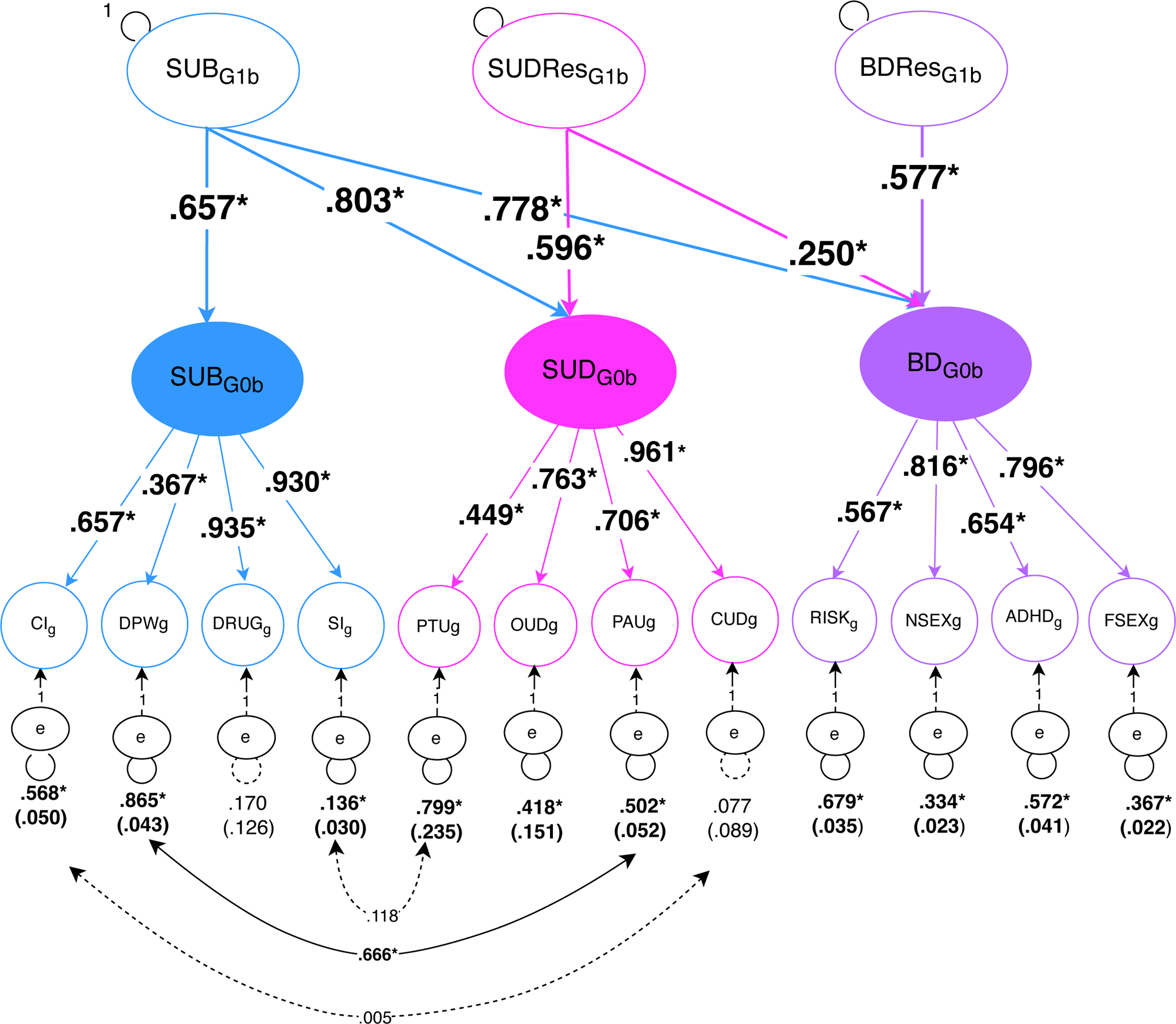

## Notes

### Competing Interest Statement

The authors have declared no competing interest.

### Clinical Protocols

https://osf.io/fjy5h

### Author Declarations

Summary statistics from the Million Veterans Program were obtained via an approved dbGAP application, which did not require IRB approval. The 23andMe summary data, which were based on human data that 23andMe collected under an IRB-approved protocol in accordance with US federal guidelines for "Protection of Human Subjects," were obtained via a data transfer agreement between the University of Colorado Boulder and 23andMe. All other data that this project used were publicly available or were obtained from the researcher(s) who produced the summary results.

