## Supplementary material for "Partitioning the Genomic Components of Behavioral Disinhibition and Substance Use (Disorder) Using Genomic Structural Equation Modeling": Figure Captions

**Main Figures**

**Figure 1. The genetic covariance/correlation matrix (otherwise known as the S Matrix) depicting unstandardized genetic covariances (in the lower left triangle) and genetic correlations (in the upper right triangle) for the twelve externalizing traits used in the final models.** Diagonals show the SNP (single nucleotide polymorphisms)-based heritabilities. CI = Cannabis Initiation, SI = Smoking Initiation, DRUG = Drug Experimentation (the number of different classes of drugs an individual has used out of eleven), DPW = Drinks Per Week, FSEX = Age at First Sexual Intercourse (reverse-coded), ADHD = Attention-deficit Hyperactivity Disorder, NSEX = Number of Lifetime Sexual Partners, RISK = General Risk Tolerance; OUD = Opioid Use Disorder, CUD = Cannabis Use Disorder, PAU = Problem Alcohol Use, PTU = Problem Tobacco Use.

**Figure 2.** **A standardized correlated factors depiction of the three lower-order externalizing common factors depicted in the measurement models.** In order to extract influences of within-substance intercorrelations, the residuals of PTU and SI, CUD and CI, and PAU and DPW are correlated across the SUB and SUD factors. SUB_G0_ is the lower-order Substance Use factor, SUD_G0_ is the lower-order Substance Use Disorder factor, and BD_G0_ is the lower-order Behavioral Disinhibition factor. Each factor describes four indicators, the latter of which are defined by the summary results from genome-wide association studies for the trait in question. The *g* subscripts denote genomic variance. In addition to SUB_G0_, SUD_G0_, and BD_G0_, the single trait indicators are represented as latent factors to demonstrate that their genomic properties are not directly observed. CI = Cannabis Initiation, SI = Smoking Initiation, DRUG = Drug Experimentation (the number of different classes of drugs an individual has used out of eleven), DPW = Drinks Per Week, FSEX = Age at First Sexual Intercourse (reverse-coded), ADHD = Attention-deficit Hyperactivity Disorder, NSEX = Number of Lifetime Sexual Partners, RISK = General Risk Tolerance; OUD = Opioid Use Disorder, CUD = Cannabis Use Disorder, PAU = Problem Alcohol Use, PTU = Problem Tobacco Use. **p* < .05. Standard errors are in parentheses, and dotted paths denote non-significance.

**Figure 3. The Two Main Cholesky Models.** Hierarchical trivariate Cholesky decompositions integrating the lower-order factors from the measurement models. In order to extract influences of within-substance intercorrelations, the residuals of PTU and SI, CUD and CI, and PAU and DPW are correlated across the SUB and SUD factors. In these models, only the higher-order factors are explicitly standardized, while loadings are standardized with respect to the full model, including endogenous latent factors. CI = Cannabis Initiation, DPW = Drinks per Week, DRUG = Drug Experimentation, SI = Smoking Initiation, PTU = Problem Tobacco Use, OUD = Opioid Use Disorder, PAU = Problem Alcohol Use, CUD = Cannabis Use Disorder, RISK = Risk Tolerance, NSEX = Number of Lifetime Sexual Partners, ADHD = Attention Deficit/Hyperactivity Disorder, FSEX = Age at First Sexual Intercourse (reverse-coded). All measures, including single trait indicators, are represented as latent factors to demonstrate that their genomic properties are not directly observed. **p* < .05. Standard errors are in parentheses, and dotted paths denote non-significance. **A)** In Model 1, the higher-order left-most factor SUD_G1a_ describes all of the variance in the lower-order factor SUD_G0a_ (Substance Use Disorder) while also describing some of the variance in the lower-order factors SUB_G0a_ (Substance Use) and BD_G0a_ (Behavioral Disinhibition). SUBRes_G1a_ is an orthogonal second-position higher-order factor which captures the remaining variance in SUB_G0a_ and on which BD_G0a_ also loads. Finally, BDRes_G1a_ explains the residual variance in BD not yet accounted for in the model. **B)** In Model 2, the left-most factor SUB_G1b_ describes all of the variance in the lower-order factor SUB_G0b_ (Substance Use) while also describing some of the variance in the lower-order factors SUD_G0b_ (Substance Use Disorder) and BD_G0b_ (Behavioral Disinhibition). SUDRes_G1b_ is an orthogonal second-position higher-order factor which captures the remaining variance in SUD_G0b_ and on which BD_G0b_ also loads.

**Supplementary Figures**

**Supplementary Figure 1. The genetic covariance/correlation matrix (otherwise known as the S Matrix) depicting unstandardized genetic covariances (in the lower left triangle) and genetic correlations (in the upper right triangle) between the fifteen externalizing traits initially explored for the main models.** Diagonals show the SNP (single nucleotide polymorphisms)-based heritabilities. CI = Cannabis Initiation, CPD = Cigarettes per Day, SI = Smoking Initiation, FREQ = Drinking Frequency, DRINK.STAT = Drinking Status, DRUG = Drug Experimentation (the number of different classes of drugs an individual has used out of eleven), DPW = Drinks Per Week, FSEX = Age at First Sexual Intercourse (reverse-coded), ADHD = Attention-deficit Hyperactivity Disorder, NSEX = Number of Lifetime Sexual Partners, RISK = General Risk Tolerance; OUD = Opioid Use Disorder, CUD = Cannabis Use Disorder, PAU = Problem Alcohol Use, PTU = Problem Tobacco Use.

**Supplementary Figure 2. Measurement models**. Each factor describes four indicators, which are defined by the summary results from genome-wide association studies for the trait in question. The *g* subscripts denote genomic variance. In addition to BD, SUD, and SUB, the single trait indicators are represented as latent factors to demonstrate that their genomic properties are not directly observed. **p* < .05. Standard errors are in parentheses, and dotted paths denote non-significance. **(A)** BD, the Behavioral Disinhibition factor, FSEX = Age at First Sexual Intercourse (reverse-coded), ADHD = Attention-deficit Hyperactivity Disorder, NSEX = Number of Lifetime Sexual Partners, RISK = General Risk Tolerance; **(B)** SUD, the Substance Use Disorder factor, OUD = Opioid Use Disorder, CUD = Cannabis Use Disorder, PAU = Problem Alcohol Use, PTU = Problem Tobacco Use; **(C)** SUB, the Substance Use factor, CI = Cannabis Initiation, SI = Smoking Initiation, DRUG = Drug Experimentation (the number of different classes of drugs an individual has used out of eleven), DPW = Drinks Per Week.

**Supplementary Figure 3**. **A standardized post-hoc correlated factors depiction of the lower-order BD factor, along with the modified SUD and SUB common factors, the latter of which drop the OUD (opioid use disorder) and DRUG (polysubstance use) indicators, respectively.** In order to extract influences of within-substance intercorrelations, the residuals of PTU and SI, CUD and CI, and PAU and DPW are correlated across the MOD_SUB and MOD_SUD factors. MOD_SUB_G0_ is the three-indicator modified lower-order Substance Use factor, while MOD_SUD_G0_ is the three-indicator modified lower-order Substance Use Disorder factor, which collectively include only cannabis, alcohol, and nicotine phenotypes. BD_G0_ is the lower-order Behavioral Disinhibition factor and is unmodified from the previous models. Each factor describes a set of indicators, the latter of which are defined by the summary results from genome-wide association studies for the trait in question. The *g* subscripts denote genomic variance. In addition to MOD_SUB_G0_, MOD_SUD_G0_, and BD_G0_, the single trait indicators are represented as latent factors to demonstrate that their genomic properties are not directly observed. CI = Cannabis Initiation, SI = Smoking Initiation, DPW = Drinks Per Week; FSEX = Age at First Sexual Intercourse (reverse-coded), ADHD = Attention-deficit Hyperactivity Disorder, NSEX = Number of Lifetime Sexual Partners, RISK = General Risk Tolerance; CUD = Cannabis Use Disorder, PAU = Problem Alcohol Use; PTU = Problem Tobacco Use. **p* < .05. Standard errors are in parentheses.

**Supplementary Figure 4. The Two Cholesky Models with the modified SUD and SUB common factors.** In order to extract influences of within-substance intercorrelations, the residuals of PTU and SI, CUD and CI, and PAU and DPW are correlated across the MOD_SUB and MOD_SUD factors. In these models, only the higher-order factors are explicitly standardized, while loadings are standardized with respect to the full model, including endogenous latent factors. The *g* subscripts denote genomic variance. CI = Cannabis Initiation, SI = Smoking Initiation, DPW = Drinks Per Week; FSEX = Age at First Sexual Intercourse (reverse-coded), ADHD = Attention-deficit Hyperactivity Disorder, NSEX = Number of Lifetime Sexual Partners, RISK = General Risk Tolerance; CUD = Cannabis Use Disorder, PAU = Problem Alcohol Use, PTU = Problem Tobacco Use. All measures, including single trait indicators, are represented as latent factors to demonstrate that their genomic properties are not directly observed. **p* < .05. Standard errors are in parentheses, and dotted paths denote non-significance. **A)** In Modified Model 1, the higher-order left-most factor MOD_SUD_G1c_ describes all of the variance in the lower-order factor MOD_SUD_G0c_ (Modified Substance Use Disorder) while also describing some of the variation in the lower-order factors MOD_SUB_G0c_ (Modified Substance Use) and BD_G0c_ (Behavioral Disinhibition). MOD_SUBRes_G1c_ is an orthogonal second-position higher-order factor which captures the remaining variance in MOD_SUB_G0c_ and on which BD_G0c_ also loads. Finally, MOD_BDRes_G1c_ explains the residual variance in BD_G0c_ not yet accounted for in the model. **B)** In Modified Model 2, the higher-order left-most factor MOD_SUB_G1d_ describes all of the variance in the lower-order factor MOD_SUB_G0d_ (Modified Substance Use) while also describing some of the variation in the lower-order factors MOD_SUD_G0d_ (Modified Substance Use Disorder) and BD_G0d_ (Behavioral Disinhibition). MOD_SUDRes_G1d_ is an orthogonal second-position higher-order factor which captures the remaining variance in MOD_SUD_G0d_ and on which BD_G0d_ also loads. Finally, MOD_BDRes_G1d_ explains the residual variance in BD_G0d_ not yet accounted for in the model.

**Supplementary Figure 5. A standardized correlated factors depiction of the three lower-order externalizing common factors depicted in the measurement models, with no residual correlations between the SUB and SUD indicators.** SUB_G0_ is the lower-order Substance Use factor, SUD_G0_ is the lower-order Substance Use Disorder factor, and BD_G0_ is the lower-order Behavioral Disinhibition factor. Each factor describes four indicators, which are defined by the summary results from genome-wide association studies for the trait in question. The *g* subscripts denote genomic variance. In addition to SUB_G0_, SUD_G0_, and BD_G0_, the single trait indicators are represented as latent factors to demonstrate that their genomic properties are not directly observed. CI = Cannabis Initiation, SI = Smoking Initiation, DRUG = Drug Experimentation (the number of different classes of drugs an individual has used out of eleven), DPW = Drinks Per Week, FSEX = Age at First Sexual Intercourse (reverse-coded), ADHD = Attention-deficit Hyperactivity Disorder, NSEX = Number of Lifetime Sexual Partners, RISK = General Risk Tolerance; OUD = Opioid Use Disorder, CUD = Cannabis Use Disorder, PAU = Problem Alcohol Use, PTU = Problem Tobacco Use. **p* < .05. Standard errors are in parentheses, and dotted paths denote non-significance.

**Supplementary Figure 6. A standardized post-hoc correlated factors depiction of the lower-order BD factor, along with the modified SUD and SUB common factors, with no residual correlations between the SUB and SUD indicators.** MOD_SUB_G0_ is the three-indicator modified lower-order Substance Use factor, while MOD_SUD_G0_ is the three-indicator modified lower-order Substance Use Disorder factor, which collectively include only cannabis, alcohol, and nicotine phenotypes. BD_G0_ is the lower-order Behavioral Disinhibition factor and is unmodified from the previous models. Each factor describes a set of indicators, the latter of which are defined by the summary results from genome-wide association studies for the trait in question. The *g* subscripts denote genomic variance. In addition to MOD_SUB_G0_, MOD_SUD_G0_, and BD_G0_, the single trait indicators are represented as latent factors to demonstrate that their genomic properties are not directly observed. CI = Cannabis Initiation, SI = Smoking Initiation, DPW = Drinks Per Week; FSEX = Age at First Sexual Intercourse (reverse-coded), ADHD = Attention-deficit Hyperactivity Disorder, NSEX = Number of Lifetime Sexual Partners, RISK = General Risk Tolerance; CUD = Cannabis Use Disorder, PAU = Problem Alcohol Use; PTU = Problem Tobacco Use. **p* < .05. Standard errors are in parentheses.

**Supplementary Figure 7. The Two Main Cholesky Models, with no residual correlations between the SUB and SUD indicators.** In these models, only the higher-order factors are explicitly standardized, while loadings are standardized with respect to the full model, including endogenous latent factors. CI = Cannabis Initiation, DPW = Drinks per Week, DRUG = Drug Experimentation, SI = Smoking Initiation, PTU = Problem Tobacco Use, OUD = Opioid Use Disorder, PAU = Problem Alcohol Use, CUD = Cannabis Use Disorder, RISK = Risk Tolerance, NSEX = Number of Lifetime Sexual Partners, ADHD = Attention Deficit/Hyperactivity Disorder, FSEX = Age at First Sexual Intercourse (reverse-coded). All measures, including single trait indicators, are represented as latent factors to demonstrate that their genomic properties are not directly observed. **p* < .05. Standard errors are in parentheses, and dotted paths denote non-significance. **A)** In Model 1, the higher-order left-most factor SUD_G1a_ describes all of the variance in the lower-order factor SUD_G0a_ (Substance Use Disorder) while also describing some of the variance in the lower-order factors SUB_G0a_ (Substance Use) and BD_G0a_ (Behavioral Disinhibition). SUBRes_G1a_ is an orthogonal second-position higher-order factor which captures the remaining variance in SUB_G0a_ and on which BD_G0a_ also loads. Finally, BDRes_G1a_ explains the residual variance in BD not yet accounted for in the model. **B)** In Model 2, the left-most factor SUB_G1b_ describes all of the variance in the lower-order factor SUB_G0b_ (Substance Use) while also describing some of the variance in the lower-order factors SUD_G0b_ (Substance Use Disorder) and BD_G0b_ (Behavioral Disinhibition). SUDRes_G1b_ is an orthogonal second-position higher-order factor which captures the remaining variance in SUD_G0b_ and on which BD_G0b_ also loads. Finally, BDRes_G1b_ explains the residual variance in BD not yet accounted for in the model.

**Supplementary Figure 8. The Two Cholesky Models with the modified SUD and SUB common factors, with no residual correlations between the SUB and SUD indicators.** In these models, only the higher-order factors are explicitly standardized, while loadings are standardized with respect to the full model, including endogenous latent factors. The *g* subscripts denote genomic variance. CI = Cannabis Initiation, SI = Smoking Initiation, DPW = Drinks Per Week; FSEX = Age at First Sexual Intercourse (reverse-coded), ADHD = Attention-deficit Hyperactivity Disorder, NSEX = Number of Lifetime Sexual Partners, RISK = General Risk Tolerance; CUD = Cannabis Use Disorder, PAU = Problem Alcohol Use, PTU = Problem Tobacco Use. All measures, including single trait indicators, are represented as latent factors to demonstrate that their genomic properties are not directly observed. **p* < .05. Standard errors are in parentheses, and dotted paths denote non-significance. **A)** In Modified Model 1, the higher-order left-most factor MOD_SUD_G1c_ describes all of the variance in the lower-order factor MOD_SUD_G0c_ (Modified Substance Use Disorder) while also describing some of the variation in the lower-order factors MOD_SUB_G0c_ (Modified Substance Use) and BD_G0c_ (Behavioral Disinhibition). MOD_SUBRes_G1c_ is an orthogonal second-position higher-order factor which captures the remaining variance in MOD_SUB_G0c_ and on which BD_G0c_ also loads. Finally, MOD_BDRes_G1c_ explains the residual variance in BD_G0c_ not yet accounted for in the model. **B)** In Modified Model 2, the higher-order left-most factor MOD_SUB_G1d_ describes all of the variance in the lower-order factor MOD_SUB_G0d_ (Modified Substance Use) while also describing some of the variation in the lower-order factors MOD_SUD_G0d_ (Modified Substance Use Disorder) and BD_G0d_ (Behavioral Disinhibition). MOD_SUDRes_G1d_ is an orthogonal second-position higher-order factor which captures the remaining variance in MOD_SUD_G0d_ and on which BD_G0d_ also loads. Finally, MOD_BDRes_G1d_ explains the residual variance in BD_G0d_ not yet accounted for in the model.
