## Supplementary Note for "Partitioning the Genomic Components of Behavioral Disinhibition and Substance Use (Disorder) Using Genomic Structural Equation Modeling"

For the lower-order SUB (Substance Use) factor, we initially explored seven non-pathological indicators related to substance use: cannabis initiation (Pasman et al., 2018), smoking initiation (Liu et al., 2019; Saunders et al., 2022), cigarettes per day (in current or former smokers) (Liu et al., 2019; Saunders et al., 2022), drinks per week (Liu et al., 2019; Saunders et al., 2022), drinking frequency (Colbert et al., 2021), drinking status (drinker/non-drinker) (Liu et al., 2019), and drug experimentation (the number of different classes of drugs an individual has used out of eleven) (Sanchez-Roige et al., 2019) . In line with the trivariate structure of the proposed models, we explored only single-factor solutions for SUB. Thus, contrary to our pre-registration (<https://osf.io/fjy5h>), in which we stated that we would use an exploratory factor analysis function, we instead began by using the usermodel() function in Genomic SEM (Grotzinger et al., 2019) to conduct a common factor analysis that utilized the genetic covariances between the relevant munged GWAS summary statistics. Otherwise, we proceeded in line with the criteria for our SUB factor as laid out in our preregistered plan, in which we proposed to: 1) omit traits if their standardized loadings were <.20; 2) achieve a factor with a comparative fit index of .85 or higher; 3) include at least three classes or categories of substances. Supplementary Figure 1 shows the genetic covariance/correlation matrix depicting unstandardized genetic covariances (in the lower left triangle) and genetic correlations (in the upper right triangle) between the eight indicators used for the Behavioral Disinhibition and Substance Use Disorder factors (see Supplementary Figures 2A-2B) and the seven pre-registered substance use traits.

The first iteration of our SUB factor (Model 0A; Supplementary Table S3), on which all seven pre-registered substance use traits (see Table 1) loaded, produced a negative standardized residual variance (-0.025) for drinks per week (DPW). Standardized loadings were considerably greater than .20 for every trait except cigarettes per day. This was not entirely surprising given that cigarettes per day has been reported as having negative or ~0 correlations with other substance use traits such as cannabis use and DPW(Abdellaoui and Verweij, 2021; Saunders et al., 2022). For Model 0B, in which we constrained the DPW variance term to be positive, the standardized loading for cigarettes per day remained negative (-0.126; see Supplementary Table S4). Thus, we omitted cigarettes per day in Model 0C, which showed the highest standardized loadings for DPW (1.007, S.E. = .028), Drinking Frequency (.811, S.E. = .030), Drug Experimentation (.680, S.E. = .065), and Drinking Status (.665, S.E = .030). Cannabis Initiation (CI) and Smoking Initiation (SI) loadings were smaller (.482, S.E. = .031 and .332, S.E. = .021, respectively) but still above our preregistered threshold (see Supplementary Table S5). Unfortunately, Model 0C was associated with a CFI of .815 (SRMR = .196, AIC = 1036.288), indicating relatively poor fit that did not satisfy our preregistered criteria. In Model 0D, we eliminated the Drinking Status variable because it was one of three drinking variables remaining in the factor (while smoking, cannabis, and polysubstance use were only represented by one trait each in this model) and because the former correlated at -.015 with SI. The factor loadings for DPW and Drinking Frequency in Model 0C were notably high when compared to some of the other loadings, suggesting that alcohol traits may have been conferring disproportionate influence on the common factor, particularly since the remaining three indicators all represented different substance classes. Because DPW showed a slightly larger correlation with all of the remaining SUB indicator than did Drinking Frequency, we retained the former and dropped the latter. Model 0D showed much improved fit (CFI = .993, SRMR = .054, AIC = 26.850) when compared to 0C but yielded a negative standardized residual variance (-.148) for Drug Experimentation (DRUG) (Supplementary Table S6), so we constrained this residual variance to be positive. Model 0E (see Supplementary Table S7), which, like the Behavioral Disinhibition and Substance Use Disorder factors (Supplementary Figures 2A-2B) included four traits—DRUG, CI, SI, and DPW (see Supplementary Figure 2C)--had good model fit (CFI = .994, SRMR = .063, AIC = 26.145). The factor also captured more than three substance categories, and all standardized factor loadings were .483 or greater. Though the standardized loading for DRUG was greater than one, the corresponding error term was not negative and the standardized loading dropped below one after SUB was integrated into the correlated factors model and the trivariate decompositions (see Figures 2-3), so model 0E was retained as the final SUB factor.

Liu, M., Jiang, Y., Wedow, R., Li, Y., Brazel, D.M., Chen, F., Datta, G., Davila-Velderrain, J., McGuire, D., Tian, C., Zhan, X., Choquet, H., Docherty, A.R., Faul, J.D., Foerster, J.R., Fritsche, L.G., Gabrielsen, M.E., Gordon, S.D., Haessler, J., Hottenga, J.-J., Huang, H., Jang, S.-K., Jansen, P.R., Ling, Y., Mägi, R., Matoba, N., McMahon, G., Mulas, A., Orrù, V., Palviainen, T., Pandit, A., Reginsson, G.W., Skogholt, A.H., Smith, J.A., Taylor, A.E., Turman, C., Willemsen, G., Young, H., Young, K.A., Zajac, G.J.M., Zhao, W., Zhou, W., Bjornsdottir, G., Boardman, J.D., Boehnke, M., Boomsma, D.I., Chen, C., Cucca, F., Davies, G.E., Eaton, C.B., Ehringer, M.A., Esko, T., Fiorillo, E., Gillespie, N.A., Gudbjartsson, D.F., Haller, T., Harris, K.M., Heath, A.C., Hewitt, J.K., Hickie, I.B., Hokanson, J.E., Hopfer, C.J., Hunter, D.J., Iacono, W.G., Johnson, E.O., Kamatani, Y., Kardia, S.L.R., Keller, M.C., Kellis, M., Kooperberg, C., Kraft, P., Krauter, K.S., Laakso, M., Lind, P.A., Loukola, A., Lutz, S.M., Madden, P.A.F., Martin, N.G., McGue, M., McQueen, M.B., Medland, S.E., Metspalu, A., Mohlke, K.L., Nielsen, J.B., Okada, Y., Peters, U., Polderman, T.J.C., Posthuma, D., Reiner, A.P., Rice, J.P., Rimm, E., Rose, R.J., Runarsdottir, V., Stallings, M.C., Stančáková, A., Stefansson, H., Thai, K.K., Tindle, H.A., Tyrfingsson, T., Wall, T.L., Weir, D.R., Weisner, C., Whitfield, J.B., Winsvold, B.S., Yin, J., Zuccolo, L., Bierut, L.J., Hveem, K., Lee, J.J., Munafò, M.R., Saccone, N.L., Willer, C.J., Cornelis, M.C., David, S.P., Hinds, D.A., Jorgenson, E., Kaprio, J., Stitzel, J.A., Stefansson, K., Thorgeirsson, T.E., Abecasis, G., Liu, D.J., Vrieze, S., 2019. Association studies of up to 1.2 million individuals yield new insights into the genetic etiology of tobacco and alcohol use. Nat. Genet. 51, 237–244. https://doi.org/10.1038/s41588-018-0307-5

Pasman, J.A., Verweij, K.J.H., Gerring, Z., Stringer, S., Sanchez-Roige, S., Treur, J.L., Abdellaoui, A., Nivard, M.G., Baselmans, B.M.L., Ong, J.-S., Ip, H.F., van der Zee, M.D., Bartels, M., Day, F.R., Fontanillas, P., Elson, S.L., de Wit, H., Davis, L.K., MacKillop, J., Derringer, J.L., Branje, S.J.T., Hartman, C.A., Heath, A.C., van Lier, P.A.C., Madden, P.A.F., Mägi, R., Meeus, W., Montgomery, G.W., Oldehinkel, A.J., Pausova, Z., Ramos-Quiroga, J.A., Paus, T., Ribases, M., Kaprio, J., Boks, M.P.M., Bell, J.T., Spector, T.D., Gelernter, J., Boomsma, D.I., Martin, N.G., MacGregor, S., Perry, J.R.B., Palmer, A.A., Posthuma, D., Munafò, M.R., Gillespie, N.A., Derks, E.M., Vink, J.M., 2018. GWAS of lifetime cannabis use reveals new risk loci, genetic overlap with psychiatric traits, and a causal effect of schizophrenia liability. Nat. Neurosci. 21, 1161–1170. https://doi.org/10.1038/s41593-018-0206-1

Sanchez-Roige, S., Fontanillas, P., Elson, S.L., Gray, J.C., de Wit, H., MacKillop, J., Palmer, A.A., 2019. Genome-Wide Association Studies of Impulsive Personality Traits (BIS-11 and UPPS-P) and Drug Experimentation in up to 22,861 Adult Research Participants Identify Loci in the CACNA1I and CADM2 genes. J. Neurosci. Off. J. Soc. Neurosci. 39, 2562–2572. https://doi.org/10.1523/JNEUROSCI.2662-18.2019

Saunders, G.R.B., Wang, X., Chen, F., Jang, S.-K., Liu, M., Wang, C., Gao, S., Jiang, Y., Khunsriraksakul, C., Otto, J.M., Addison, C., Akiyama, M., Albert, C.M., Aliev, F., Alonso, A., Arnett, D.K., Ashley-Koch, A.E., Ashrani, A.A., Barnes, K.C., Barr, R.G., Bartz, T.M., Becker, D.M., Bielak, L.F., Benjamin, E.J., Bis, J.C., Bjornsdottir, G., Blangero, J., Bleecker, E.R., Boardman, J.D., Boerwinkle, E., Boomsma, D.I., Boorgula, M.P., Bowden, D.W., Brody, J.A., Cade, B.E., Chasman, D.I., Chavan, S., Chen, Y.-D.I., Chen, Z., Cheng, I., Cho, M.H., Choquet, H., Cole, J.W., Cornelis, M.C., Cucca, F., Curran, J.E., de Andrade, M., Dick, D.M., Docherty, A.R., Duggirala, R., Eaton, C.B., Ehringer, M.A., Esko, T., Faul, J.D., Fernandes Silva, L., Fiorillo, E., Fornage, M., Freedman, B.I., Gabrielsen, M.E., Garrett, M.E., Gharib, S.A., Gieger, C., Gillespie, N., Glahn, D.C., Gordon, S.D., Gu, C.C., Gu, D., Gudbjartsson, D.F., Guo, X., Haessler, J., Hall, M.E., Haller, T., Harris, K.M., He, J., Herd, P., Hewitt, J.K., Hickie, I., Hidalgo, B., Hokanson, J.E., Hopfer, C., Hottenga, J., Hou, L., Huang, H., Hung, Y.-J., Hunter, D.J., Hveem, K., Hwang, S.-J., Hwu, C.-M., Iacono, W., Irvin, M.R., Jee, Y.H., Johnson, E.O., Joo, Y.Y., Jorgenson, E., Justice, A.E., Kamatani, Y., Kaplan, R.C., Kaprio, J., Kardia, S.L.R., Keller, M.C., Kelly, T.N., Kooperberg, C., Korhonen, T., Kraft, P., Krauter, K., Kuusisto, J., Laakso, M., Lasky-Su, J., Lee, W.-J., Lee, J.J., Levy, D., Li, L., Li, K., Li, Y., Lin, K., Lind, P.A., Liu, C., Lloyd-Jones, D.M., Lutz, S.M., Ma, J., Mägi, R., Manichaikul, A., Martin, N.G., Mathur, R., Matoba, N., McArdle, P.F., McGue, M., McQueen, M.B., Medland, S.E., Metspalu, A., Meyers, D.A., Millwood, I.Y., Mitchell, B.D., Mohlke, K.L., Moll, M., Montasser, M.E., Morrison, A.C., Mulas, A., Nielsen, J.B., North, K.E., Oelsner, E.C., Okada, Y., Orrù, V., Palmer, N.D., Palviainen, T., Pandit, A., Park, S.L., Peters, U., Peters, A., Peyser, P.A., Polderman, T.J.C., Rafaels, N., Redline, S., Reed, R.M., Reiner, A.P., Rice, J.P., Rich, S.S., Richmond, N.E., Roan, C., Rotter, J.I., Rueschman, M.N., Runarsdottir, V., Saccone, N.L., Schwartz, D.A., Shadyab, A.H., Shi, J., Shringarpure, S.S., Sicinski, K., Skogholt, A.H., Smith, J.A., Smith, N.L., Sotoodehnia, N., Stallings, M.C., Stefansson, H., Stefansson, K., Stitzel, J.A., Sun, X., Syed, M., Tal-Singer, R., Taylor, A.E., Taylor, K.D., Telen, M.J., Thai, K.K., Tiwari, H., Turman, C., Tyrfingsson, T., Wall, T.L., Walters, R.G., Weir, D.R., Weiss, S.T., White, W.B., Whitfield, J.B., Wiggins, K.L., Willemsen, G., Willer, C.J., Winsvold, B.S., Xu, H., Yanek, L.R., Yin, J., Young, K.L., Young, K.A., Yu, B., Zhao, W., Zhou, W., Zöllner, S., Zuccolo, L., Batini, C., Bergen, A.W., Bierut, L.J., David, S.P., Gagliano Taliun, S.A., Hancock, D.B., Jiang, B., Munafò, M.R., Thorgeirsson, T.E., Liu, D.J., Vrieze, S., 2022. Genetic diversity fuels gene discovery for tobacco and alcohol use. Nature 612, 720–724. https://doi.org/10.1038/s41586-022-05477-4
