## Supplementary Table Captions for "Partitioning the Genomic Components of Behavioral Disinhibition and Substance Use (Disorder) Using Genomic Structural Equation Modeling"

**The parameters associated with the Behavioral Disinhibition measurement model** Unstand Est = unstandardized estimate; SE = standard error; STD_Genotype estimates are standardized with respect to the indicators; STD_All estimates are standardized with respect to the full model (equivalent to STD_Genotype in the current model). BD = the Behavioral Disinhibition factor, FSEX = Age at First Sexual Intercourse (reverse-coded), ADHD = Attention-deficit Hyperactivity Disorder, NSEX = Number of Lifetime Sexual Partners, RISK = General Risk Tolerance. BD is standardized.

1. **The parameters associated with the Substance Use Disorder measurement model** Unstand Est = unstandardized estimate; SE = standard error; STD_Genotype estimates are standardized with respect to the indicators; STD_All estimates are standardized with respect to the full model (equivalent to STD_Genotype in the current model). SUD = the Substance Use Disorder factor, OUD = Opioid Use Disorder, CUD = Cannabis Use Disorder, PAU = Problem Alcohol Use, PTU = Problem Tobacco Use. SUD is standardized.
2. **The parameters associated with Model 0A** Unstand Est = unstandardized estimate; SE = standard error; STD_Genotype estimates are standardized with respect to the indicators; STD_All estimates are standardized with respect to the full model (equivalent to STD_Genotype in the current model). CI = Cannabis Initiation, DPW = Drinks per Week, SI = Smoking Initiation, FREQ = Drinking Frequency, Drink.Stat = Drinking Status, DRUG = Drug Experimentation, CPD = Cigarettes per Day. F1 is standardized.
3. **The parameters associated with Model 0B** Unstand Est = unstandardized estimate; SE = standard error; STD_Genotype estimates are standardized with respect to the indicators; STD_All estimates are standardized with respect to the full model (equivalent to STD_Genotype in the current model). CI = Cannabis Initiation, DPW = Drinks per Week, SI = Smoking Initiation, FREQ = Drinking Frequency, Drink.Stat = Drinking Status, DRUG = Drug Experimentation, CPD = Cigarettes per Day. F1 is standardized.
4. **The parameters associated with Model 0C** Unstand Est = unstandardized estimate; SE = standard error; STD_Genotype estimates are standardized with respect to the indicators; STD_All estimates are standardized with respect to the full model (equivalent to STD_Genotype in the current model). CI = Cannabis Initiation, DPW = Drinks per Week, SI = Smoking Initiation, FREQ = Drinking Frequency, Drink.Stat = Drinking Status, DRUG = Drug Experimentation. F1 is standardized.
5. **The parameters associated with Model 0D** Unstand Est = unstandardized estimate; SE = standard error; STD_Genotype estimates are standardized with respect to the indicators; STD_All estimates are standardized with respect to the full model (equivalent to STD_Genotype in the current model). CI = Cannabis Initiation, DPW = Drinks per Week, SI = Smoking Initiation, DRUG = Drug Experimentation. F1 is standardized.
6. **The parameters associated with Model 0E (The Final SUB Factor)** Unstand Est = unstandardized estimate; SE = standard error; STD_Genotype estimates are standardized with respect to the indicators; STD_All estimates are standardized with respect to the full model (equivalent to STD_Genotype in the current model). CI = Cannabis Initiation, DPW = Drinks per Week, SI = Smoking Initiation, DRUG = Drug Experimentation. F1 is standardized.
7. **The parameters associated with a twelve-indicator common factor solution.** Unstand Est = unstandardized estimate; SE = standard error; STD_Genotype estimates are standardized with respect to the indicators; STD_All estimates are standardized with respect to the full model (equivalent to STD_Genotype in the current model). CI = Cannabis Initiation, DPW = Drinks per Week, DRUG = Drug Experimentation, SI = Smoking Initiation, PTU = Problem Tobacco Use, OUD = Opioid Use Disorder, PAU = Problem Alcohol Use, CUD = Cannabis Use Disorder, RISK = Risk Tolerance, NSEX = Number of Lifetime Sexual Partners, ADHD = Attention Deficit/Hyperactivity Disorder, FSEX = Age at First Sexual Intercourse (reverse-coded). F1 is standardized. The residuals of PTU and SI, CUD and CI, and PAU and DPW are correlated.
8. **The parameters associated with a correlated factors model of the original SUB, SUD, and BD factors, with residuals of PTU and SI, CUD and CI, and PAU and DPW correlated.** Unstand Est = unstandardized estimate; SE = standard error; STD_Genotype estimates are standardized with respect to the indicators; STD_All estimates are standardized with respect to the full model (equivalent to STD_Genotype in the correlated factors model). BD = Behavioral Disinhibition; SUB = Substance Use; SUD = Substance Use Disorder; CI = Cannabis Initiation, DPW = Drinks per Week, DRUG = Drug Experimentation, SI = Smoking Initiation, PTU = Problem Tobacco Use, OUD = Opioid Use Disorder, PAU = Problem Alcohol Use, CUD = Cannabis Use Disorder, RISK = Risk Tolerance, NSEX = Number of Lifetime Sexual Partners, ADHD = Attention Deficit/Hyperactivity Disorder, FSEX = Age at First Sexual Intercourse (reverse-coded). BD, SUB, and SUD are standardized.
9. **The parameters associated with Model 1, with residuals of PTU and SI, CUD and CI, and PAU and DPW correlated.** Unstand Est = unstandardized estimate; SE = standard error; STD_Genotype estimates are standardized with respect to the indicators; STD_All estimates are standardized with respect to the full model, including endogenous latent factors. SUDG1a = The higher-order Substance Use Disorder factor; SUBResG1a = The higher-order Residual Substance Use Factor; BDResG1a = The higher-order Behavioral Disinhibition-Specific factor. BD = (lower-order) Behavioral Disinhibition; SUB = (lower-order) Substance Use; SUD = (lower-order) Substance Use Disorder; CI = Cannabis Initiation, DPW = Drinks per Week, DRUG = Drug Experimentation, SI = Smoking Initiation, PTU = Problem Tobacco Use, OUD = Opioid Use Disorder, PAU = Problem Alcohol Use, CUD = Cannabis Use Disorder, RISK = Risk Tolerance, NSEX = Number of Lifetime Sexual Partners, ADHD = Attention Deficit/Hyperactivity Disorder, FSEX = Age at First Sexual Intercourse (reverse-coded). SUDG1a, SUBResG1a, and BDResG1a are standardized.
10. **The parameters associated with Model 2, with residuals of PTU and SI, CUD and CI, and PAU and DPW correlated.** Unstand Est = unstandardized estimate; SE = standard error; STD_Genotype estimates are standardized with respect to the indicators; STD_All estimates are standardized with respect to the full model, including endogenous latent factors. SUBG1b = The higher-order Substance Use factor; SUDResG1b = The higher-order Residual Substance Use Disorder Factor; BDResG1b = The higher-order Behavioral Disinhibition-Specific factor. BD = (lower-order) Behavioral Disinhibition; SUB = (lower-order) Substance Use; SUD = (lower-order) Substance Use Disorder; CI = Cannabis Initiation, DPW = Drinks per Week, DRUG = Drug Experimentation, SI = Smoking Initiation, PTU = Problem Tobacco Use, OUD = Opioid Use Disorder, PAU = Problem Alcohol Use, CUD = Cannabis Use Disorder, RISK = Risk Tolerance, NSEX = Number of Lifetime Sexual Partners, ADHD = Attention Deficit/Hyperactivity Disorder, FSEX = Age at First Sexual Intercourse (reverse-coded). SUBG1b, SUDResG1b, and BDResG1b are standardized.
11. **The parameter associated with a standardized correlated factors model with modified lower-order SU, modified lower-order SUD, and BD, with residuals of PTU and SI, CUD and CI, and PAU and DPW correlated.** Unstand Est = unstandardized estimate; SE = standard error; STD_Genotype estimates are standardized with respect to the indicators; STD_All estimates are standardized with respect to the full model (equivalent to STD_Genotype in the correlated factors model). BD = Behavioral Disinhibition; SUB = Modified Substance Use; SUD = Modified Substance Use Disorder; CI = Cannabis Initiation, DPW = Drinks per Week, SI = Smoking Initiation, PTU = Problem Tobacco Use, PAU = Problem Alcohol Use, CUD = Cannabis Use Disorder, RISK = Risk Tolerance, NSEX = Number of Lifetime Sexual Partners, ADHD = Attention Deficit/Hyperactivity Disorder, FSEX = Age at First Sexual Intercourse (reverse-coded). The three-indicator modified lower-order SU factor and the three-indicator modified lower-order SUD factor collectively include only cannabis, alcohol, and nicotine phenotypes. BD, SUB, and SUD are standardized.
12. **The parameters associated with Modified Model 1, with residuals of PTU and SI, CUD and CI, and PAU and DPW correlated.** Unstand Est = unstandardized estimate; SE = standard error; STD_Genotype estimates are standardized with respect to the indicators; STD_All estimates are standardized with respect to the full model, including endogenous latent factors. SUDG1c = The higher-order Modified Substance Use Disorder factor; SUBResG1c = The higher-order Modified Residual Substance Use Factor; BDResG1c = The higher-order Modified Residual Behavioral Disinhibition-Specific factor. BD = (lower-order) Behavioral Disinhibition; SUB = (lower-order) Modified Substance Use; SUD = (lower-order) Modified Substance Use Disorder; CI = Cannabis Initiation, DPW = Drinks per Week, SI = Smoking Initiation, PTU = Problem Tobacco Use, PAU = Problem Alcohol Use, CUD = Cannabis Use Disorder, RISK = Risk Tolerance, NSEX = Number of Lifetime Sexual Partners, ADHD = Attention Deficit/Hyperactivity Disorder, FSEX = Age at First Sexual Intercourse (reverse-coded). The three-indicator modified lower-order SU factor and the three-indicator modified lower-order SUD factor collectively include only cannabis, alcohol, and nicotine phenotypes. SUDG1c, SUBResG1c, and BDResG1c are standardized.
13. **The parameters associated with Modified Model 2, with residuals of PTU and SI, CUD and CI, and PAU and DPW correlated.** Unstand Est = unstandardized estimate; SE = standard error; STD_Genotype estimates are standardized with respect to the indicators; STD_All estimates are standardized with respect to the full model, including endogenous latent factors. SUBG1d = The higher-order Modified Substance Use factor; SUDResG1d = The higher-order Modified Residual Substance Use Disorder Factor; BDResG1d = The higher-order Modified Residual Behavioral Disinhibition-Specific factor. BD = (lower-order) Behavioral Disinhibition; SUB = (lower-order) Modified Substance Use; SUD = (lower-order) Modified Substance Use Disorder; CI = Cannabis Initiation, DPW = Drinks per Week, SI = Smoking Initiation, PTU = Problem Tobacco Use, PAU = Problem Alcohol Use, CUD = Cannabis Use Disorder, RISK = Risk Tolerance, NSEX = Number of Lifetime Sexual Partners, ADHD = Attention Deficit/Hyperactivity Disorder, FSEX = Age at First Sexual Intercourse (reverse-coded). The three-indicator modified lower-order SU factor and the three-indicator modified lower-order SUD factor collectively include only cannabis, alcohol, and nicotine phenotypes. SUBG1d, SUDResG1d, and BDResG1d are standardized.
14. **The parameters associated with a correlated factors model of the original SUB, SUD, and BD factors, with no correlated residuals.** Unstand Est = unstandardized estimate; SE = standard error; STD_Genotype estimates are standardized with respect to the indicators; STD_All estimates are standardized with respect to the full model (equivalent to STD_Genotype in the correlated factors model). BD = Behavioral Disinhibition; SUB = Substance Use; SUD = Substance Use Disorder; CI = Cannabis Initiation, DPW = Drinks per Week, DRUG = Drug Experimentation, SI = Smoking Initiation, PTU = Problem Tobacco Use, OUD = Opioid Use Disorder, PAU = Problem Alcohol Use, CUD = Cannabis Use Disorder, RISK = Risk Tolerance, NSEX = Number of Lifetime Sexual Partners, ADHD = Attention Deficit/Hyperactivity Disorder, FSEX = Age at First Sexual Intercourse (reverse-coded). BD, SUB, and SUD are standardized.
15. **The parameter associated with a standardized correlated factors model with modified lower-order SU, modified lower-order SUD, and BD, with no correlated residuals.** Unstand Est = unstandardized estimate; SE = standard error; STD_Genotype estimates are standardized with respect to the indicators; STD_All estimates are standardized with respect to the full model (equivalent to STD_Genotype in the correlated factors model). BD = Behavioral Disinhibition; SUB = Modified Substance Use; SUD = Modified Substance Use Disorder; CI = Cannabis Initiation, DPW = Drinks per Week, SI = Smoking Initiation, PTU = Problem Tobacco Use, PAU = Problem Alcohol Use, CUD = Cannabis Use Disorder, RISK = Risk Tolerance, NSEX = Number of Lifetime Sexual Partners, ADHD = Attention Deficit/Hyperactivity Disorder, FSEX = Age at First Sexual Intercourse (reverse-coded). The three-indicator modified lower-order SU factor and the three-indicator modified lower-order SUD factor collectively include only cannabis, alcohol, and nicotine phenotypes. BD, SUB, and SUD are standardized.
16. **The parameters associated with Model 1, with no correlated residuals.** Unstand Est = unstandardized estimate; SE = standard error; STD_Genotype estimates are standardized with respect to the indicators; STD_All estimates are standardized with respect to the full model, including endogenous latent factors. SUDG1a = The higher-order Substance Use Disorder factor; SUBResG1a = The higher-order Residual Substance Use factor; BDResG1a = The higher-order Behavioral Disinhibition-Specific factor. BD = (lower-order) Behavioral Disinhibition; SUB = (lower-order) Substance Use; SUD = (lower-order) Substance Use Disorder; CI = Cannabis Initiation, DPW = Drinks per Week, DRUG = Drug Experimentation, SI = Smoking Initiation, PTU = Problem Tobacco Use, OUD = Opioid Use Disorder, PAU = Problem Alcohol Use, CUD = Cannabis Use Disorder, RISK = Risk Tolerance, NSEX = Number of Lifetime Sexual Partners, ADHD = Attention Deficit/Hyperactivity Disorder, FSEX = Age at First Sexual Intercourse (reverse-coded). SUDG1a, SUBResG1a, and BDResG1a are standardized.
17. **The parameters associated with Model 2, with no correlated residuals.** Unstand Est = unstandardized estimate; SE = standard error; STD_Genotype estimates are standardized with respect to the indicators; STD_All estimates are standardized with respect to the full model, including endogenous latent factors. SUBG1b = The higher-order Substance Use factor; SUDResG1b = The higher-order Residual Substance Use Disorder factor; BDResG1b = The higher-order Residual Behavioral Disinhibition-Specific factor. BD = (lower-order) Behavioral Disinhibition; SUB = (lower-order) Substance Use; SUD = (lower-order) Substance Use Disorder; CI = Cannabis Initiation, DPW = Drinks per Week, DRUG = Drug Experimentation, SI = Smoking Initiation, PTU = Problem Tobacco Use, OUD = Opioid Use Disorder, PAU = Problem Alcohol Use, CUD = Cannabis Use Disorder, RISK = Risk Tolerance, NSEX = Number of Lifetime Sexual Partners, ADHD = Attention Deficit/Hyperactivity Disorder, FSEX = Age at First Sexual Intercourse (reverse-coded). SUBG1b, SUDResG1b, and BDResG1b are standardized.
18. **The parameters associated with Modified Model 1, with no correlated residuals.** Unstand Est = unstandardized estimate; SE = standard error; STD_Genotype estimates are standardized with respect to the indicators; STD_All estimates are standardized with respect to the full model, including endogenous latent factors. SUDG1c = The higher-order Modified Substance Use Disorder factor; SUBResG1c = The higher-order Modified Residual Substance Use factor; BDResG1c = The higher-order Modified Residual Behavioral Disinhibition-Specific factor. BD = (lower-order) Behavioral Disinhibition; SUB = (lower-order) Modified Substance Use; SUD = (lower-order) Modified Substance Use Disorder; CI = Cannabis Initiation, DPW = Drinks per Week, SI = Smoking Initiation, PTU = Problem Tobacco Use, PAU = Problem Alcohol Use, CUD = Cannabis Use Disorder, RISK = Risk Tolerance, NSEX = Number of Lifetime Sexual Partners, ADHD = Attention Deficit/Hyperactivity Disorder, FSEX = Age at First Sexual Intercourse (reverse-coded). The three-indicator modified lower-order SU factor and the three-indicator modified lower-order SUD factor collectively include only cannabis, alcohol, and nicotine phenotypes. SUDG1c, SUBResG1c, and BDResG1c are standardized.
19. **The parameters associated with Modified Model 2, with no correlated residuals.** Unstand Est = unstandardized estimate; SE = standard error; STD_Genotype estimates are standardized with respect to the indicators; STD_All estimates are standardized with respect to the full model, including endogenous latent factors. SUBG1d = The higher-order Modified Substance Use factor; SUDResG1d = The higher-order Modified Residual Substance Use Disorder factor; BDResG1d = The higher-order Modified Residual Behavioral Disinhibition-Specific factor. BD = (lower-order) Behavioral Disinhibition; SUB = (lower-order) Modified Substance Use; SUD = (lower-order) Modified Substance Use Disorder; CI = Cannabis Initiation, DPW = Drinks per Week, SI = Smoking Initiation, PTU = Problem Tobacco Use, PAU = Problem Alcohol Use, CUD = Cannabis Use Disorder, RISK = Risk Tolerance, NSEX = Number of Lifetime Sexual Partners, ADHD = Attention Deficit/Hyperactivity Disorder, FSEX = Age at First Sexual Intercourse (reverse-coded). The three-indicator modified lower-order SU factor and the three-indicator modified lower-order SUD factor collectively include only cannabis, alcohol, and nicotine phenotypes. SUBG1d, SUDResG1d, and BDResG1d are standardized.
