## Supplementary Tables for "Partitioning the Genomic Components of Behavioral Disinhibition and Substance Use (Disorder) Using Genomic Structural Equation Modeling"

| **Factor/Indicator** | | **Unstd. Est.** | **Unstd. SE** | **Std. Genotype** | **Std. Genotype SE** | **Std. All** | ***P*-value** |
| --- | --- | --- | --- | --- | --- | --- | --- |
| **Loadings** | | | | | | | |
| BD | FSEX | 0.324 | 0.009 | 0.836 | 0.022 | 0.836 | < 5e-300 |
|  | ADHD | 0.296 | 0.015 | 0.656 | 0.033 | 0.656 | 0.000 |
|  | NSEX | 0.238 | 0.006 | 0.757 | 0.021 | 0.757 | 0.000 |
|  | RISK | 0.180 | 0.008 | 0.601 | 0.027 | 0.601 | 0.000 |
| **Residuals** | | | | | | | |
| ADHD | ADHD | 0.116 | 0.009 | 0.570 | 0.044 | 0.570 | 0.000 |
| FSEX | FSEX | 0.045 | 0.005 | 0.300 | 0.030 | 0.300 | 0.000 |
| NSEX | NSEX | 0.042 | 0.003 | 0.426 | 0.026 | 0.426 | 0.000 |
| RISK | RISK | 0.058 | 0.003 | 0.639 | 0.035 | 0.639 | 0.000 |
| **Variances & Covariances** | | | | | | | |
| BD | BD | 1.000 | NA | 1.000 | NA | 1.000 | NA |

Supplementary Table 1

| **Factor**/**Indicator** | | **Unstd. Est.** | **Unstd. SE** | **Std. Genotype** | **Std. Genotype SE** | **Std. All** | ***P*-value** |
| --- | --- | --- | --- | --- | --- | --- | --- |
| **Loadings** | | | | | | | |
| SUD | PTU | 0.211 | 0.028 | 0.575 | 0.077 | 0.575 | 0.000 |
|  | CUD | 0.259 | 0.020 | 0.787 | 0.059 | 0.787 | 0.000 |
|  | PAU | 0.176 | 0.011 | 0.699 | 0.044 | 0.699 | 0.000 |
|  | OUD | 0.356 | 0.026 | 1.014 | 0.075 | 1.014 | 0.000 |
| **Residuals** | | | | | | | |
| OUD | OUD | -0.003 | 0.022 | -0.028 | 0.175 | -0.028 | 0.874 |
| PAU | PAU | 0.033 | 0.005 | 0.511 | 0.075 | 0.511 | 0.000 |
| CUD | CUD | 0.041 | 0.012 | 0.381 | 0.111 | 0.381 | 0.001 |
| PTU | PTU | 0.090 | 0.034 | 0.670 | 0.249 | 0.670 | 0.007 |
| **Variances & Covariances** | | | | | | | |
| SUD | SUD | 1.000 | NA | 1.000 | NA | 1.000 | NA |

Supplementary Table 2

| **Factor**/**Indicator** | | **Unstd. Est.** | **Unstd. SE** | **Std. Genotype** | **Std. Genotype SE** | **Std. All** | ***P-value*** |
| --- | --- | --- | --- | --- | --- | --- | --- |
| F1 | CI | 0.169 | 0.011 | 0.467 | 0.031 | 0.467 | 3.30E-51 |
|  | DPW | 0.168 | 0.005 | -0.126 | 0.028 | 1.013 | 9.89E-286 |
|  | SI | 0.096 | 0.007 | 0.295 | 0.021 | 0.295 | 2.18E-46 |
|  | FREQ | 0.205 | 0.007 | 0.829 | 0.03 | 0.829 | 2.65E-172 |
|  | Drink.Stat | 0.192 | 0.008 | 0.713 | 0.03 | 0.713 | 6.87E-128 |
|  | DRUG | 0.214 | 0.022 | 0.641 | 0.065 | 0.641 | 5.39E-23 |
|  | CPD | -0.036 | 0.006 | 1.013 | 0.022 | -0.126 | 7.41E-09 |
| CI | CI | 0.103 | 0.008 | 0.782 | 0.059 | 0.782 | 1.48E-40 |
| CPD | CPD | 0.081 | 0.004 | 0.984 | 0.049 | 0.984 | 1.65E-89 |
| SI | SI | 0.096 | 0.003 | 0.913 | 0.03 | 0.913 | 5.02E-197 |
| FREQ | FREQ | 0.019 | 0.002 | 0.313 | 0.04 | 0.313 | 4.30E-15 |
| Drink.Stat | Drink.Stat | 0.036 | 0.003 | 0.492 | 0.041 | 0.492 | 9.24E-34 |
| DRUG | DRUG | 0.066 | 0.022 | 0.589 | 0.195 | 0.589 | 2.50E-03 |
| DPW | DPW | -0.001 | 0.001 | -0.025 | 0.044 | -0.025 | 5.64E-01 |
| F1 | F1 | 1.000 | NA | 1.000 | NA | 1.000 | NA |

Supplementary Table 3.

| **Factor**/**Indicator** | | **Unstd. Est.** | **Unstd. SE** | **Std. Genotype** | **Std. Genotype SE** | **Std. All** | ***P-value*** |
| --- | --- | --- | --- | --- | --- | --- | --- |
| F1 | CI | 0.17 | 0.011 | 0.469 | 0.031 | 0.469 | 1.86E-51 |
|  | DPW | 0.165 | 0.005 | 1.004 | 0.028 | 1.004 | 2.49E-275 |
|  | SI | 0.096 | 0.007 | 0.295 | 0.021 | 0.295 | 3.82E-46 |
|  | FREQ | 0.206 | 0.007 | 0.83 | 0.03 | 0.83 | 6.73E-172 |
|  | Drink.Stat | 0.193 | 0.008 | 0.715 | 0.03 | 0.715 | 1.45E-128 |
|  | DRUG | 0.215 | 0.022 | 0.642 | 0.065 | 0.642 | 4.75E-23 |
|  | CPD | -0.036 | 0.006 | -0.126 | 0.022 | -0.126 | 6.23E-09 |
| DPW | DPW | 0.001 | 0.001 | 0.001 | 0.043 | 0.001 | 3.95E-01 |
| CI | CI | 0.102 | 0.008 | 0.78 | 0.059 | 0.78 | 3.78E-40 |
| CPD | CPD | 0.081 | 0.004 | 0.984 | 0.049 | 0.984 | 1.88E-89 |
| SI | SI | 0.096 | 0.003 | 0.913 | 0.03 | 0.913 | 1.09E-196 |
| FREQ | FREQ | 0.019 | 0.002 | 0.311 | 0.04 | 0.311 | 1.86E-14 |
| Drink.Stat | Drink.Stat | 0.035 | 0.003 | 0.489 | 0.041 | 0.489 | 2.19E-32 |
| DRUG | DRUG | 0.065 | 0.022 | 0.588 | 0.195 | 0.588 | 2.66E-03 |
| F1 | F1 | 1.000 | NA | 1.000 | NA | 1.000 | NA |

Supplementary Table 4.

| **Factor**/**Indicator** | | **Unstd. Est.** | **Unstd. SE** | **Std. Genotype** | **Std. Genotype SE** | **Std. All** | ***P-value*** |
| --- | --- | --- | --- | --- | --- | --- | --- |
| F1 | CI | 0.175 | 0.011 | 0.482 | 0.031 | 0.482 | 3.87E-55 |
|  | DPW | 0.165 | 0.005 | 1.007 | 0.028 | 1.007 | 7.25E-268 |
|  | SI | 0.108 | 0.007 | 0.332 | 0.021 | 0.332 | 1.84E-56 |
|  | FREQ | 0.201 | 0.007 | 0.811 | 0.03 | 0.811 | 1.03E-163 |
|  | Drink.Stat | 0.18 | 0.008 | 0.665 | 0.03 | 0.665 | 4.55E-109 |
|  | DRUG | 0.228 | 0.022 | 0.68 | 0.065 | 0.68 | 2.23E-25 |
| DPW | DPW | 0.001 | 0.001 | 0.001 | 0.044 | 0.001 | 4.03E-01 |
| CI | CI | 0.1 | 0.008 | 0.768 | 0.058 | 0.768 | 4.19E-39 |
| SI | SI | 0.093 | 0.003 | 0.889 | 0.03 | 0.889 | 9.07E-193 |
| FREQ | FREQ | 0.021 | 0.002 | 0.343 | 0.041 | 0.343 | 5.56E-17 |
| Drink.Stat | Drink.Stat | 0.04 | 0.003 | 0.558 | 0.041 | 0.558 | 2.98E-41 |
| DRUG | DRUG | 0.059 | 0.021 | 0.538 | 0.192 | 0.538 | 5.43E-03 |
| F1 | F1 | 1.000 | NA | 1.000 | NA | 1.000 | NA |

Supplementary Table 5.

| **Factor**/**Indicator** | | **Unstd. Est.** | **Unstd. SE** | **Std. Genotype** | **Std. Genotype SE** | **Std. All** | ***P-value*** |
| --- | --- | --- | --- | --- | --- | --- | --- |
| F1 | CI | 0.261 | 0.013 | 0.72 | 0.036 | 0.72 | 6.80E-89 |
|  | DPW | 0.08 | 0.005 | 0.483 | 0.028 | 0.483 | 1.53E-66 |
|  | SI | 0.249 | 0.01 | 0.77 | 0.03 | 0.77 | 1.36E-145 |
|  | DRUG | 0.357 | 0.029 | 1.071 | 0.086 | 1.071 | 6.06E-36 |
| DPW | DPW | 0.021 | 0.001 | 0.766 | 0.047 | 0.766 | 1.23E-60 |
| CI | CI | 0.063 | 0.007 | 0.482 | 0.053 | 0.482 | 1.26E-19 |
| SI | SI | 0.043 | 0.004 | 0.407 | 0.039 | 0.407 | 1.44E-25 |
| DRUG | DRUG | -0.016 | 0.02 | -0.148 | 0.178 | -0.148 | 4.07E-01 |
| F1 | F1 | 1.000 | NA | 1.000 | NA | 1.000 | NA |

Supplementary Table 6.

| **Factor**/**Indicator** | | **Unstd. Est.** | **Unstd. SE** | **Std. Genotype** | **Std. Genotype SE** | **Std. All** | ***P-value*** |
| --- | --- | --- | --- | --- | --- | --- | --- |
| F1 | CI | 0.261 | 0.013 | 0.72 | 0.036 | 0.72 | 3.35E-88 |
|  | DPW | 0.08 | 0.005 | 0.483 | 0.028 | 0.483 | 4.10E-66 |
|  | SI | 0.251 | 0.01 | 0.774 | 0.03 | 0.774 | 1.51E-144 |
|  | DRUG | 0.35 | 0.028 | 1.05 | 0.085 | 1.05 | 1.04E-34 |
| DPW | DPW | 0.021 | 0.001 | 0.767 | 0.047 | 0.767 | 1.04E-60 |
| DRUG | DRUG | 0.001 | 0.02 | 0.001 | 0.177 | 0.001 | 9.59E-01 |
| CI | CI | 0.063 | 0.007 | 0.481 | 0.053 | 0.481 | 1.81E-19 |
| SI | SI | 0.042 | 0.004 | 0.401 | 0.04 | 0.401 | 5.73E-24 |
| F1 | F1 | 1.000 | NA | 1.000 | NA | 1.000 | NA |

Supplementary Table 7.

| **Factor**/**Indicator** | | **Unstd. Est.** | **Unstd. SE** | **Std. Genotype** | **Std. Genotype SE** | **Std. All** | ***P*-value** |
| --- | --- | --- | --- | --- | --- | --- | --- |
| **Loadings** | | | | | | | |
| F1 | FSEX | 0.285 | 0.007 | 0.734 | 0.017 | 0.734 | < 5e-300 |
|  | ADHD | 0.278 | 0.013 | 0.617 | 0.030 | 0.617 | 0.000 |
|  | NSEX | 0.242 | 0.005 | 0.766 | 0.017 | 0.766 | < 5e-300 |
|  | RISK | 0.160 | 0.008 | 0.532 | 0.025 | 0.532 | 0.000 |
|  | CI | 0.222 | 0.010 | 0.611 | 0.028 | 0.611 | 0.000 |
|  | DPW | 0.056 | 0.004 | 0.333 | 0.021 | 0.333 | 0.000 |
|  | SI | 0.271 | 0.006 | 0.828 | 0.019 | 0.828 | < 5e-300 |
|  | DRUG | 0.296 | 0.021 | 0.868 | 0.063 | 0.868 | 0.000 |
|  | PTU | 0.145 | 0.020 | 0.395 | 0.055 | 0.395 | 0.000 |
|  | CUD | 0.286 | 0.011 | 0.863 | 0.033 | 0.863 | 0.000 |
|  | PAU | 0.157 | 0.006 | 0.623 | 0.024 | 0.623 | 0.000 |
|  | OUD | 0.237 | 0.015 | 0.674 | 0.042 | 0.674 | 0.000 |
| **Residuals** | | | | | | | |
| SI | SI | 0.033 | 0.002 | 0.314 | 0.018 | 0.314 | 0.000 |
| CI | CI | 0.082 | 0.007 | 0.627 | 0.050 | 0.627 | 0.000 |
| DRUG | DRUG | 0.029 | 0.020 | 0.246 | 0.172 | 0.246 | 0.153 |
| DPW | DPW | 0.025 | 0.001 | 0.889 | 0.043 | 0.889 | 0.000 |
| FSEX | FSEX | 0.069 | 0.003 | 0.461 | 0.022 | 0.461 | 0.000 |
| ADHD | ADHD | 0.126 | 0.008 | 0.619 | 0.040 | 0.619 | 0.000 |
| NSEX | NSEX | 0.041 | 0.002 | 0.413 | 0.023 | 0.413 | 0.000 |
| RISK | RISK | 0.065 | 0.003 | 0.716 | 0.035 | 0.716 | 0.000 |
| OUD | OUD | 0.067 | 0.019 | 0.545 | 0.154 | 0.545 | 0.000 |
| CUD | CUD | 0.028 | 0.009 | 0.255 | 0.077 | 0.255 | 0.001 |
| PAU | PAU | 0.039 | 0.003 | 0.611 | 0.047 | 0.611 | 0.000 |
| PTU | PTU | 0.114 | 0.032 | 0.844 | 0.235 | 0.844 | 0.000 |
| DPW | PAU | 0.019 | 0.002 | 0.440 | 0.036 | 0.596 | 0.000 |
| CI | CUD | -0.002 | 0.006 | -0.019 | 0.046 | -0.047 | 0.684 |
| SI | PTU | 0.006 | 0.006 | 0.047 | 0.047 | 0.090 | 0.320 |
| **Variances & Covariances** | | | | | | | |
| F1 | F1 | 1.000 | NA | 1.000 | NA | 1.000 | NA |

Supplementary Table 8.

| **Factor/Indicator** | | **Unstd. Est.** | **Unstd. SE** | **Std. Genotype** | **Std. Genotype SE** | **Std. All** | ***P-value*** |
| --- | --- | --- | --- | --- | --- | --- | --- |
| **Loadings** | | | | | | | |
| BD | FSEX | 0.309 | 0.007 | 0.796 | 0.018 | 0.796 | < 5e-300 |
|  | ADHD | 0.295 | 0.014 | 0.654 | 0.031 | 0.654 | 1.20E-96 |
|  | NSEX | 0.258 | 0.006 | 0.816 | 0.018 | 0.816 | < 5e-300 |
|  | RISK | 0.17 | 0.008 | 0.566 | 0.026 | 0.566 | 7.10E-105 |
| SUB | CI | 0.238 | 0.011 | 0.657 | 0.029 | 0.657 | 1.86E-113 |
|  | DPW | 0.062 | 0.004 | 0.367 | 0.022 | 0.367 | 3.26E-62 |
|  | SI | 0.304 | 0.008 | 0.93 | 0.024 | 0.93 | < 5e-300 |
|  | DRUG | 0.319 | 0.023 | 0.935 | 0.066 | 0.935 | 2.56E-45 |
| SUD | PTU | 0.165 | 0.023 | 0.449 | 0.062 | 0.449 | 3.23E-13 |
|  | CUD | 0.319 | 0.013 | 0.961 | 0.039 | 0.961 | 9.54E-133 |
|  | PAU | 0.178 | 0.007 | 0.706 | 0.027 | 0.706 | 1.97E-150 |
|  | OUD | 0.268 | 0.016 | 0.763 | 0.046 | 0.763 | 6.09E-63 |
| **Residuals** | | | | | | | |
| DPW | DPW | 0.024 | 0.001 | 0.865 | 0.043 | 0.865 | 1.40E-90 |
| DRUG | DRUG | 0.015 | 0.02 | 0.126 | 0.17 | 0.126 | 4.59E-01 |
| CI | CI | 0.075 | 0.007 | 0.568 | 0.05 | 0.568 | 5.28E-30 |
| SI | SI | 0.014 | 0.003 | 0.136 | 0.03 | 0.136 | 7.26E-06 |
| FSEX | FSEX | 0.055 | 0.003 | 0.367 | 0.022 | 0.367 | 3.16E-62 |
| ADHD | ADHD | 0.116 | 0.008 | 0.572 | 0.041 | 0.572 | 1.71E-44 |
| NSEX | NSEX | 0.033 | 0.002 | 0.334 | 0.023 | 0.334 | 1.90E-48 |
| RISK | RISK | 0.061 | 0.003 | 0.679 | 0.035 | 0.679 | 1.80E-83 |
| OUD | OUD | 0.052 | 0.019 | 0.418 | 0.151 | 0.418 | 5.76E-03 |
| CUD | CUD | 0.008 | 0.01 | 0.077 | 0.089 | 0.077 | 3.89E-01 |
| PAU | PAU | 0.032 | 0.003 | 0.502 | 0.052 | 0.502 | 3.02E-22 |
| PTU | PTU | 0.108 | 0.032 | 0.799 | 0.235 | 0.799 | 6.95E-04 |
| CI | CUD | 0 | 0.005 | 0.001 | 0.045 | 0.005 | 9.82E-01 |
| SI | PTU | 0.005 | 0.006 | 0.039 | 0.047 | 0.118 | 4.10E-01 |
| DPW | PAU | 0.019 | 0.002 | 0.439 | 0.036 | 0.666 | 1.07E-34 |
| **Variances & Covariances** | | | | | | | |
| BD | BD | 1.00 | NA | 1.00 | NA | 1.00 | NA |
| SUB | SUB | 1.00 | NA | 1.00 | NA | 1.00 | NA |
| SUD | SUD | 1.00 | NA | 1.00 | NA | 1.00 | NA |
| BD | SUB | 0.778 | 0.016 | 0.778 | 0.016 | 0.778 | < 5e-300 |
| BD | SUD | 0.774 | 0.027 | 0.774 | 0.027 | 0.774 | 0 |
| SUB | SUD | 0.803 | 0.027 | 0.803 | 0.027 | 0.803 | 0 |

Supplementary Table 9.

| **Factor**/**Indicator** | | **Unstd. Est.** | **Unstd. SE** | **Std. Genotype** | **Std. Genotype SE** | **Std. All** | ***P*-value** |
| --- | --- | --- | --- | --- | --- | --- | --- |
| **Higher-order Loadings** | | | | | | | |
| SUDG1a | SUD | 0.165 | 0.023 | 0.449 | 0.062 | 0.449 | 0.000 |
|  | SUB | 0.192 | 0.011 | 0.528 | 0.030 | 0.803 | 0.000 |
|  | BD | 0.239 | 0.009 | 0.616 | 0.024 | 0.774 | 0.000 |
| SUBResG1a | SUB | 0.142 | 0.010 | 0.392 | 0.028 | 0.596 | 0.000 |
|  | BD | 0.081 | 0.016 | 0.209 | 0.041 | 0.262 | 0.000 |
| BDResG1a | BD | 0.178 | 0.008 | 0.459 | 0.021 | 0.577 | 0.000 |
| **Lower-order Loadings** | | | | | | | |
| SUD | PTU | 1.000 | NA | 1.000 | NA | 0.449 | NA |
|  | CUD | 1.933 | 0.271 | 2.141 | 0.300 | 0.961 | 0.000 |
|  | PAU | 1.079 | 0.151 | 1.572 | 0.220 | 0.706 | 0.000 |
|  | OUD | 1.625 | 0.243 | 1.700 | 0.254 | 0.763 | 0.000 |
| SUB | CI | 1.000 | NA | 1.000 | NA | 0.657 | NA |
|  | DRUG | 1.339 | 0.098 | 1.422 | 0.104 | 0.935 | 0.000 |
|  | SI | 1.274 | 0.057 | 1.414 | 0.063 | 0.930 | 0.000 |
|  | DPW | 0.259 | 0.017 | 0.559 | 0.038 | 0.367 | 0.000 |
| BD | FSEX | 1.000 | NA | 1.000 | NA | 0.796 | NA |
|  | ADHD | 0.956 | 0.051 | 0.822 | 0.044 | 0.654 | 0.000 |
|  | NSEX | 0.837 | 0.021 | 1.026 | 0.026 | 0.816 | < 5e-300 |
|  | RISK | 0.551 | 0.024 | 0.712 | 0.031 | 0.566 | 0.000 |
| **Residuals** | | | | | | | |
| CI | CI | 0.075 | 0.007 | 0.568 | 0.050 | 0.568 | 0.000 |
| DPW | DPW | 0.024 | 0.001 | 0.865 | 0.043 | 0.865 | 0.000 |
| SI | SI | 0.014 | 0.003 | 0.136 | 0.030 | 0.136 | 0.000 |
| DRUG | DRUG | 0.015 | 0.020 | 0.126 | 0.170 | 0.126 | 0.459 |
| FSEX | FSEX | 0.055 | 0.003 | 0.367 | 0.022 | 0.367 | 0.000 |
| ADHD | ADHD | 0.116 | 0.008 | 0.572 | 0.041 | 0.572 | 0.000 |
| NSEX | NSEX | 0.033 | 0.002 | 0.334 | 0.023 | 0.334 | 0.000 |
| RISK | RISK | 0.061 | 0.003 | 0.679 | 0.035 | 0.679 | 0.000 |
| OUD | OUD | 0.052 | 0.019 | 0.418 | 0.151 | 0.418 | 0.006 |
| CUD | CUD | 0.008 | 0.010 | 0.077 | 0.089 | 0.077 | 0.389 |
| PAU | PAU | 0.032 | 0.003 | 0.502 | 0.052 | 0.502 | 0.000 |
| PTU | PTU | 0.108 | 0.032 | 0.799 | 0.235 | 0.799 | 0.001 |
| SI | PTU | 0.005 | 0.006 | 0.039 | 0.047 | 0.118 | 0.410 |
| DPW | PAU | 0.019 | 0.002 | 0.439 | 0.036 | 0.666 | 0.000 |
| CI | CUD | 0.000 | 0.005 | 0.001 | 0.045 | 0.005 | 0.982 |
| SUD | SUD | 0.000 | 0.000 | 0.000 | NA | NA | NA |
| SUB | SUB | 0.000 | 0.000 | 0.000 | NA | NA | NA |
| BD | BD | 0.000 | 0.000 | 0.000 | NA | NA | NA |
| **Variances and covariances** | | | | | | | |
| SUDG1a | SUDG1a | 1.000 | NA | 1.000 | NA | 1.000 | NA |
| SUBResG1a | SUBResG1a | 1.000 | NA | 1.000 | NA | 1.000 | NA |
| BDResG1a | BDResG1a | 1.000 | NA | 1.000 | NA | 1.000 | NA |

Supplementary Table 10.

| **Factor**/**Indicator** | | **Unstd. Est.** | **Unstd. SE** | **Std. Genotype** | **Std. Genotype SE** | **Std. All** | ***P*-value** |
| --- | --- | --- | --- | --- | --- | --- | --- |
| **Higher-order Loadings** | | | | | | | |
| SUBG1b | SUB | 0.238 | 0.011 | 0.657 | 0.029 | 0.657 | 0.000 |
|  | SUD | 0.133 | 0.018 | 0.360 | 0.049 | 0.803 | 0.000 |
|  | BD | 0.240 | 0.007 | 0.619 | 0.018 | 0.778 | 0.000 |
| SUDResG1b | SUD | 0.098 | 0.016 | 0.267 | 0.042 | 0.596 | 0.000 |
|  | BD | 0.077 | 0.013 | 0.199 | 0.034 | 0.250 | 0.000 |
| BDResG1b | BD | 0.178 | 0.008 | 0.459 | 0.021 | 0.577 | 0.000 |
| **Lower-order Loadings** | | | | | | | |
| SUD | PTU | 1.000 | NA | 1.000 | NA | 0.449 | NA |
|  | CUD | 1.933 | 0.271 | 2.141 | 0.300 | 0.961 | 0.000 |
|  | PAU | 1.079 | 0.151 | 1.572 | 0.220 | 0.706 | 0.000 |
|  | OUD | 1.625 | 0.243 | 1.700 | 0.254 | 0.763 | 0.000 |
| SUB | CI | 1.000 | NA | 1.000 | NA | 0.657 | NA |
|  | DRUG | 1.339 | 0.098 | 1.422 | 0.104 | 0.935 | 0.000 |
|  | SI | 1.274 | 0.057 | 1.414 | 0.063 | 0.930 | 0.000 |
|  | DPW | 0.259 | 0.017 | 0.559 | 0.038 | 0.367 | 0.000 |
| BD | FSEX | 1.000 | NA | 1.000 | NA | 0.796 | NA |
|  | ADHD | 0.956 | 0.051 | 0.822 | 0.044 | 0.654 | 0.000 |
|  | NSEX | 0.837 | 0.021 | 1.026 | 0.026 | 0.816 | < 5e-300 |
|  | RISK | 0.551 | 0.024 | 0.712 | 0.031 | 0.566 | 0.000 |
| **Residuals** | | | | | | | |
| CI | CI | 0.075 | 0.007 | 0.568 | 0.050 | 0.568 | 0.000 |
| DPW | DPW | 0.024 | 0.001 | 0.865 | 0.043 | 0.865 | 0.000 |
| SI | SI | 0.014 | 0.003 | 0.136 | 0.030 | 0.136 | 0.000 |
| DRUG | DRUG | 0.015 | 0.020 | 0.126 | 0.170 | 0.126 | 0.459 |
| FSEX | FSEX | 0.055 | 0.003 | 0.367 | 0.022 | 0.367 | 0.000 |
| ADHD | ADHD | 0.116 | 0.008 | 0.572 | 0.041 | 0.572 | 0.000 |
| NSEX | NSEX | 0.033 | 0.002 | 0.334 | 0.023 | 0.334 | 0.000 |
| RISK | RISK | 0.061 | 0.003 | 0.679 | 0.035 | 0.679 | 0.000 |
| OUD | OUD | 0.052 | 0.019 | 0.418 | 0.151 | 0.418 | 0.006 |
| CUD | CUD | 0.008 | 0.010 | 0.077 | 0.089 | 0.077 | 0.389 |
| PAU | PAU | 0.032 | 0.003 | 0.502 | 0.052 | 0.502 | 0.000 |
| PTU | PTU | 0.108 | 0.032 | 0.799 | 0.235 | 0.799 | 0.001 |
| SI | PTU | 0.005 | 0.006 | 0.039 | 0.047 | 0.118 | 0.410 |
| DPW | PAU | 0.019 | 0.002 | 0.439 | 0.036 | 0.666 | 0.000 |
| CI | CUD | 0.000 | 0.005 | 0.001 | 0.045 | 0.005 | 0.982 |
| SUD | SUD | 0.000 | 0.000 | 0.000 | NA | NA | NA |
| SUB | SUB | 0.000 | 0.000 | 0.000 | NA | NA | NA |
| BD | BD | 0.000 | 0.000 | 0.000 | NA | NA | NA |
| **Variances and covariances** | | | | | | | |
| SUBG1b | SUBG1b | 1.000 | NA | 1.000 | NA | 1.000 | NA |
| SUDResG1b | SUDResG1b | 1.000 | NA | 1.000 | NA | 1.000 | NA |
| BDResG1b | BDResG1b | 1.000 | NA | 1.000 | NA | 1.000 | NA |

Supplementary Table 11.

| **Factor**/**Indicator** | | **Unstd. Est.** | **Unstd. SE** | **Std. Genotype** | **Std. Genotype SE** | **Std. All** | ***P*-value** |
| --- | --- | --- | --- | --- | --- | --- | --- |
| **Loadings** | | | | | | | |
| BD | FSEX | 0.308 | 0.007 | 0.793 | 0.018 | 0.793 | < 5e-300 |
|  | ADHD | 0.3 | 0.014 | 0.666 | 0.032 | 0.666 | 5.79E-96 |
|  | NSEX | 0.256 | 0.006 | 0.812 | 0.018 | 0.812 | < 5e-300 |
|  | RISK | 0.171 | 0.008 | 0.57 | 0.026 | 0.57 | 1.71E-104 |
| SUB | CI | 0.243 | 0.01 | 0.67 | 0.029 | 0.67 | 9.29E-120 |
|  | DPW | 0.058 | 0.004 | 0.347 | 0.022 | 0.347 | 2.03E-54 |
|  | SI | 0.306 | 0.008 | 0.946 | 0.025 | 0.946 | 4.38E-304 |
| SUD | PTU | 0.159 | 0.023 | 0.434 | 0.062 | 0.434 | 2.28E-12 |
|  | CUD | 0.296 | 0.014 | 0.9 | 0.042 | 0.9 | 4.01E-100 |
|  | PAU | 0.163 | 0.008 | 0.646 | 0.031 | 0.646 | 3.32E-97 |
| **Residuals** | | | | | | | |
| CI | CI | 0.072 | 0.007 | 0.551 | 0.05 | 0.551 | 3.40E-28 |
| SI | SI | 0.011 | 0.004 | 0.106 | 0.034 | 0.106 | 1.77E-03 |
| FSEX | FSEX | 0.056 | 0.003 | 0.371 | 0.021 | 0.371 | 3.33E-68 |
| ADHD | ADHD | 0.113 | 0.008 | 0.556 | 0.041 | 0.556 | 2.45E-41 |
| NSEX | NSEX | 0.034 | 0.002 | 0.341 | 0.023 | 0.341 | 3.10E-50 |
| RISK | RISK | 0.061 | 0.003 | 0.675 | 0.035 | 0.675 | 6.37E-85 |
| CUD | CUD | 0.021 | 0.01 | 0.19 | 0.09 | 0.19 | 3.60E-02 |
| PAU | PAU | 0.037 | 0.003 | 0.583 | 0.054 | 0.583 | 2.18E-27 |
| PTU | PTU | 0.109 | 0.032 | 0.812 | 0.235 | 0.812 | 5.53E-04 |
| DPW | DPW | 0.024 | 0.001 | 0.88 | 0.044 | 0.88 | 4.71E-89 |
| CI | CUD | 0 | 0.006 | 0 | 0.047 | 0 | 9.97E-01 |
| SI | PTU | 0.004 | 0.006 | 0.035 | 0.049 | 0.119 | 4.80E-01 |
| DPW | PAU | 0.019 | 0.002 | 0.458 | 0.036 | 0.64 | 6.34E-37 |
| **Variances & Covariances** | | | | | | | |
| BD | BD | 1.000 | NA | 1.000 | NA | 1.000 | NA |
| SUB | SUB | 1.000 | NA | 1.000 | NA | 1.000 | NA |
| SUD | SUD | 1.000 | NA | 1.000 | NA | 1.000 | NA |
| BD | SUB | 0.773 | 0.017 | 0.773 | 0.017 | 0.773 | < 5e-300 |
| BD | SUD | 0.849 | 0.035 | 0.849 | 0.035 | 0.849 | 5.58E-132 |
| SUB | SUD | 0.861 | 0.033 | 0.861 | 0.033 | 0.861 | 3.55E-153 |

Supplementary Table 12

| **Factor**/**Indicator** | | **Unstd. Est.** | **Unstd. SE** | **Std. Genotype** | **Std. Genotype SE** | **Std. All** | ***P*-value** |
| --- | --- | --- | --- | --- | --- | --- | --- |
| **Higher-order Loadings** | | | | | | | |
| SUDG1c | SUD | 0.159 | 0.023 | 0.434 | 0.062 | 0.434 | 0.000 |
|  | SUB | 0.209 | 0.012 | 0.577 | 0.034 | 0.861 | 0.000 |
|  | BD | 0.261 | 0.012 | 0.673 | 0.030 | 0.849 | 0.000 |
| SUBResG1c | SUB | 0.124 | 0.014 | 0.341 | 0.039 | 0.509 | 0.000 |
|  | BD | 0.026 | 0.029 | 0.066 | 0.076 | 0.083 | 0.383 |
| BDResG1c | BD | 0.161 | 0.014 | 0.414 | 0.037 | 0.522 | 0.000 |
| **Lower-order Loadings** | | | | | | | |
| SUD | PTU | 1.00 | NA | 1.00 | NA | 1.00 | NA |
|  | CUD | 1.864 | 0.268 | 2.076 | 0.299 | 0.900 | 0.000 |
|  | PAU | 1.024 | 0.146 | 1.490 | 0.213 | 0.646 | 0.000 |
| SUB | CI | 1.000 | NA | 1.000 | NA | 0.670 | NA |
|  | SI | 1.261 | 0.057 | 1.411 | 0.063 | 0.946 | 0.000 |
|  | DPW | 0.237 | 0.017 | 0.517 | 0.037 | 0.347 | 0.000 |
| BD | FSEX | 1.000 | NA | 1.000 | NA | 0.793 | NA |
|  | ADHD | 0.976 | 0.052 | 0.840 | 0.045 | 0.666 | 0.000 |
|  | NSEX | 0.831 | 0.021 | 1.023 | 0.026 | 0.812 | < 5e-300 |
|  | RISK | 0.556 | 0.024 | 0.719 | 0.031 | 0.570 | 0.000 |
| **Residuals** | | | | | | | |
| CI | CI | 0.072 | 0.007 | 0.551 | 0.050 | 0.551 | 0.000 |
| DPW | DPW | 0.024 | 0.001 | 0.880 | 0.044 | 0.880 | 0.000 |
| SI | SI | 0.011 | 0.004 | 0.106 | 0.034 | 0.106 | 0.002 |
| DRUG | DRUG | NA | NA | NA | NA | NA | NA |
| FSEX | FSEX | 0.056 | 0.003 | 0.371 | 0.021 | 0.371 | 0.000 |
| ADHD | ADHD | 0.113 | 0.008 | 0.556 | 0.041 | 0.556 | 0.000 |
| NSEX | NSEX | 0.034 | 0.002 | 0.341 | 0.023 | 0.341 | 0.000 |
| RISK | RISK | 0.061 | 0.003 | 0.675 | 0.035 | 0.675 | 0.000 |
| OUD | OUD | NA | NA | NA | NA | NA | NA |
| CUD | CUD | 0.021 | 0.010 | 0.190 | 0.090 | 0.190 | 0.036 |
| PAU | PAU | 0.037 | 0.003 | 0.583 | 0.054 | 0.583 | 0.000 |
| PTU | PTU | 0.109 | 0.032 | 0.812 | 0.235 | 0.812 | 0.001 |
| SI | PTU | 0.004 | 0.006 | 0.035 | 0.049 | 0.119 | 0.480 |
| DPW | PAU | 0.019 | 0.002 | 0.458 | 0.036 | 0.640 | 0.000 |
| CI | CUD | 0.000 | 0.006 | 0.000 | 0.047 | 0.000 | 0.997 |
| SUD | SUD | 0.000 | 0.000 | 0.000 | NA | NA | NA |
| SUB | SUB | 0.000 | 0.000 | 0.000 | NA | NA | NA |
| BD | BD | 0.000 | 0.000 | 0.000 | NA | NA | NA |
| **Variances and covariances** | | | | | | | |
| SUDG1c | SUDG1c | 1.000 | NA | 1.000 | NA | 1.000 | NA |
| SUBResG1c | SUBResG1c | 1.000 | NA | 1.000 | NA | 1.000 | NA |
| BDResG1c | BDResG1c | 1.000 | NA | 1.000 | NA | 1.000 | NA |

Supplementary Table 13

| **Factor**/**Indicator** | | **Unstd. Est.** | **Unstd. SE** | **Std. Genotype** | **Std. Genotype SE** | **Std. All** | ***P*-value** |
| --- | --- | --- | --- | --- | --- | --- | --- |
| **Higher-order Loadings** | | | | | | | |
| SUBG1d | SUB | 0.243 | 0.010 | 0.670 | 0.029 | 0.670 | 0.000 |
|  | SUD | 0.137 | 0.019 | 0.373 | 0.052 | 0.861 | 0.000 |
|  | BD | 0.238 | 0.007 | 0.613 | 0.018 | 0.773 | 0.000 |
| SUDResG1d | SUD | 0.081 | 0.016 | 0.221 | 0.043 | 0.509 | 0.000 |
|  | BD | 0.111 | 0.021 | 0.286 | 0.053 | 0.360 | 0.000 |
| BDResG1d | BD | 0.161 | 0.014 | 0.414 | 0.037 | 0.522 | 0.000 |
| **Lower-order Loadings** | | | | | | | |
| SUD | PTU | 1.000 | NA | 1.000 | NA | 0.434 | NA |
|  | CUD | 1.864 | 0.268 | 2.076 | 0.299 | 0.900 | 0.000 |
|  | PAU | 1.024 | 0.146 | 1.490 | 0.213 | 0.646 | 0.000 |
| SUB | CI | 1.000 | NA | 1.000 | NA | 0.670 | NA |
|  | SI | 1.261 | 0.057 | 1.411 | 0.063 | 0.946 | 0.000 |
|  | DPW | 0.237 | 0.017 | 0.517 | 0.037 | 0.347 | 0.000 |
| BD | FSEX | 1.000 | NA | 1.000 | NA | 0.793 | NA |
|  | ADHD | 0.976 | 0.052 | 0.840 | 0.045 | 0.666 | 0.000 |
|  | NSEX | 0.831 | 0.021 | 1.023 | 0.026 | 0.812 | < 5e-300 |
|  | RISK | 0.556 | 0.024 | 0.719 | 0.031 | 0.570 | 0.000 |
| **Residuals** | | | | | | | |
| CI | CI | 0.072 | 0.007 | 0.551 | 0.050 | 0.551 | 0.000 |
| DPW | DPW | 0.024 | 0.001 | 0.880 | 0.044 | 0.880 | 0.000 |
| SI | SI | 0.011 | 0.004 | 0.106 | 0.034 | 0.106 | 0.002 |
| FSEX | FSEX | 0.056 | 0.003 | 0.371 | 0.021 | 0.371 | 0.000 |
| ADHD | ADHD | 0.113 | 0.008 | 0.556 | 0.041 | 0.556 | 0.000 |
| NSEX | NSEX | 0.034 | 0.002 | 0.341 | 0.023 | 0.341 | 0.000 |
| RISK | RISK | 0.061 | 0.003 | 0.675 | 0.035 | 0.675 | 0.000 |
| CUD | CUD | 0.021 | 0.010 | 0.190 | 0.090 | 0.190 | 0.036 |
| PAU | PAU | 0.037 | 0.003 | 0.583 | 0.054 | 0.583 | 0.000 |
| PTU | PTU | 0.109 | 0.032 | 0.812 | 0.235 | 0.812 | 0.001 |
| SI | PTU | 0.004 | 0.006 | 0.035 | 0.049 | 0.119 | 0.480 |
| DPW | PAU | 0.019 | 0.002 | 0.458 | 0.036 | 0.640 | 0.000 |
| CI | CUD | 0.000 | 0.006 | 0.000 | 0.047 | 0.000 | 0.997 |
| SUD | SUD | 0.000 | 0.000 | 0.000 | NA | NA | NA |
| SUB | SUB | 0.000 | 0.000 | 0.000 | NA | NA | NA |
| BD | BD | 0.000 | 0.000 | 0.000 | NA | NA | NA |
| **Variances and covariances** | | | | | | | |
| SUBG1d | SUBG1d | 1.000 | NA | 1.000 | NA | 1.000 | NA |
| SUDResG1d | SUDResG1d | 1.000 | NA | 1.000 | NA | 1.000 | NA |
| BDResG1d | BDResG1d | 1.000 | NA | 1.000 | NA | 1.000 | NA |

Supplementary Table 14

| **Factor**/**Indicator** | | **Unstd. Est.** | **Unstd. SE** | **Std. Genotype** | **Std. Genotype SE** | **Std. All** | ***P-value*** |
| --- | --- | --- | --- | --- | --- | --- | --- |
| **Loadings** | | | | | | | |
| BD | FSEX | 0.308 | 0.007 | 0.793 | 0.018 | 0.793 | < 5e-300 |
|  | ADHD | 0.294 | 0.014 | 0.652 | 0.031 | 0.652 | 1.06E-95 |
|  | NSEX | 0.259 | 0.006 | 0.818 | 0.018 | 0.818 | < 5e-300 |
|  | RISK | 0.171 | 0.008 | 0.569 | 0.026 | 0.569 | 6.31E-105 |
| SUB | CI | 0.237 | 0.01 | 0.652 | 0.029 | 0.652 | 8.21E-114 |
|  | DPW | 0.067 | 0.004 | 0.402 | 0.022 | 0.402 | 2.34E-75 |
|  | SI | 0.297 | 0.008 | 0.909 | 0.023 | 0.909 | < 5e-300 |
|  | DRUG | 0.317 | 0.022 | 0.928 | 0.066 | 0.928 | 2.65E-45 |
| SUD | PTU | 0.165 | 0.021 | 0.45 | 0.058 | 0.45 | 6.81E-15 |
|  | CUD | 0.311 | 0.012 | 0.938 | 0.037 | 0.938 | 1.18E-138 |
|  | PAU | 0.184 | 0.007 | 0.728 | 0.027 | 0.728 | 1.09E-158 |
|  | OUD | 0.264 | 0.016 | 0.75 | 0.045 | 0.75 | 3.21E-62 |
| **Residuals** | | | | | | | |
| DPW | DPW | 0.024 | 0.001 | 0.838 | 0.042 | 0.838 | 1.15E-88 |
| DRUG | DRUG | 0.016 | 0.02 | 0.138 | 0.17 | 0.138 | 0.41722022 |
| CI | CI | 0.076 | 0.007 | 0.575 | 0.05 | 0.575 | 3.57E-30 |
| SI | SI | 0.019 | 0.003 | 0.174 | 0.029 | 0.174 | 1.09E-09 |
| FSEX | FSEX | 0.056 | 0.003 | 0.371 | 0.022 | 0.371 | 1.07E-62 |
| ADHD | ADHD | 0.117 | 0.008 | 0.575 | 0.041 | 0.575 | 7.95E-45 |
| NSEX | NSEX | 0.033 | 0.002 | 0.33 | 0.023 | 0.33 | 3.76E-47 |
| RISK | RISK | 0.061 | 0.003 | 0.676 | 0.035 | 0.676 | 7.70E-83 |
| OUD | OUD | 0.054 | 0.019 | 0.437 | 0.151 | 0.437 | 0.00376914 |
| CUD | CUD | 0.013 | 0.01 | 0.12 | 0.087 | 0.12 | 0.16972108 |
| PAU | PAU | 0.03 | 0.003 | 0.47 | 0.051 | 0.47 | 4.57E-20 |
| PTU | PTU | 0.108 | 0.031 | 0.798 | 0.231 | 0.798 | 0.00056513 |
| **Variances & Covariances** | | | | | | | |
| BD | BD | 1.000 | NA | 1.000 | NA | 1.000 | NA |
| SUB | SUB | 1.000 | NA | 1.000 | NA | 1.000 | NA |
| SUD | SUD | 1.000 | NA | 1.000 | NA | 1.000 | NA |
| BD | SUB | 0.78 | 0.016 | 0.78 | 0.016 | 0.78 | < 5e-300 |
| BD | SUD | 0.77 | 0.027 | 0.77 | 0.027 | 0.77 | 7.98E-181 |
| SUB | SUD | 0.852 | 0.028 | 0.852 | 0.028 | 0.852 | 1.22E-201 |

Supplementary Table 15

| **Factor**/**Indicator** | | **Unstd. Est.** | **Unstd. SE** | **Std. Genotype** | **Std. Genotype SE** | **Std. All** | ***P*-value** |
| --- | --- | --- | --- | --- | --- | --- | --- |
| **Loadings** | | | | | | | |
| BD | FSEX | 0.307 | 0.007 | 0.790 | 0.018 | 0.790 | < 5e-300 |
|  | ADHD | 0.299 | 0.014 | 0.663 | 0.032 | 0.663 | 0.000 |
|  | NSEX | 0.256 | 0.006 | 0.815 | 0.018 | 0.815 | < 5e-300 |
|  | RISK | 0.172 | 0.008 | 0.574 | 0.026 | 0.574 | 0.000 |
| SUB | CI | 0.240 | 0.010 | 0.664 | 0.028 | 0.664 | 0.000 |
|  | DPW | 0.064 | 0.004 | 0.384 | 0.022 | 0.384 | 0.000 |
|  | SI | 0.297 | 0.008 | 0.916 | 0.025 | 0.916 | 0.000 |
| SUD | PTU | 0.158 | 0.021 | 0.432 | 0.057 | 0.432 | 0.000 |
|  | CUD | 0.288 | 0.013 | 0.874 | 0.041 | 0.874 | 0.000 |
|  | PAU | 0.168 | 0.008 | 0.666 | 0.031 | 0.666 | 0.000 |
| **Residuals** | | | | | | | |
| DPW | DPW | 0.023 | 0.001 | 0.852 | 0.043 | 0.852 | 0.000 |
| CI | CI | 0.073 | 0.007 | 0.559 | 0.050 | 0.559 | 0.000 |
| SI | SI | 0.017 | 0.003 | 0.161 | 0.031 | 0.161 | 0.000 |
| FSEX | FSEX | 0.056 | 0.003 | 0.375 | 0.021 | 0.375 | 0.000 |
| ADHD | ADHD | 0.114 | 0.008 | 0.561 | 0.041 | 0.561 | 0.000 |
| NSEX | NSEX | 0.033 | 0.002 | 0.336 | 0.023 | 0.336 | 0.000 |
| RISK | RISK | 0.060 | 0.003 | 0.671 | 0.035 | 0.671 | 0.000 |
| CUD | CUD | 0.026 | 0.010 | 0.237 | 0.089 | 0.237 | 0.008 |
| PAU | PAU | 0.035 | 0.003 | 0.556 | 0.054 | 0.556 | 0.000 |
| PTU | PTU | 0.109 | 0.031 | 0.813 | 0.231 | 0.813 | 0.000 |
| **Variances & Covariances** | | | | | | | |
| BD | BD | 1.000 | NA | 1.000 | NA | 1.000 | NA |
| SUB | SUB | 1.000 | NA | 1.000 | NA | 1.000 | NA |
| SUD | SUD | 1.000 | NA | 1.000 | NA | 1.000 | NA |
| BD | SUB | 0.780 | 0.017 | 0.780 | 0.017 | 0.780 | < 5e-300 |
| BD | SUD | 0.844 | 0.035 | 0.844 | 0.035 | 0.844 | 0.000 |
| SUB | SUD | 0.932 | 0.036 | 0.932 | 0.036 | 0.932 | 0.000 |

Supplementary Table 16

| **Factor**/**Indicator** | | **Unstd. Est.** | **Unstd. SE** | **Std. Genotype** | **Std. Genotype SE** | **Std. All** | ***P-value*** |
| --- | --- | --- | --- | --- | --- | --- | --- |
| **Higher-order Loadings** | | | | | | | |
| SUDG1a | SUD | 0.165 | 0.021 | 0.45 | 0.058 | 0.45 | 6.81E-15 |
|  | SUB | 0.202 | 0.011 | 0.556 | 0.031 | 0.852 | 1.28E-70 |
|  | BD | 0.237 | 0.009 | 0.611 | 0.024 | 0.77 | 2.99E-143 |
| SUBResG1a | SUB | 0.124 | 0.012 | 0.341 | 0.033 | 0.523 | 3.18E-25 |
|  | BD | 0.073 | 0.019 | 0.188 | 0.048 | 0.237 | 9.72E-05 |
| BDResG1a | BD | 0.182 | 0.008 | 0.47 | 0.02 | 0.592 | 1.18E-117 |
| **Lower-order Loadings** | | | | | | | |
| SUD | PTU | 1.00 | NA | 1.00 | NA | 0.45 | NA |
|  | CUD | 1.882 | 0.245 | 2.086 | 0.271 | 0.938 | 1.44E-14 |
|  | PAU | 1.111 | 0.145 | 1.619 | 0.211 | 0.728 | 1.81E-14 |
|  | OUD | 1.594 | 0.224 | 1.668 | 0.234 | 0.75 | 1.02E-12 |
| SUB | CI | 1.00 | NA | 1.00 | NA | 0.652 | NA |
|  | DRUG | 1.34 | 0.097 | 1.423 | 0.103 | 0.928 | 1.74E-43 |
|  | SI | 1.255 | 0.055 | 1.394 | 0.061 | 0.909 | 1.88E-114 |
|  | DPW | 0.285 | 0.018 | 0.616 | 0.039 | 0.402 | 1.58E-55 |
| BD | FSEX | 1.00 | NA | 1.00 | NA | 0.793 | NA |
|  | ADHD | 0.955 | 0.051 | 0.821 | 0.044 | 0.652 | 1.06E-78 |
|  | NSEX | 0.841 | 0.021 | 1.031 | 0.026 | 0.818 | < 5e-300 |
|  | RISK | 0.555 | 0.024 | 0.718 | 0.031 | 0.569 | 1.13E-117 |
| **Residuals** | | | | | | | |
| CI | CI | 0.076 | 0.007 | 0.575 | 0.05 | 0.575 | 3.57E-30 |
| DPW | DPW | 0.024 | 0.001 | 0.838 | 0.042 | 0.838 | 1.15E-88 |
| SI | SI | 0.019 | 0.003 | 0.174 | 0.029 | 0.174 | 1.09E-09 |
| DRUG | DRUG | 0.0161 | 0.0199 | 0.1382 | 0.1704 | 0.1382 | 0.417 |
| FSEX | FSEX | 0.056 | 0.003 | 0.371 | 0.022 | 0.371 | 1.07E-62 |
| ADHD | ADHD | 0.117 | 0.008 | 0.575 | 0.041 | 0.575 | 7.95E-45 |
| NSEX | NSEX | 0.033 | 0.002 | 0.33 | 0.023 | 0.33 | 3.76E-47 |
| RISK | RISK | 0.061 | 0.003 | 0.676 | 0.035 | 0.676 | 7.70E-83 |
| OUD | OUD | 0.054 | 0.019 | 0.437 | 0.151 | 0.437 | 0.004 |
| CUD | CUD | 0.013 | 0.01 | 0.12 | 0.087 | 0.12 | 0.17 |
| PAU | PAU | 0.03 | 0.003 | 0.47 | 0.051 | 0.47 | 4.57E-20 |
| PTU | PTU | 0.108 | 0.031 | 0.798 | 0.231 | 0.798 | 0.001 |
| SUD | SUD | 0 | NA | 0 | NA | 0 | NA |
| SUB | SUB | 0 | NA | 0 | NA | 0 | NA |
| BD | BD | 0 | NA | 0 | NA | 0 | NA |

Supplementary Table 17

| **Factor**/**Indicator** | | **Unstd. Est.** | **Unstd. SE** | **Std. Genotype** | **Std. Genotype SE** | **Std. All** | ***P-value*** |
| --- | --- | --- | --- | --- | --- | --- | --- |
| **Higher-order Loadings** | | | | | | | |
| SUBG1b | SUB | 0.237 | 0.01 | 0.652 | 0.029 | 0.652 | 8.22E-114 |
|  | SUD | 0.141 | 0.018 | 0.383 | 0.048 | 0.852 | 1.39E-15 |
|  | BD | 0.24 | 0.007 | 0.619 | 0.018 | 0.78 | 9.52E-269 |
| SUDResG1b | SUD | 0.086 | 0.015 | 0.235 | 0.04 | 0.523 | 4.95E-09 |
|  | BD | 0.062 | 0.015 | 0.16 | 0.038 | 0.201 | 2.36E-05 |
| BDResG1b | BD | 0.182 | 0.008 | 0.47 | 0.02 | 0.592 | 1.18E-117 |
| **Lower-order Loadings** | | | | | | | |
| SUB | CI | 1 | NA | 1 | NA | 0.652 | NA |
|  | DRUG | 1.34 | 0.097 | 1.423 | 0.103 | 0.928 | 1.74E-43 |
|  | SI | 1.255 | 0.055 | 1.394 | 0.061 | 0.909 | 1.90E-114 |
|  | DPW | 0.285 | 0.018 | 0.616 | 0.039 | 0.402 | 1.58E-55 |
| SUD | PTU | 1 | NA | 1 | NA | 0.45 | NA |
|  | CUD | 1.882 | 0.245 | 2.086 | 0.271 | 0.938 | 1.44E-14 |
|  | PAU | 1.111 | 0.145 | 1.619 | 0.211 | 0.728 | 1.81E-14 |
|  | OUD | 1.594 | 0.224 | 1.668 | 0.234 | 0.75 | 1.02E-12 |
| BD | FSEX | 1 | NA | 1 | NA | 0.793 | NA |
|  | ADHD | 0.955 | 0.051 | 0.821 | 0.044 | 0.652 | 1.06E-78 |
|  | NSEX | 0.841 | 0.021 | 1.031 | 0.026 | 0.818 | < 5e-300 |
|  | RISK | 0.555 | 0.024 | 0.718 | 0.031 | 0.569 | 1.13E-117 |
| **Residuals** | | | | | | | |
| CI | CI | 0.076 | 0.007 | 0.575 | 0.05 | 0.575 | 3.57E-30 |
| DPW | DPW | 0.024 | 0.001 | 0.838 | 0.042 | 0.838 | 0 |
| SI | SI | 0.019 | 0.003 | 0.174 | 0.029 | 0.174 | 1.09E-09 |
| DRUG | DRUG | 0.016 | 0.02 | 0.138 | 0.17 | 0.138 | 4.17E-01 |
| FSEX | FSEX | 0.056 | 0.003 | 0.371 | 0.022 | 0.371 | 1.07E-62 |
| ADHD | ADHD | 0.117 | 0.008 | 0.575 | 0.041 | 0.575 | 7.95E-45 |
| NSEX | NSEX | 0.033 | 0.002 | 0.33 | 0.023 | 0.33 | 3.76E-47 |
| RISK | RISK | 0.061 | 0.003 | 0.676 | 0.035 | 0.676 | 7.70E-83 |
| OUD | OUD | 0.054 | 0.019 | 0.437 | 0.151 | 0.437 | 3.77E-03 |
| CUD | CUD | 0.013 | 0.01 | 0.12 | 0.087 | 0.12 | 1.70E-01 |
| PAU | PAU | 0.03 | 0.003 | 0.47 | 0.051 | 0.47 | 4.57E-20 |
| PTU | PTU | 0.108 | 0.031 | 0.798 | 0.231 | 0.798 | 5.65E-04 |
| BD | SUD | 0 | NA | 0 | NA | 0 | NA |
| BD | SUB | 0 | NA | 0 | NA | 0 | NA |
| SUB | SUD | 0 | NA | 0 | NA | 0 | NA |

Supplementary Table 18

| **Factor**/**Indicator** | | **Unstd. Est.** | **Unstd. SE** | **Std. Genotype** | **Std. Genotype SE** | **Std. All** | ***P*-value** |
| --- | --- | --- | --- | --- | --- | --- | --- |
| **Higher-order Loadings** | | | | | | | |
| SUDG1c | SUD | 0.158 | 0.021 | 0.432 | 0.057 | 0.432 | 0.000 |
|  | SUB | 0.224 | 0.013 | 0.619 | 0.035 | 0.932 | 0.000 |
|  | BD | 0.259 | 0.011 | 0.667 | 0.030 | 0.844 | 0.000 |
| SUBResG1c | SUB | 0.087 | 0.023 | 0.240 | 0.062 | 0.361 | 0.000 |
|  | BD | -0.006 | 0.050 | -0.015 | 0.128 | -0.019 | 0.905 |
| BDResG1c | BD | 0.164 | 0.019 | 0.423 | 0.050 | 0.535 | 0.000 |
| **Lower-order Loadings** | | | | | | | |
| SUD | PTU | 1.000 | NA | 1.000 | NA | 0.432 | NA |
|  | CUD | 1.816 | 0.238 | 2.022 | 0.265 | 0.874 | 0.000 |
|  | PAU | 1.060 | 0.139 | 1.542 | 0.202 | 0.666 | 0.000 |
| SUB | CI | 1.000 | NA | 1.000 | NA | 0.664 | NA |
|  | SI | 1.233 | 0.054 | 1.380 | 0.060 | 0.916 | 0.000 |
|  | DPW | 0.265 | 0.018 | 0.579 | 0.038 | 0.384 | 0.000 |
| BD | FSEX | 1.000 | NA | 1.000 | NA | 0.790 | NA |
|  | ADHD | 0.974 | 0.052 | 0.839 | 0.045 | 0.663 | 0.000 |
|  | NSEX | 0.836 | 0.021 | 1.031 | 0.026 | 0.815 | < 5e-300 |
|  | RISK | 0.562 | 0.024 | 0.726 | 0.031 | 0.574 | 0.000 |
| **Residuals** | | | | | | | |
| CI | CI | 0.073 | 0.007 | 0.559 | 0.050 | 0.559 | 0.000 |
| DPW | DPW | 0.023 | 0.001 | 0.852 | 0.043 | 0.852 | 0.000 |
| SI | SI | 0.017 | 0.003 | 0.161 | 0.031 | 0.161 | 0.000 |
| FSEX | FSEX | 0.056 | 0.003 | 0.375 | 0.021 | 0.375 | 0.000 |
| ADHD | ADHD | 0.114 | 0.008 | 0.561 | 0.041 | 0.561 | 0.000 |
| NSEX | NSEX | 0.033 | 0.002 | 0.336 | 0.023 | 0.336 | 0.000 |
| RISK | RISK | 0.060 | 0.003 | 0.671 | 0.035 | 0.671 | 0.000 |
| CUD | CUD | 0.026 | 0.010 | 0.237 | 0.089 | 0.237 | 0.008 |
| PAU | PAU | 0.035 | 0.003 | 0.556 | 0.054 | 0.556 | 0.000 |
| PTU | PTU | 0.109 | 0.031 | 0.813 | 0.231 | 0.813 | 0.000 |
| SUD | SUD | 0.000 | 0.000 | 0.000 | NA | NA | NA |
| SUB | SUB | 0.000 | 0.000 | 0.000 | NA | NA | NA |
| BD | BD | 0.000 | 0.000 | 0.000 | NA | NA | NA |
| **Variances and covariances** | | | | | | | |
| SUDG1c | SUDG1c | 1.000 | NA | 1.000 | NA | 1.000 | NA |
| SUBResG1c | SUBResG1c | 1.000 | NA | 1.000 | NA | 1.000 | NA |
| BDResG1c | BDResG1c | 1.000 | NA | 1.000 | NA | 1.000 | NA |

Supplementary Table 19

| **Factor**/**Indicator** | | **Unstd. Est.** | **Unstd. SE** | **Std. Genotype** | **Std. Genotype SE** | **Std. All** | ***P*-value** |
| --- | --- | --- | --- | --- | --- | --- | --- |
| **Higher-order Loadings** | | | | | | | |
| SUBG1d | SUB | 0.240 | 0.010 | 0.664 | 0.028 | 0.664 | 0.000 |
|  | SUD | 0.148 | 0.019 | 0.403 | 0.050 | 0.932 | 0.000 |
|  | BD | 0.239 | 0.007 | 0.617 | 0.018 | 0.780 | 0.000 |
| SUDResG1d | SUD | 0.057 | 0.018 | 0.156 | 0.050 | 0.361 | 0.002 |
|  | BD | 0.099 | 0.032 | 0.256 | 0.082 | 0.323 | 0.002 |
| BDResG1d | BD | 0.164 | 0.019 | 0.423 | 0.050 | 0.535 | 0.000 |
| **Lower-order Loadings** | | | | | | | |
| SUD | PTU | 1.000 | NA | 1.000 | NA | 0.432 | NA |
|  | CUD | 1.816 | 0.238 | 2.022 | 0.265 | 0.874 | 0.000 |
|  | PAU | 1.060 | 0.139 | 1.542 | 0.202 | 0.666 | 0.000 |
| SUB | CI | 1.000 | NA | 1.000 | NA | 0.664 | NA |
|  | SI | 1.233 | 0.054 | 1.380 | 0.060 | 0.916 | 0.000 |
|  | DPW | 0.265 | 0.018 | 0.579 | 0.038 | 0.384 | 0.000 |
| BD | FSEX | 1.000 | NA | 1.000 | NA | 0.790 | NA |
|  | ADHD | 0.974 | 0.052 | 0.839 | 0.045 | 0.663 | 0.000 |
|  | NSEX | 0.836 | 0.021 | 1.031 | 0.026 | 0.815 | < 5e-300 |
|  | RISK | 0.562 | 0.024 | 0.726 | 0.031 | 0.574 | 0.000 |
| **Residuals** | | | | | | | |
| CI | CI | 0.073 | 0.007 | 0.559 | 0.050 | 0.559 | 0.000 |
| DPW | DPW | 0.023 | 0.001 | 0.852 | 0.043 | 0.852 | 0.000 |
| SI | SI | 0.017 | 0.003 | 0.161 | 0.031 | 0.161 | 0.000 |
| FSEX | FSEX | 0.056 | 0.003 | 0.375 | 0.021 | 0.375 | 0.000 |
| ADHD | ADHD | 0.114 | 0.008 | 0.561 | 0.041 | 0.561 | 0.000 |
| NSEX | NSEX | 0.033 | 0.002 | 0.336 | 0.023 | 0.336 | 0.000 |
| RISK | RISK | 0.060 | 0.003 | 0.671 | 0.035 | 0.671 | 0.000 |
| CUD | CUD | 0.026 | 0.010 | 0.237 | 0.089 | 0.237 | 0.008 |
| PAU | PAU | 0.035 | 0.003 | 0.556 | 0.054 | 0.556 | 0.000 |
| PTU | PTU | 0.109 | 0.031 | 0.813 | 0.231 | 0.813 | 0.000 |
| SUD | SUD | 0.000 | NA | 0.000 | NA | 0.000 | NA |
| SUB | SUB | 0.000 | NA | 0.000 | NA | 0.000 | NA |
| BD | BD | 0.000 | NA | 0.000 | NA | 0.000 | NA |
| **Variances and covariances** | | | | | | | |
| SUBG1d | SUBG1d | 1.000 | NA | 1.000 | NA | 1.000 | NA |
| SUDResG1d | SUDResG1d | 1.000 | NA | 1.000 | NA | 1.000 | NA |
| BDResG1d | BDResG1d | 1.000 | NA | 1.000 | NA | 1.000 | NA |

Supplementary Table 20
