## Supplementary figures and images for "Partitioning the Genomic Components of Behavioral Disinhibition and Substance Use (Disorder) Using Genomic Structural Equation Modeling"

### Supplementary Figure 1

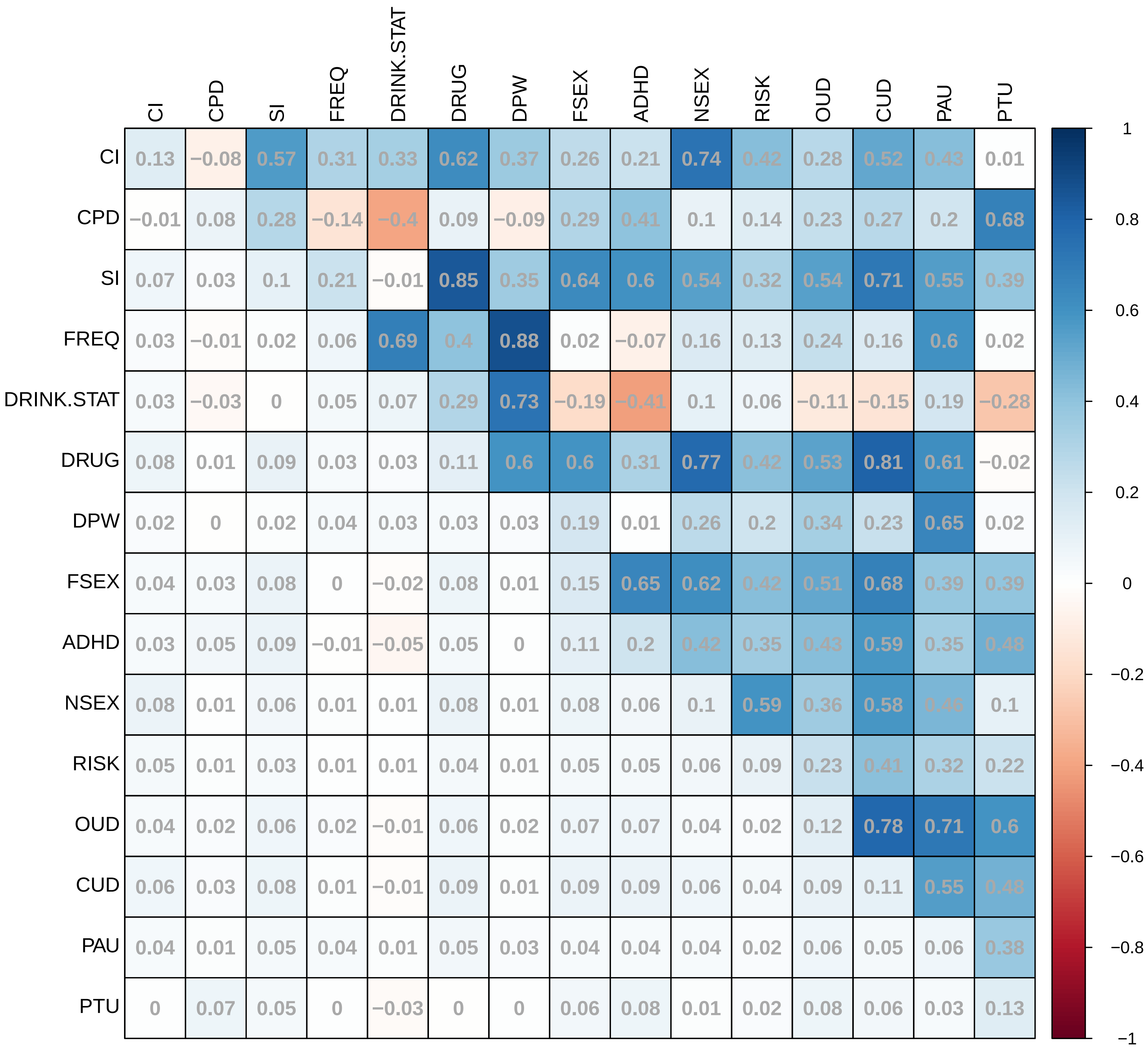

### Supplementary Figure 2A

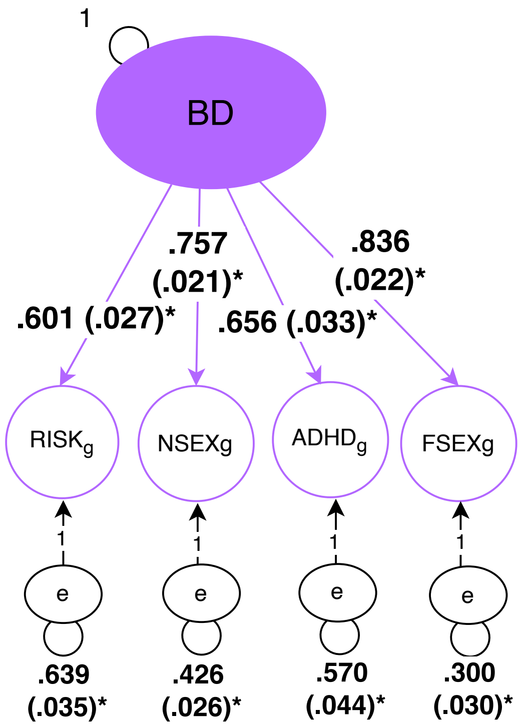

### Supplementary Figure 2B

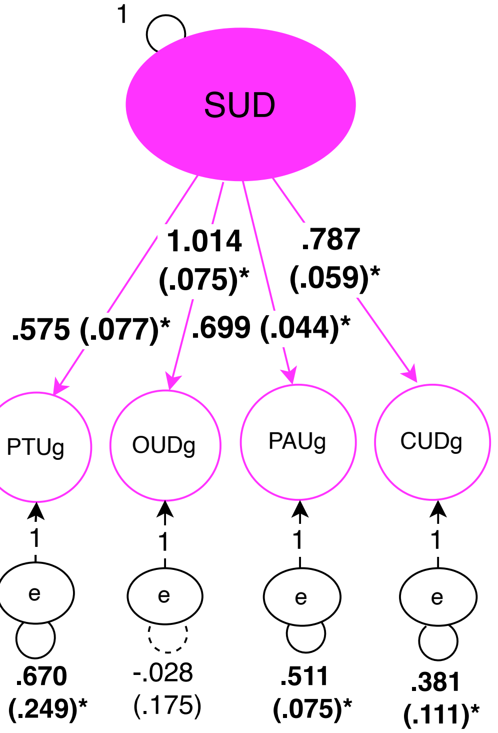

### Supplementary Figure 2C

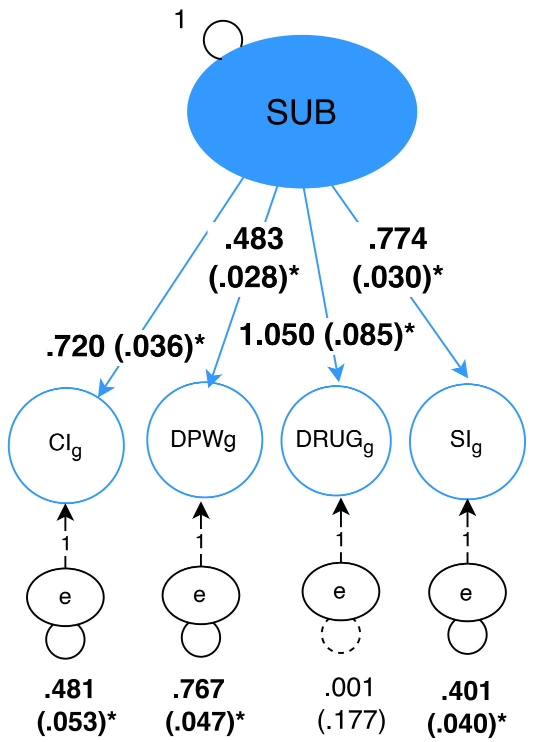

### Supplementary Figure 3

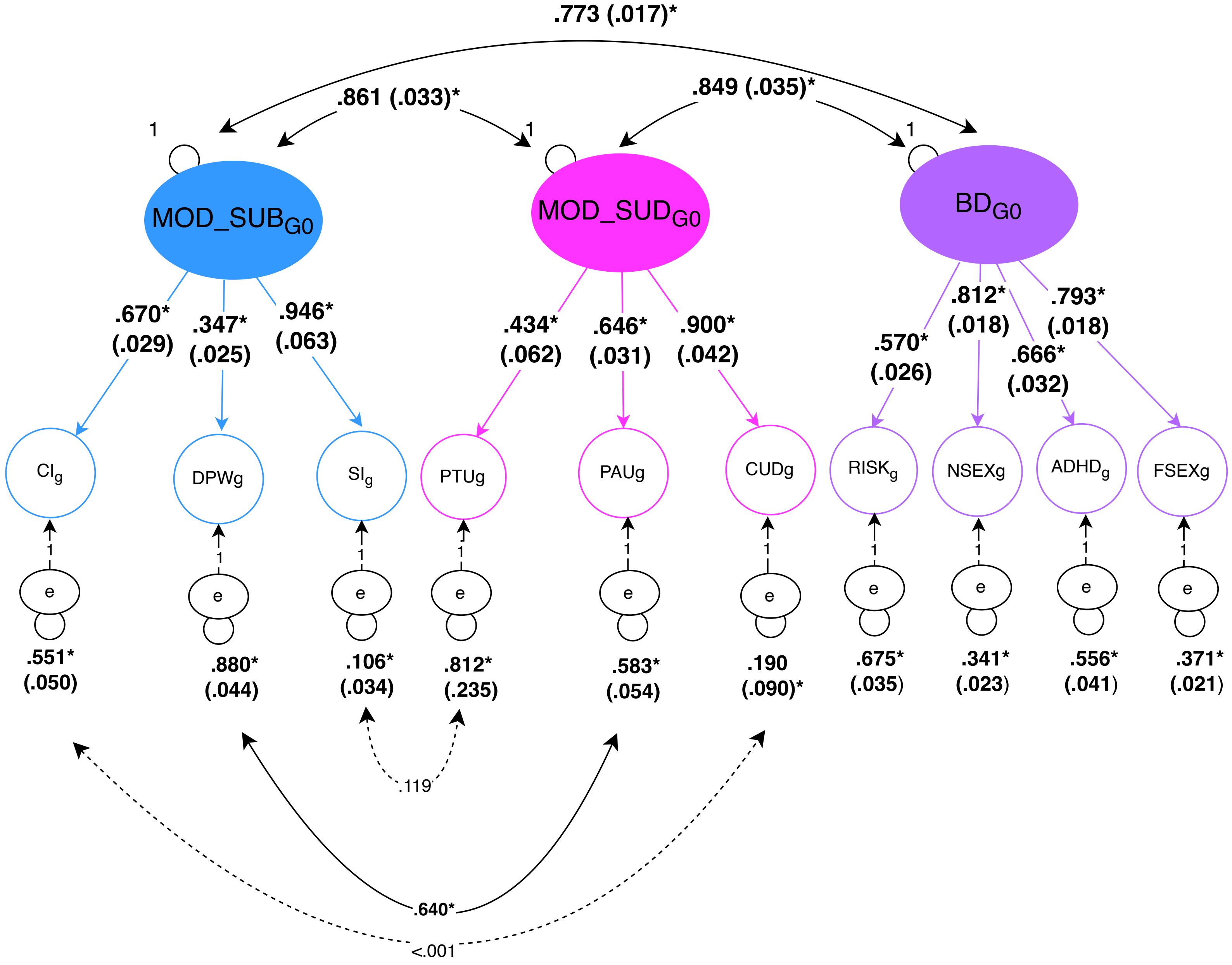

### Supplementary Figure 4A

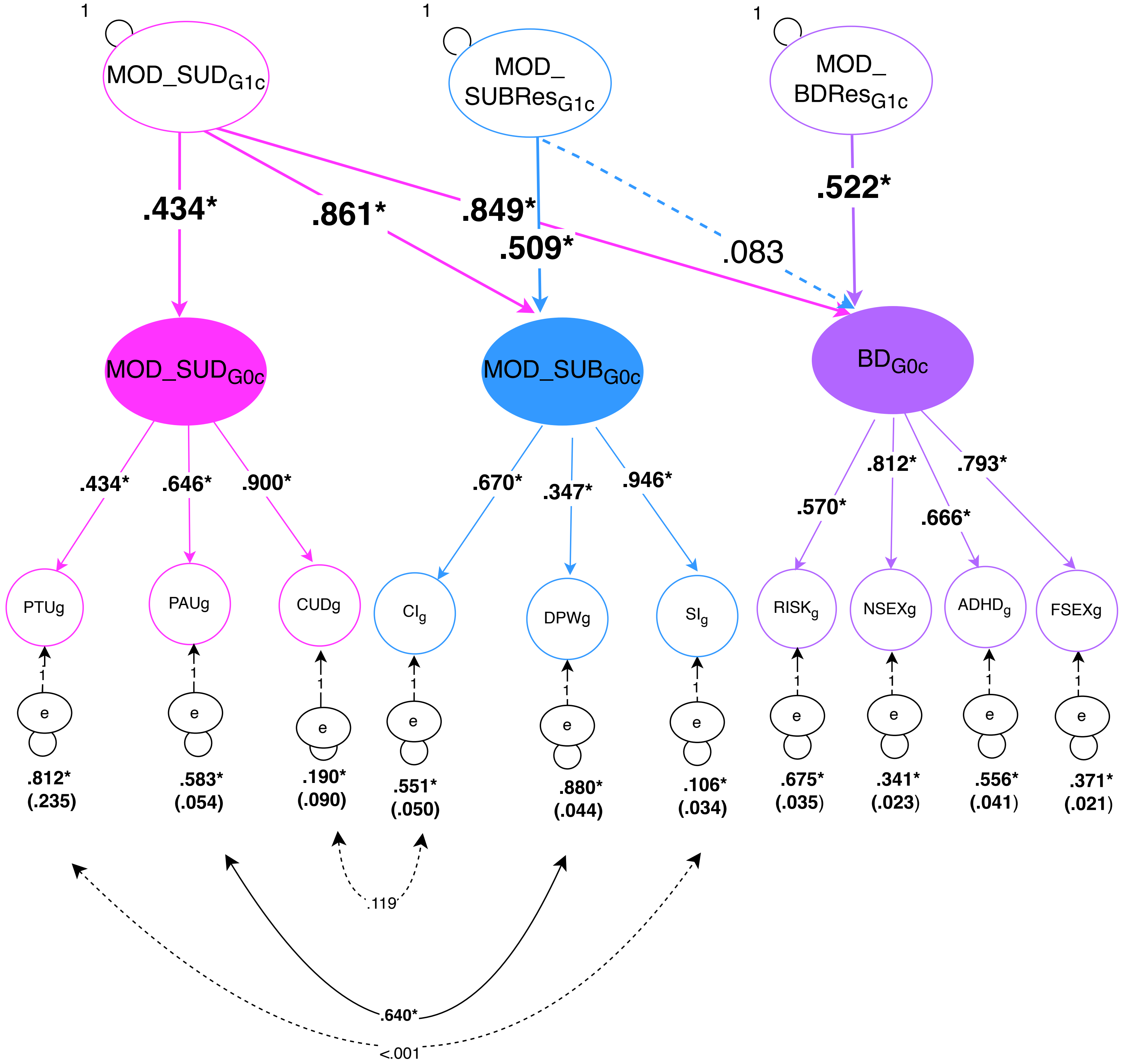

### Supplementary Figure 4B

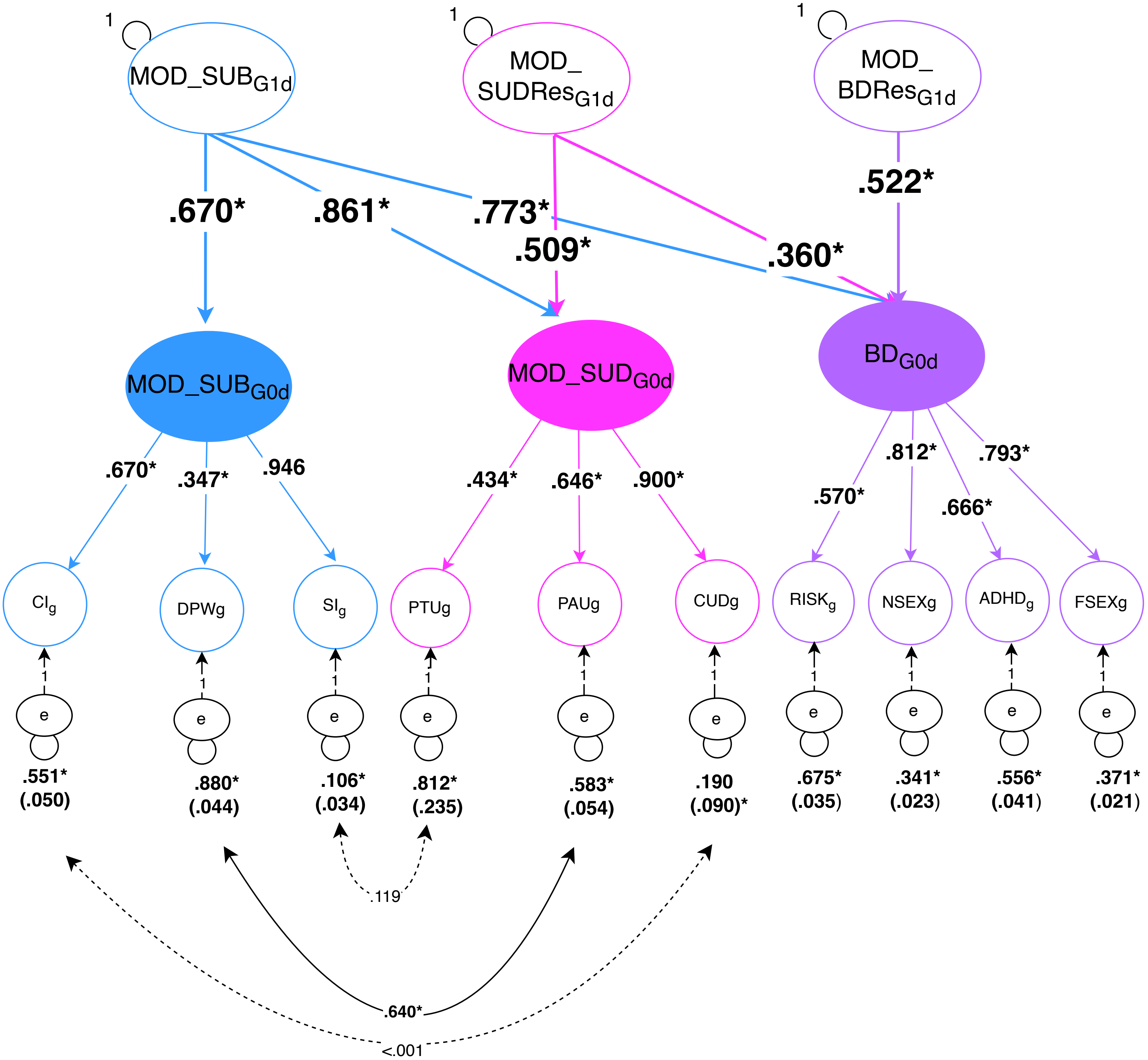

### Supplementary Figure 5

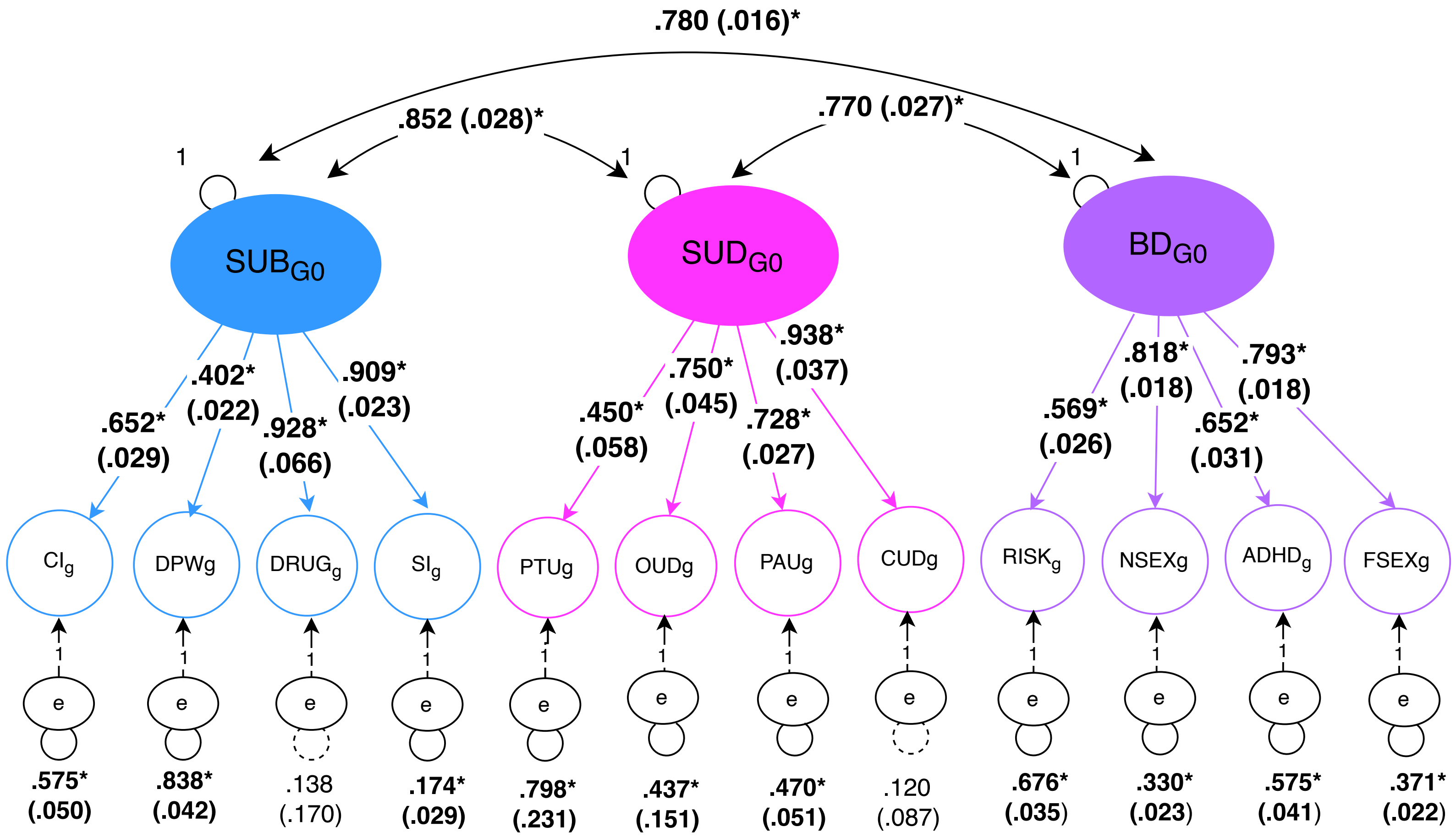

### Supplementary Figure 6

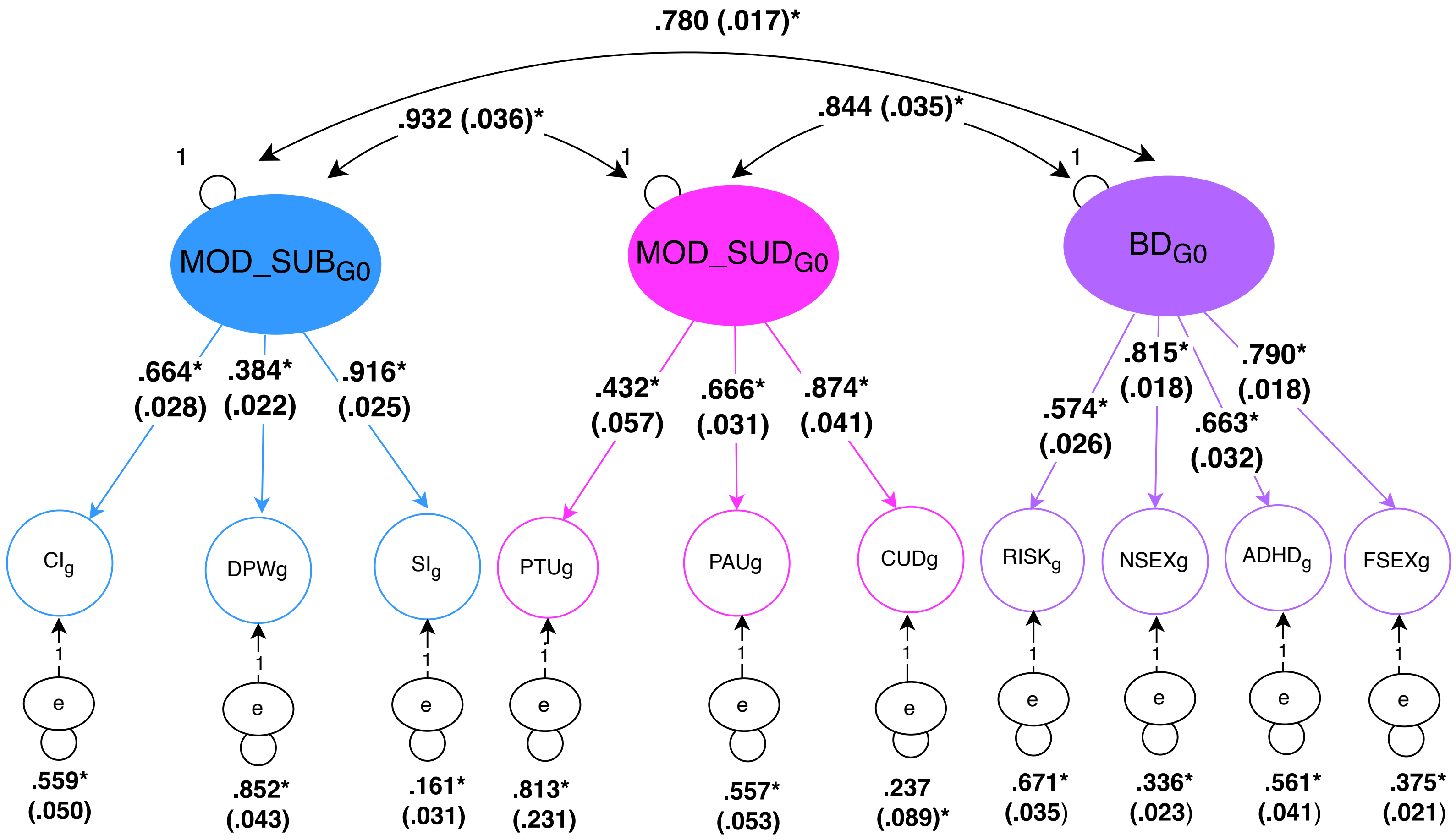

### Supplementary Figure 7A

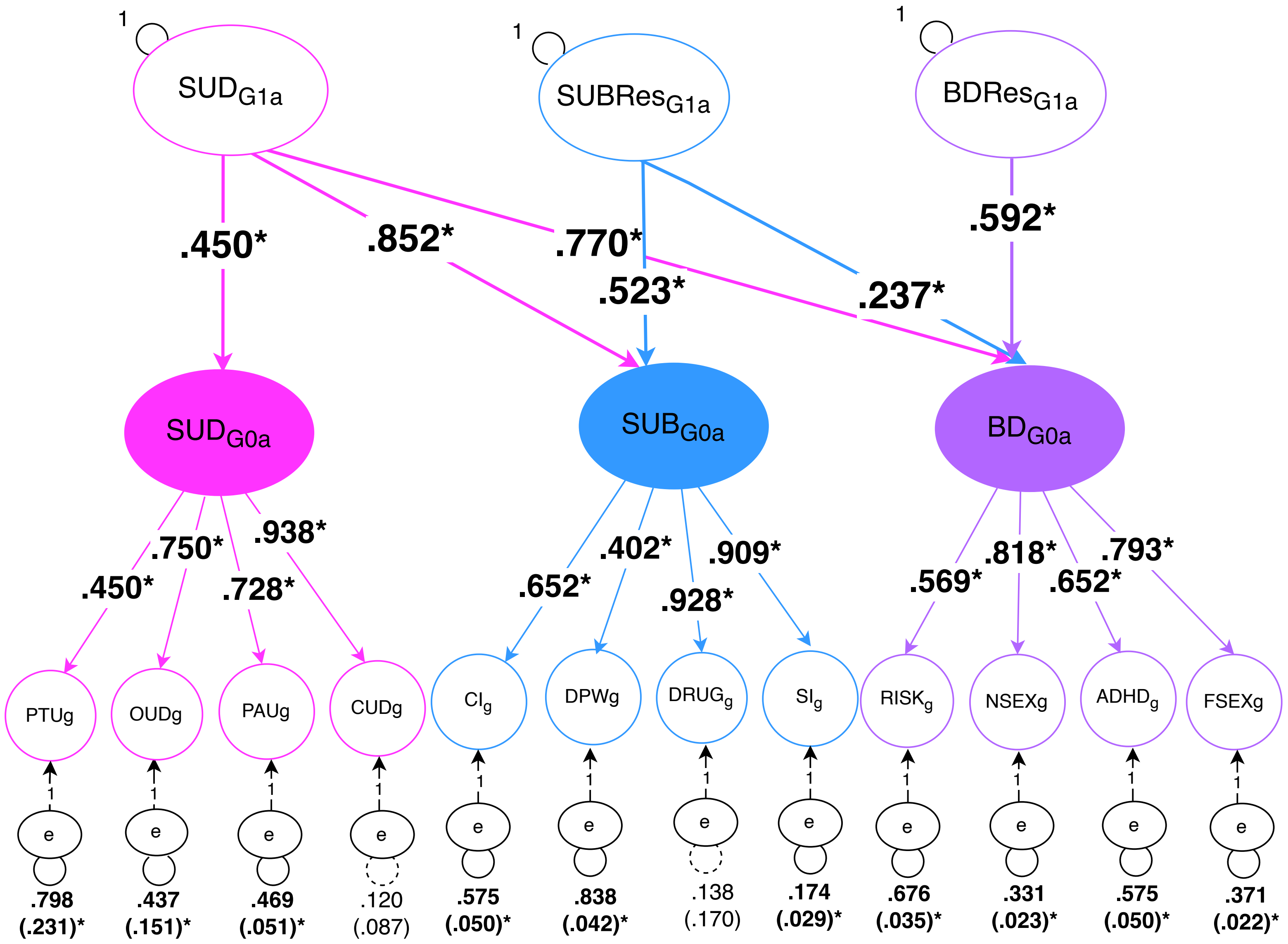

### Supplementary Figure 7B

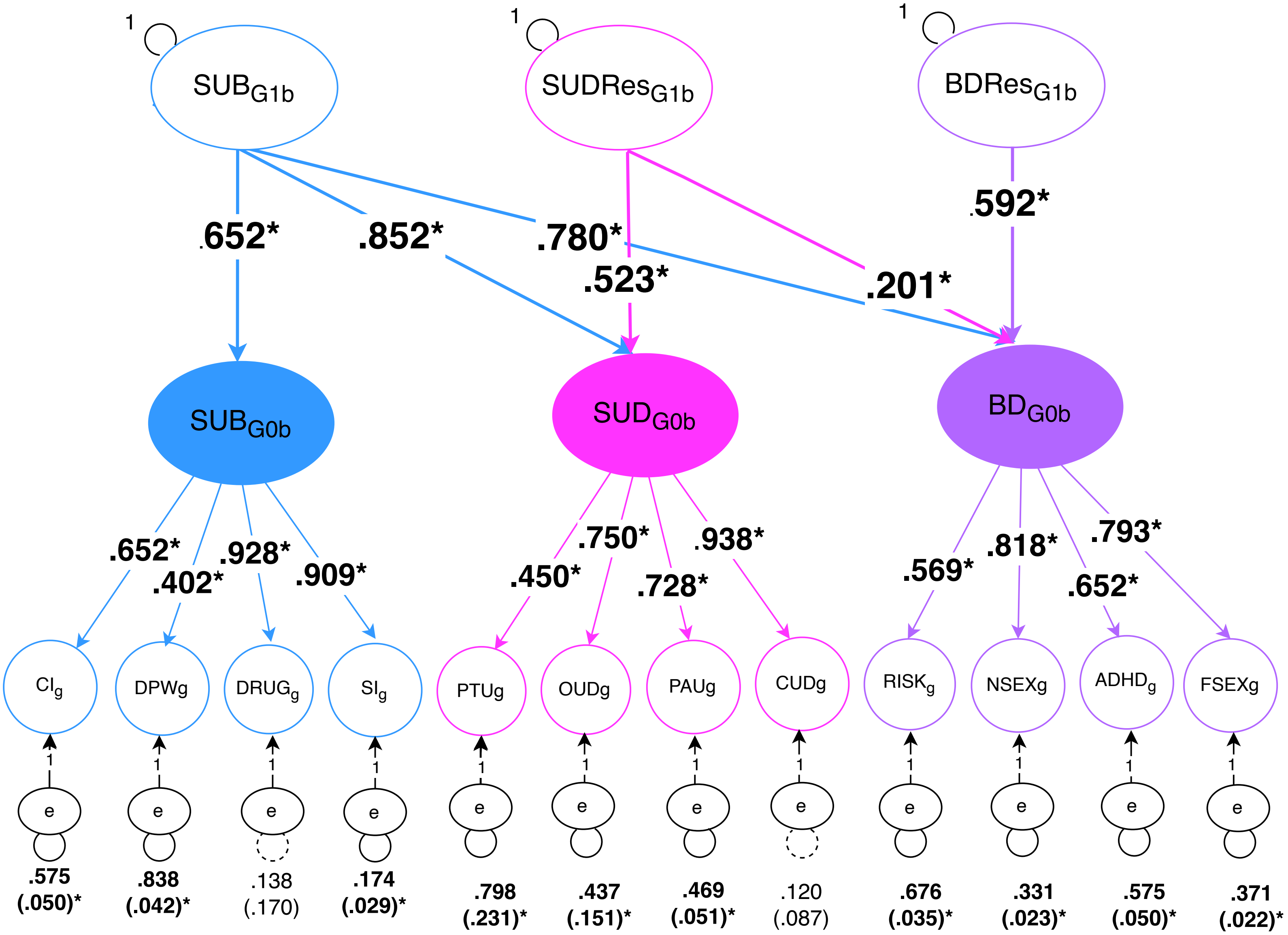

### Supplementary Figure 8A

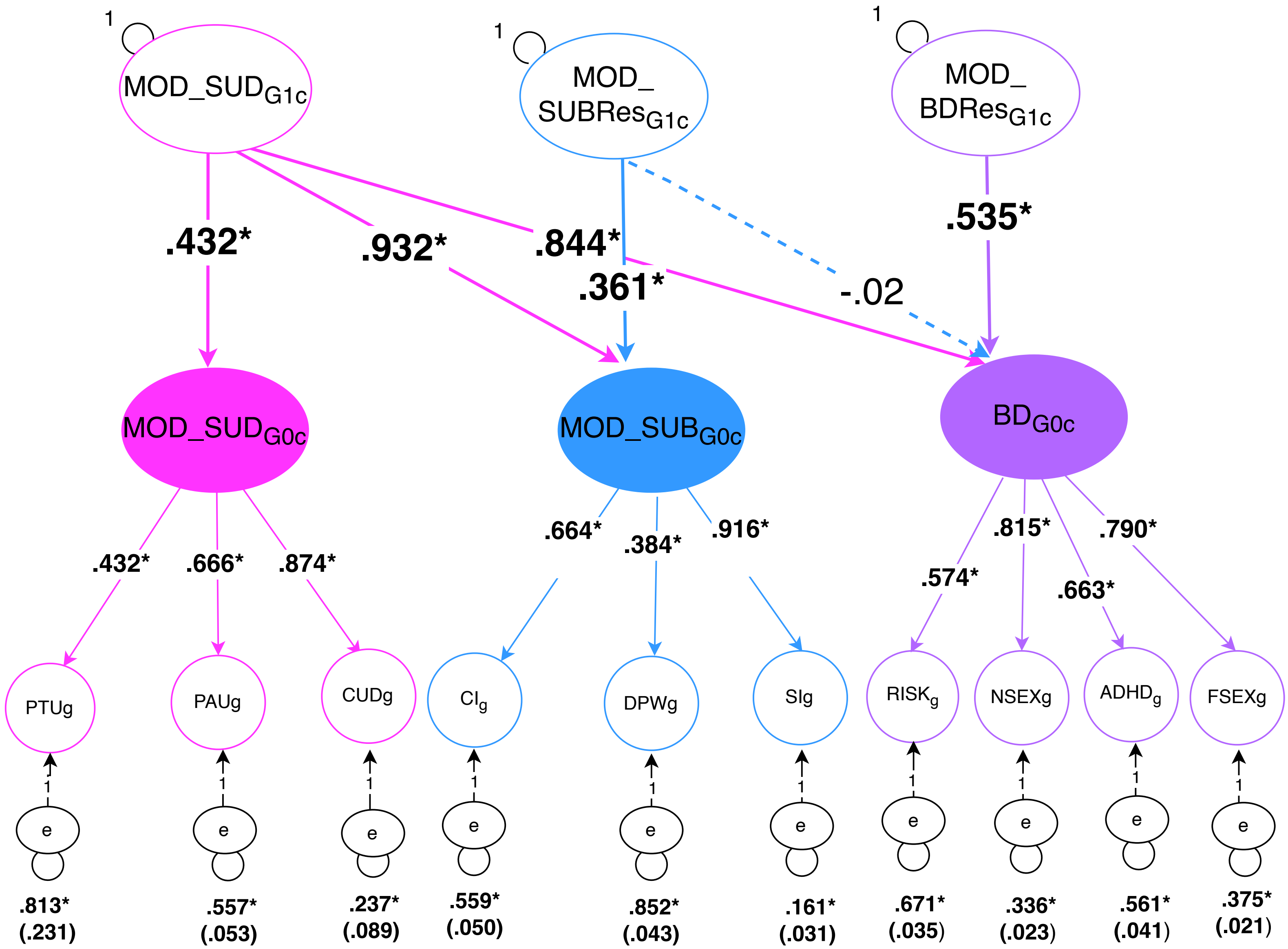

### Supplementary Figure 8B

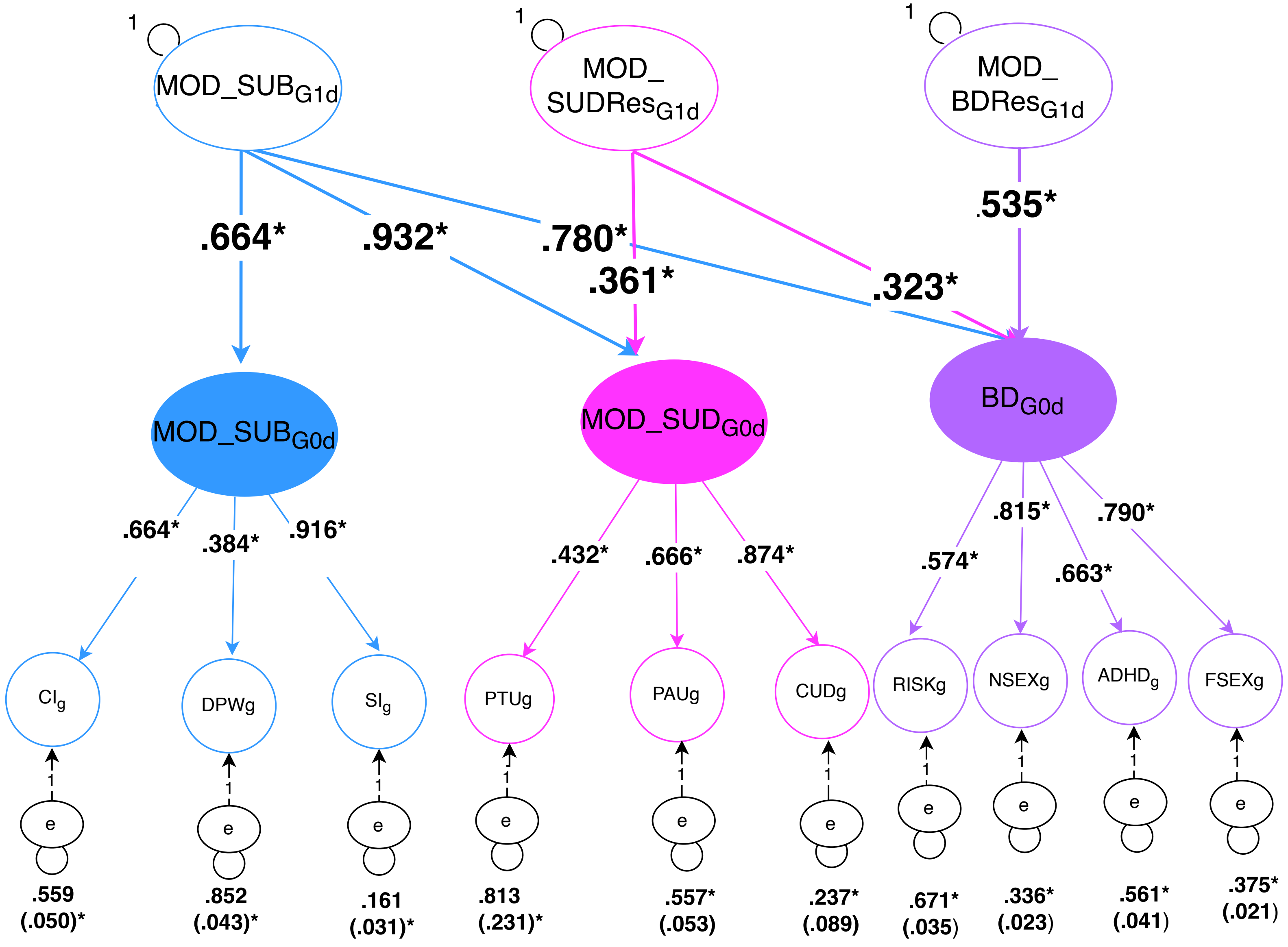
